# Supervised yogic intervention improves pain, cortical excitability and flexibility in fibromyalgia patients: An objective evidence based of journey from case control study to randomized controlled trial

**DOI:** 10.64898/2026.02.20.26346039

**Authors:** Aasheesh Kumar, Uma Kumar, Maroof Ahmad Khan, Raj Kumar Yadav, Akanksha Singh, Srikumar Venkataraman, Kishore Kumar Deepak, Rima Dada, Renu Bhatia

## Abstract

**Background and Aim:** Fibromyalgia is an idiopathic chronic widespread pain syndrome affecting 2-4% of the general population globally. Besides widespread fibromyalgia pain, morning stiffness, associated neurologic as well as sleep problems are also reported. Disease is more prevalent in females of middle-age group with low socioeconomic status, thus deteriorating overall productivity and psychosocial health. There is no permanent cure of the disease. This study aimed to explore, validate and assess the effect of four weeks of supervised yogic intervention on pain status, quality of life, sleep, cortical excitability, flexibility and range of motion in fibromyalgia patients, as compared to standard therapy.

**Method:** Case-control study, interventional study and assessor-blined randomized controlled trial, conducted in 120 fibromyalgia patients (60 yoga group: 60 waitlisted controls) and 60 age-matched healthy controls. Pain was assessed subjectively, using questionnaires and objectively, using quantitative sensory testing and ELISA. Sleep and quality of life were assessed using common and disease specific decsiptors. Flexibility and range of motion was assessed using sit and reach box, lateral goniometry and modified Schober’s test. Transcranial magnetic stimulation on M1 was used to assess corticomotor excitability of participants. Study parameters were assessed at baseline and after four weeks of the intervention.

**Results:** A significantly poor sleep, flexibility and quality of life was reported in the fibromyalgia patients due to excruciating pain (VAS = 6.92±0.12); corticomotor function was also abnormal in the patients, which were restored after four weeks of yogic intervention. On subjective and objective assessment of pain, we found significant relief and improvement in pain status in the yoga group as compared to the waitlisted controls. Fibromyalgia impact, sleep, quality of life and flexibility were also found solely better in fibromyalgia patients undergoing yogic interventions. Cortical parameters, specifically RMT, MEPs and MEP recruitment curves showed a significant improvement in yoga group as compared to waitlisted controls.

**Conclusion:** Four weeks of regular and supervised yogic intervention may ameliorate pain, improve flexibility and range of motion and changes cortical plasticity in the Indian cohort of fibromyalgia patients, as compared to standard therapy. Yoga-based interventions can also improve overall quality of life and sleep impairmentsby reducing catastrophization and fibromyalgia impact.

## Introduction

Fibromyalgia is an idiopathic chronic musculoskeletal pain syndrome prevalent globally including India. Incidence of fibromyalgia syndrome has been reported over 2% in the general population. [1–2] This disorder predominantly affects women aged 30-60 years in a ratio of 9:1 compared to men. [1, 3] It is interesting to note that men with fibromyalgia have less fatigue, morning stiffness and irritable bowel syndrome when compared to women indicating a role of female hormones in its symptomatology. [4, 5] Interestingly symptoms of fibromyalgia have also been reported in very few juveniles and clinical presentation is quite similar to the adult patients. [6] Besides excruciating widespread pain at 18 paired tender points of fibromyalgia, associated neurologic as well as sleep problems are also manifested in the patients at later times. Seeing the diverse symptomatology of fibromyalgia syndrome from fatigue, cognitive impairment, sleep problems and mood swing to brain fogging and depressive disorders, a vital role of central sensitization has been widely explored in some modern day research. [7] Some researchers put forward the concept of inefficient descending pain modulation associated with sensitisation of the central axis in fibromyalgia patients. [8–9] Despite the growing number of studies which explored the possible causative and risk factors, there are only a few studies which have been able to comment on the transition from acute to chronic pain in fibromyalgia.

Diagnosis of fibromyalgia shows a great dynamicity over time; earlier it was by the history of chronic widespread pain for at least 3 months and demonstration on minimum 11 tender points. [10] Currently fibromyalgia can be diagnosed and treated based on the widespread pain Index (WPI) and Symptom Severity Score (SSS) of the patient (American College of Rheumatology, 2016). However, the musculoskeletal and neurological examinations are normal in most of the fibromyalgia patients, so it is very difficult for the clinicians to find out exact causative targets of the disease. [11] There are varied proposed pathophysiology of the disease like oxidative burden, inflammation, sensitization due to chronic pain and tenderness; amongst them, most are contraindicatory and lack robust evidence, hence supporting the idiopathic nature of the disease. Few genes are also found to be involved in the pathophysiology of the disease such as SLC64A4, TRPV2 and many more; most of them are transporters and ion channels. [12–13]

Pain modulation plays a pivotal role in pain perception and its experience. Dysfunctional endogenous pain modulation is also a risk factor for the development and perpetuation of chronic pain conditions [14]. There is an increase in excitability of neurons of the central nervous system, specifically those receiving synaptic input from sensitized nociceptors. In contrast to peripheral sensitization, central mechanisms include recruitment of large (Aβ) fibers that contribute to pain and expansion of receptive fields. [15] Therefore sensory testing becomes an important tool to quantify the sensitivity driven by chronic pain and fatigue in fibromyalgia.

Altered nociceptive system characterized by hyperalgesia or allodynia is often associated with altered motor coordination of trunk muscles during postural and functional tasks [16], alterations in motor behaviour (altered gait), reduced activity of deep muscles [17] and increased activity of superficial trunk muscles [18–19]. Motor impairment and plastic changes in M1 and other cortical areas (premotor dorsal and ventral cortices), Supplementary Motor Area (SMA), cerebellum, basal ganglia, primary sensory cortex (S1) are also reported in many chronic musculoskeletal pain conditions. Transcranial magnetic stimulation is a widely used non-invasive technology that permits the investigation of the cortical excitability, functional organization, and integrity of the primary motor cortex (M1) that is largely involved in pain processing and motor control [20–21].

There is no permanent cure of the disease till date; treatment strategies available for the patients are only general in nature and include the use of both pharmacological and non-pharmacological symptoms for the management of fibromyalgia; former comprises of common medications like: acetaminophen, ibuprofen, naproxen, cyclobenzaprine, amitriptyline, and aspirin. [3] Some of the Food and Drug Administration (FDA) approved antidepressants and analgesics such as pregabalin, duloxetine and milnacipran help to control the symptoms of fibromyalgia syndrome temporarily. Few classical as well as modern day lifestyle and therapeutic interventions are extensively studied by the researchers across the globe: physical activity and exercise: stretching, aerobic and resistance exercises (EULAR Recommendation, 2019); massage and water therapy; yoga and meditation; magnetic stimulation, Transcutaneous Electric Nerve Stimulation (TENS) and vibrator therapy. [22–23] Management of fibromyalgia becomes more cumbersome as it has diverse symptoms associated from sleeplessness and fatigue to psychological distress. Some of the literature in the late 20^th^ century investigated the beneficial effect of exercises and yoga in fibromyalgia patients. Amongst the more novel management options for fibromyalgia non-invasive brain stimulation techniques such as repetitive Transcranial Magnetic Stimulation (rTMS) stands out due to a shifting etiological focus on central-level changes in fibromyalgia with high-rated successful randomized controlled trials. Most of the studies involved participants based on older diagnostic criteria and have used unoptimized repetitive transcranial magnetic stimulation paradigms. [24] Moreover, physical presence of the patients is compulsory to get brain stimulation therapy with the expensive equipments and highly expertised clinicians. [21, 25]

Yoga as a lifestyle intervention is an example of such treatment modality in several widespread musculoskeletal disorders such as chronic fatigue syndrome and chronic low back pain, including but not confined to fibromyalgia. [26–28] Yoga is a psychosomatic cost-effective and easy to adopt in day to day life intervention strategy with no or minimal side effects. However, its mechanism of action is not yet established. Since yoga can improve pain sensitivity, it would be logical to assume that yoga might be acting, at least in part, to favourably modulate the perceptual control over pain in fibromyalgia. Yoga fulfills both the criteria in line of clinical manifestations in fibromyalgia - i) associated musculoskeletal pain and fatigue and, ii) associated neurological as psychological disorders. To the best of our knowledge, none of the studies till date had systematically studied the effect of yogic intervention on corticomotor excitability and flexibility and range of motion in fibromyalgia patients. Though there are a number of subjective studies available which show its beneficial effect in ameliorating pain and improving quality of life and psychological distress; objective evidence for the beneficial role of yoga in pain and cortical excitability is missing in the literature.

The present study was aimed to explore the effect of four weeks of supervised yogic intervention on pain status, flexibilty and cortical excitability in fibromyalgia patients. To fulfill the aim, primary objectives undertaken were (i) To compare the measures of (a) Pain status; (b) Quality of life; (c) Flexibility and range of motion; (d) Cortical excitability, and (e) Levels of blood biomarkers (β- Endorphin, Cortisol, Glutamate, Serotonin and Substance P) of fibromyalgia patients with healthy participants (Case Control Study). (ii) To study the effect of yoga on the outcome parameters, mentioned in objective 1 in fibromyalgia patients (Interventional Study) and (iii) To compare the outcome parameters of objective 1 in yoga group and waitlisted control of fibromyalgia patients (Randomized Controlled Trial). Both objective and subjective measures of pain and associated symptoms of fibromyalgia were assessed before and after the four weeks of intervention. Therefore, this could reveal the possible mechanism of action of yoga as a pain management strategy in the Indian cohort of fibromyalgia patients.

## Methods

### Ethical Considerations and Clinical Trial Registration

The protocol for the study was approved by the Institute Ethical Committee, All India Institute of Medical Sciences, New Delhi, India (Ref. No. IECPG-611/28.10.2021). All the participants gave a written consent during enrolment in the study. Participants were duly explained the purpose and the details of the study intervention including the time and number of visits required by the participant, apriori. They were also given a Patient Information Sheet (PIS) to know more about their role in the clinical trial and data privacy. Participants were free to withdraw their consent at any stage without discosing the reason for premature discontinuation in the study. All necessary precautions were taken during the study at all time points and in case of any emergency, proper care was provided to all the participants free of cost. Patients were neither granted convenience for their visits on the day of assessment nor for attending the yoga classes. For maintaining the confidentiality and anonymity of the participants, they were assigned an alphanumeric Participants’ Identity Code (PIC) viz FM_YG_0XX, FM_WL_0XX, and HC_0XX; FM/HC representing fibromyalgia patients or healthy control followed by a two-alphabetic code denoting their group (yoga group/waitlisted controls) and three-digit index of enrolment in the study. Data was immediately stored in the cloud based sheets having multiple access to avoid loss of data at any instance. Randomized Controlled Trial (RCT) was registered with the apex regulatory organization of India under the umbrella of World Health Organization (WHO) – ‘Clinical Trial Registry of India (CTRI)’ (https://ctri.nic.in/). Application was submitted for prospective conductance of the study with reference number REF/11/2021/054093 and approval number CTRI/2023/06/054093.

### Study Design

The study was conducted at Pain Research and TMS Laboratory in the Department of Physiology, of a tertiary care hospital. The study design comprised of three objectives; first was a ‘cross sectional case control study’ wherein we compared measures of pain (both subjective and objective), quality of life, flexibility, corticomotor excitability and related blood biomarkers in 60 fibromyalgia patients with age and gender matched 60 healthy controls. Second objective was an interventional study conducted in 60 fibromyalgia patients and third, was an open-label Randomized Controlled Trial (RCT) in 120 fibromyalgia patients (60 in the yoga group and waitlisted controls, each). There were three non-parallel arms in the study despite any subgroupings: i) Healthy Controls (n = 60), ii) Yoga Group (n = 60), and iii) Waitlisted Controls (n = 60). Details of experimentations performed and study parameters in various arms are given in Supplementary Table 1 and Supplementary Table 2, respectively. Waitlisted controls were not disclosed about the criteria of enrolment in the study as they were offered the same yogic intervention of four weeks after wait period in which they were adviced to continue with their standard medications with follow-up recordings only after wait period not after offered yogic intervention.

### Randomization and Allocation for RCT

Patients were randomly allocated to yoga group and waitlisted controls (n = 60 each) based on the ‘Block Randomization’ performed using an online available randomizer tool (https://www.randomizer.org/). Computer generated 20 random blocks of block size 6 were generated using a random number sequence for 120 participants. Allocations of fibromyalgia patients in the groups were done using the ‘Opaque Concealed Envelope’ method. Study was blinded to principal investigator, outcome assessor and data analysts, while it is non-blinded to both participants and therapists.

### Recruitment Criteria

Both male and female fibromyalgia patients aged 21-50 years, diagnosed according to American College Rheumatology (ACR) 2010/2016 criteria having pain rating ≥ 5 on Visual Analogue Scale (VAS) for at least 3 months were recruited from the Rheumatology and Physical Medicine and Rehabilitation clinics, majorly. Presence of any major illness (autoimmune, cardiovascular and neuropsychiatric disorders which is not associated with fibromyalgia); contraindications to TMS which includes metallic implants, facial tattoos, skull defects, pacemakers, heart valves, neuroactive drugs, history of seizures, major head trauma in past six months; history of opioid or substance abuse and pregnant or lactating women were not recruited in the study. [29] Patients with chronic pain other than fibromyalgia were excluded from the study. Age and gender matched pain-free healthy participants were recruited as a part of the case control study. Exclusion criteria for the healthy participants were the same as fibromyalgia patients.

### Subjective assessment of pain and associated symptoms of fibromyalgia using questionnaires

Pain and associated symptoms of fibromyalgia was subjectively assessed using Visual Analogue Scale (VAS), Tender Point Counts (TPCs), Short Form McGill Pain Questionnaire (SF-MPQ), Pain Catastrophizing Scale (PCS), Fibromyalgia Impact Questionnaire (FIQ) and Pittsburgh Sleep Quality Index (PSQI). Questionnaires were handed over to the patient in the vernacular language (in which they were comfortable) after asking pain scores on numerical pain rating scale (VAS). Global scores for each questionnaire were calculated and reported either domain-wise or net outcome measures of pain and associated symptoms like sleep, fibromyalgia impact and quality of life.

### Assessing pressure pain parameters using Quantitative Sensory Testing

Quantitative Sensory Testing (QST) is help to assess and quantify sensory function in patients with pain and some neurological disorders and helps in detecting sensory loss or hyperalgesia, allodynia and tenderness. Figure 1a is the schematic representation of QST performed on the participants’ reference and test sites. QST was done using the method of limits for two parameters (Pressure Pain Threshold and Pressure Pain Tolerance) at four different sites (dorsum of right hand as reference site; left and right shoulder; and lower back as test site). Pressure pain threshold was defined as the minimum pressure applied on the test site to elicit first/minimum pain sensation equivalent to VAS score of 1. Pressure pain tolerance was the maximum tolerable pressure to generate sensation of pain which is equivalent to VAS score of 5. The tests were first performed at reference site and then at the test site. Pressure pain parameters can be assessed using mechanosensory force exerted per unit area at the dermatomes of the test site by holding the ergonomic handle of the AlgoMed device and exerting pressure at a constant rate. Mechanical stimuli per se pressure modality can be regulated by examiner and the subject both; former can stop the test by removing the algometer from the test site whereas the subject can respond using the ‘response unit’ held in the hand. The pressure is increased slowly and uniformly at the test site till the pain threshold or the pain tolerance threshold at the test site is elicited in fibromyalgia patients or healthy subjects. The QST protocol of the German Research Network on Neuropathic Pain was utilized. ‘Medoc Main Station’ software was used for recording and the methods of limits were employed for assessment of both threshold and tolerance. Algometer was placed at the right angle to the dorsal side of the first dorsal interosseous of the dominant hand (reference site). The dorsal scapular surfaces (shoulder blade) and L4-L5, identified manually; all tests were performed 5 cm anterior and lateral to these sites respectively. Supplementary Figure 1 demonstrates the actual set-up for the recording.

**Figure 1:**
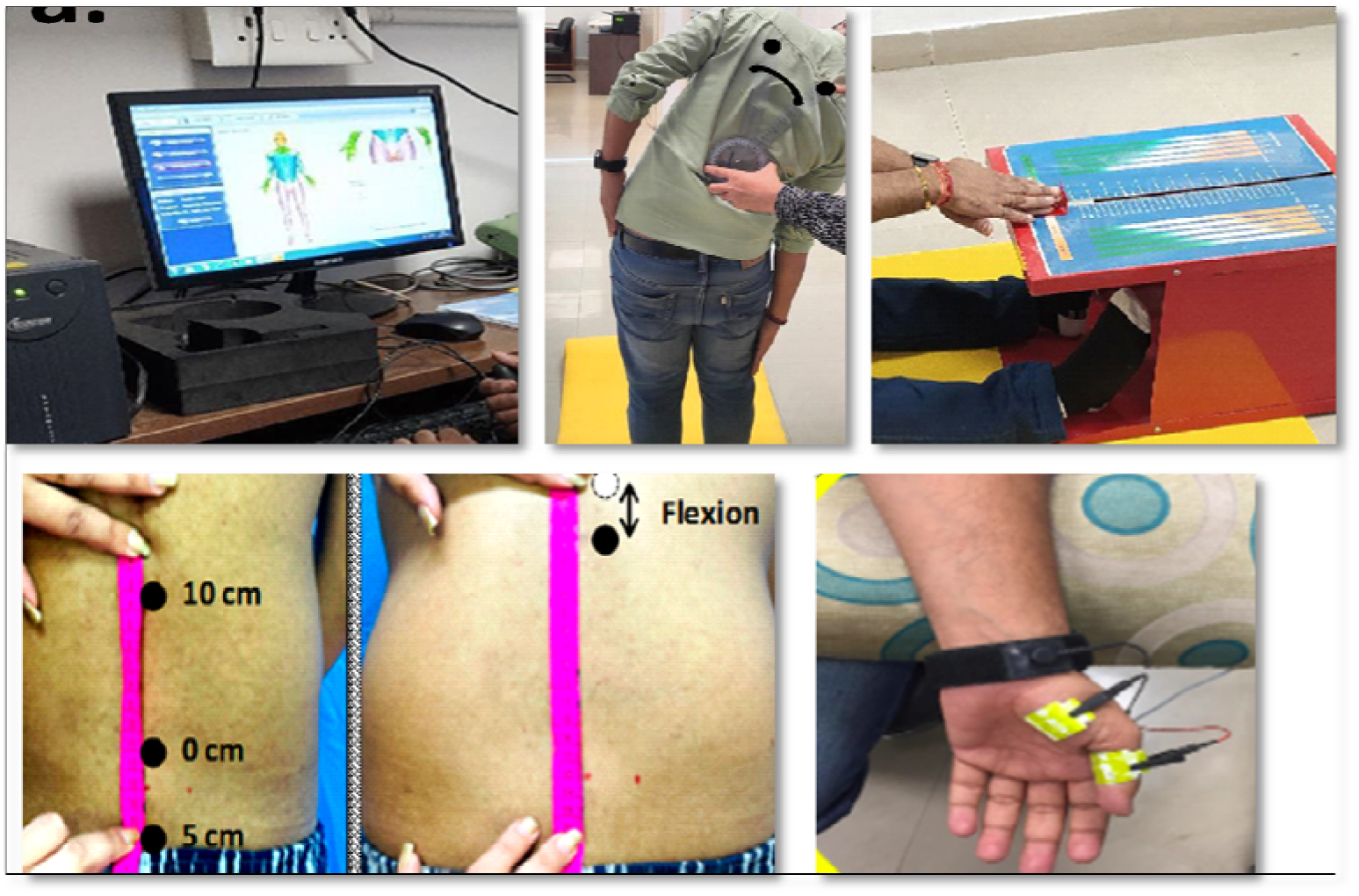
Materials and methods: quantitative sesnory testing, lateral goniometry, modified Schober’s test, sit and reach test, and electrode placement in recording of corticomotor excitability for the left abductor pollicis brevis muscle and ground electrode at wrist (left to right).

### Flexibility and range of motion assessment

Flexibility and range of motion was recorded in the fibromyalgia patients and age and gender matched healthy controls to assess the musculoskeletal performance before and after the yogic intervention. Three objective tests – Modified Schober’s Test, Lateral Goniometry and Sit and Reach Test, were deployed early morning in all the participants to avoid possible biases.

### Goniometry

Assessment of range of motion was performed by placing the central axis of the goniometer (a tool that measures the range of motion of various joints around the normal body axis) at the axis of rotation around L5. The arms of the goniometer are aligned with the vertebral axis of the subject positioned first at 0° angle between the mobile and fixed arm. Goniometer is stably fixed and positioned at the midpoint of the vertebral/spinal axis in the anterio-posterior direction at L5. Free arm was rotated according the lateral bending movement of the body and upto the the extent of individual’s maximum flexion. The angle measurement was recorded thrice and averaged (Figure 1b).

### Modified Schober’s test

The Modified Schober’s test was utilized for measuring lumbar flexibility and associated range of motion along spinal axis obtained through radiographs. It was first described by Van Adrichen and Van der Korst (1973). A reference point was marked at L5 and patients were asked to bend forward with their hands and face in relaxed position. [30] Displacement an arbitrary point drawn 15 cm anterior to the reference point were recorded thrice and averaged, in the flexed position. (Figure 1c)

### Sit and Reach Test

Sit and Reach test was used to assess lumbar flexibility in fibromyalgia patients undergoing rehabilitation intervention. It involves sitting the participant on the floor with legs stretched out without shoes and soles placed flat against the sit and reach box (American College of Sports Medicine’s Sit and Reach Test box). Slider of the sit and reach box was moved with the joined fingers of both the hands until patient felt uncomfortable, and the final position of the slider was noted; this process was repeated thrice and values were averaged. (Figure 1d)

### Recording of cortical excitability using Transcranial Magnetic Stimulation

Cortical excitability is defined as the strength of cortical outputs in response to an external magnetic/electromagnetic stimulus, which can be assessed by a Transcranial Magnetic Stimulation (TMS) machine (Figure 1e). Parameters recorded were Resting Motor Threshold (RMT), Motor Evoked Potential (MEP), Cortical Silent Period (CSP), MEP AND CSP Recruitment Curves. Methods for assessing corticomotor excitability using TMS are described in accordance with the latest guidelines on its methodology for reporting. [31] Pulses were delivered with a figure-of-eight transcranial magnetic stimulation coil (70 mm; Neuro-MS/D, Ivanova, Russia) at an angle of 45° to the sagittal plane of the scalp to generate antero-posterior waves inside the brain. Inter-stimulus intervals were randomized between 5 to 10 seconds depending on the previous stimulus. The acquired signals were amplified (1000x), bandpass filtered between 10 Hz to 2 kHz (Neuro-MEP-Micro, Ivanovo, Russia) and analysed offline (Neurosoft.EP software, Ivanovo, Russia).

### Resting motor threshold

It is the minimum stimulus intensity that can elicit a motor evoked potential with peak-to-peak amplitude of at least 50 µV strength in a minimum of five out of ten consecutive trials (50%) with the target muscle at rest [32]. A visible twitch can be obtained at the abductor pollicis brevis (ABP) muscle with recording of electromyogram by stimulating respective homunculus in the M1.

### Motor Evoked potential

The electromyography sampling window consisted of 100 ms of pre- stimulus, followed by a 200 ms of post-stimulus which can be modified according to the EMG output. Evoked potential amplitude was measured as the voltage difference between the maximum and minimum aspects of the waveform. The evoked potential amplitude of ten trials obtained at same stimulation intensity was then averaged.

### Motor evoked potential Recruitment Curve (MEP-RC)

The stimulation intensities for obtaining MEP- RC were based on individual’s Resting Motor threshold. Five stimuli at each intensity (% MSO) were stepped-up from 90% to 150% of the resting motor threshold at an increment of 10%. Thus, a total of thirty-five pulses were delivered. To construct the motor recruitment curve, the mean amplitude of evoked potential was plotted against level of stimulation intensity.

### Cortical Silent period

Participants were instructed to hold APB muscle at 30% of maximum voluntary contraction (MVC) using audio-visual cues of ongoing surface EMG recording. Silent period was measured as time difference from onset of evoked potential waveform to resumption of ongoing electromyographic activity. The silent period duration of ten trials at each stimulus intensity were averaged.

### Cortical Silent Period Recruitment curve

like the paradigm of recording MEP recruitment curve, the stimuli were given at the hotspot while the patient was asked to maintain MVC at 30%. Five stimuli were given at 90-150% of RMT at an increment of 10% in each step; the mean duration was plotted against level of stimulation intensity.

### Enzyme Linked Immunosorbent Assay

After collecting and processing the blood for serum isolation, levels of pain and associated biomarkers were estimated using Enzyme Linked Immuno-sorbent Assay (ELISA) in the laboratory settings in 50 μL standards and pilot samples in duplicates to decrease experimental errors. Biotin-labeled antibody and HRP-Streptavidin Conjugates specific to our markers were utilized. TMB Substrate was added into each well and static incubation at 37°C in dark was performed for the given time. Followed by addition of 50 μL stop solution and optical density measurement at 450 nm immediately to calculate concentration of the biomarkers (Supplemetary data X).

### Yogic Intervention Protocol

Protocol for delivering supervised *Hatha yoga* was validated by the panel of qualified medical yoga therapists and deployed for five days a week for four weeks at Integral Health and Wellness Clinic, Department of Physiology of the same tertiary care hospital. It was a comprehensive yoga therapy-based intervention program lasting for 1.5 hours per day in the morning in comfortable outfit. It consisted of an integrated and pretested intervention which consisted of practice sessions along with demonstration and rectification together. [27, 33] To ensure the quality of the program and to ensure that the participants get enough time with the expert, a maximum of 6 participants were enrolled in the program for the specified regime after proper instructions and monitoring of blood pressure and heart rate, which helped us to have good supervision on every patient. We have not included any participants in between the program; all the participants of a batch were trained together, starting and finishing together.

A typical day of the yoga session started with a set of loosening exercises (Sukshma Vyayama) depicted in Figure 2; then, simple physical postures (Asanas), relaxing exercises and finally the breathing exercises (Pranayama) for approximately one hour. Asanas consisted of supine, prone, standing, and sitting postures. All asanas and other yoga practices were supervised by expert yoga therapists and counsellors throughout the yoga regimen (Table 1 & Figure 3). Nutritional awareness programs along with counselling sessions were included for the next half an hour in which questions and unstructured group discussions and motivations were provided amongst the patients. Relaxation through Shavasana (a relaxation technique), meditation and Yog-nidra were also planned at the end of each session. Spouse and other members from the patient’s family were also encouraged to facilitate compliance. Patients were encouraged to practice a learned yoga regimen at home for at least 3 months along with the Sukshmavyayama (Figure 2-3). Patients were advised to take a healthy diet regimen during the yoga program – ‘Saatvik Aahaar’ which primarily includes green and leafy vegetables and fruits during the intervention which has high protein, high fibers, low carbohydrates, and low-fat diets. Each session of yoga therapy (5 sessions/week for 4 consecutive weeks; a total of 20 sessions) was divided into two parts: a yoga session and a counselling cum discussion session. We have recommended not stopping the ongoing medications. Any of the patients who failed to attend 90% of the sessions were not included in the study but they certainly went through the therapeutic benefits. Some of the actual photographs from our yoga centre in given in Figure 4.

**Figure 2:**
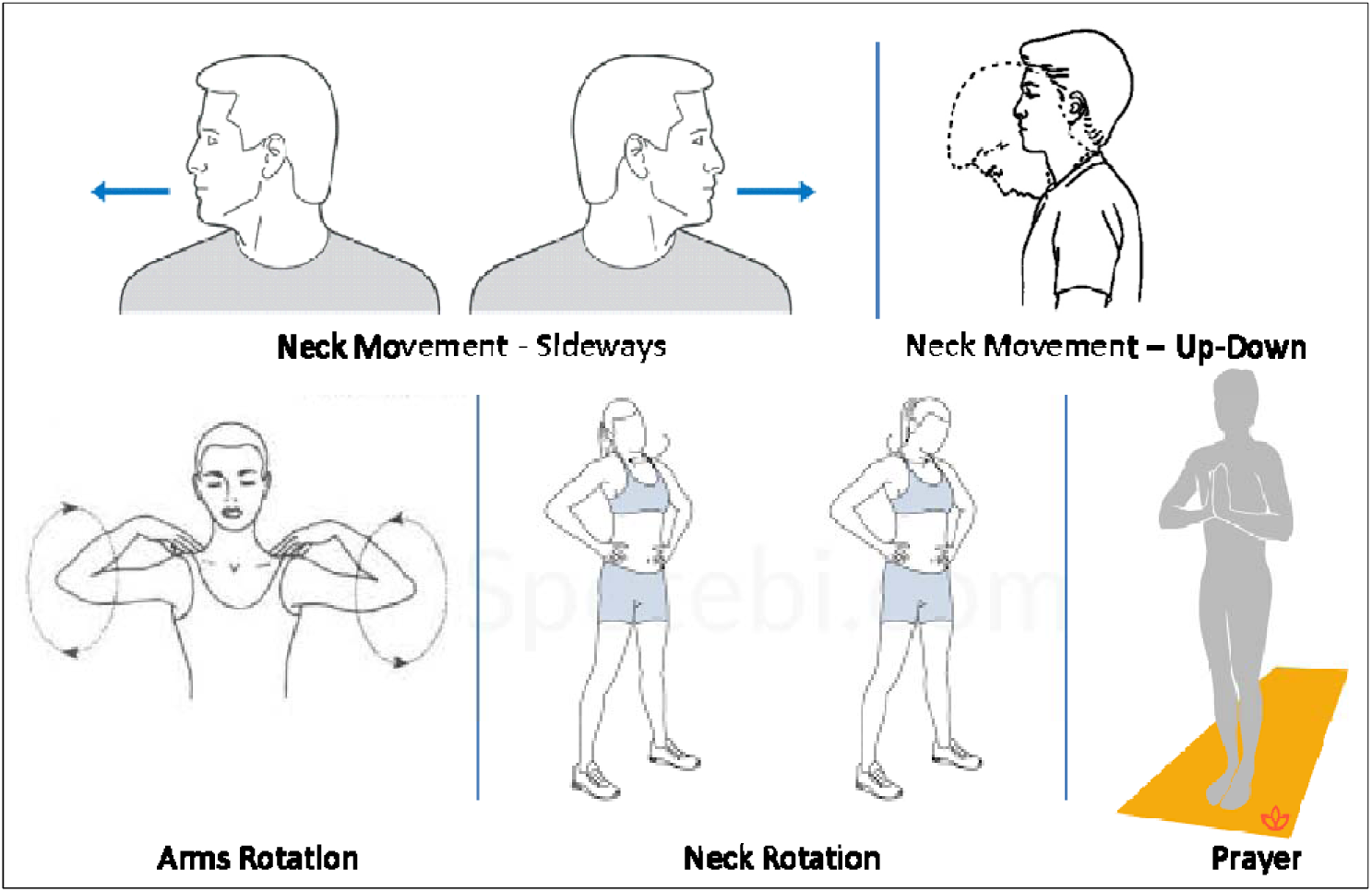
Sukshmavyayama (loosening exercises) performed by the fibromyalgia patients in the Integral Health and Wellness Clinic, Department of Physiology, AIIMS, New Delhi.

**Figure 3:**
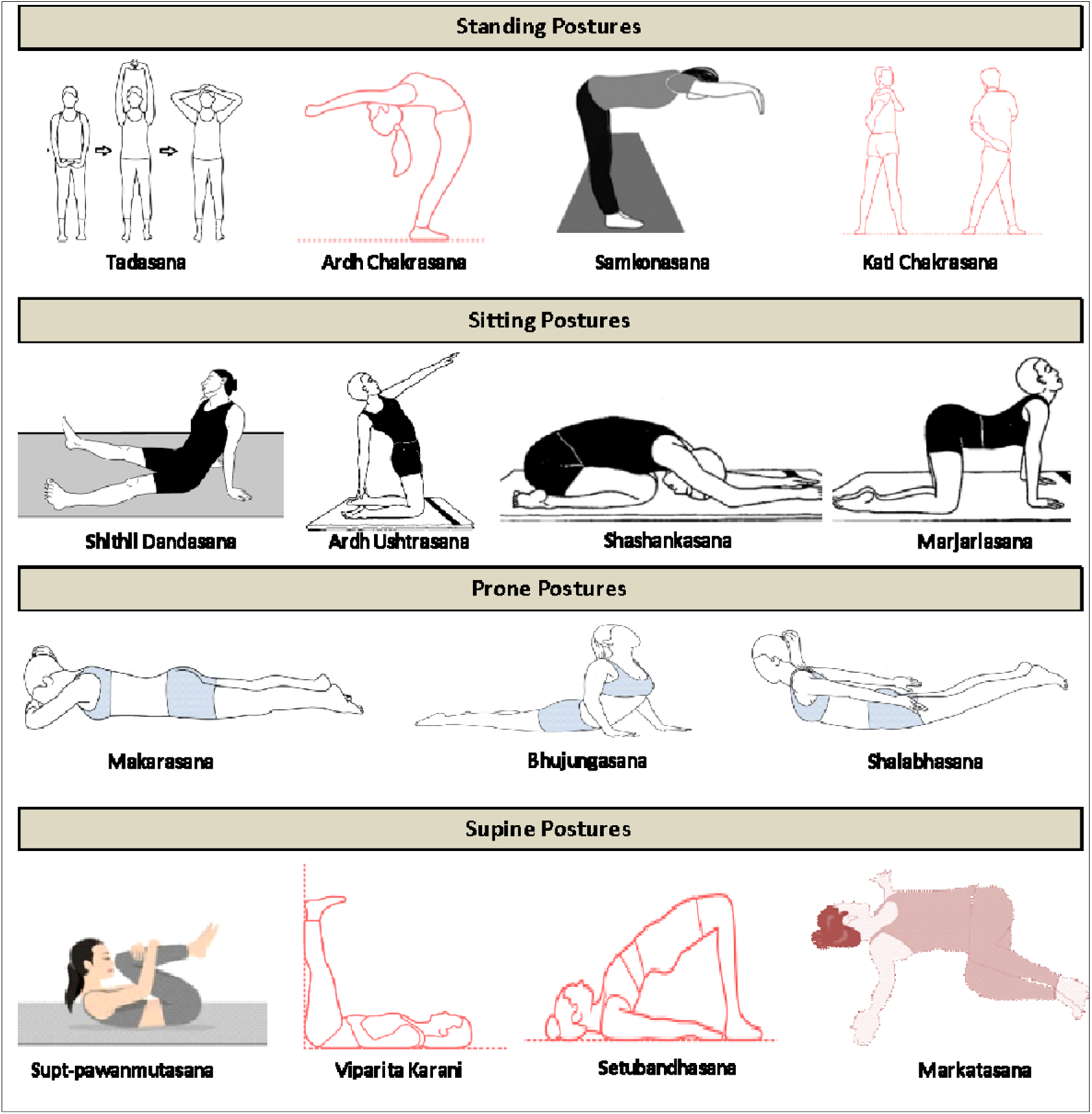
Schematic representation of yoga asanas performed by fibromyalgia patients in the Integral Health and Wellness Clinic, Department of Physiology, AIIMS, New Delhi.

**Figure 4:**
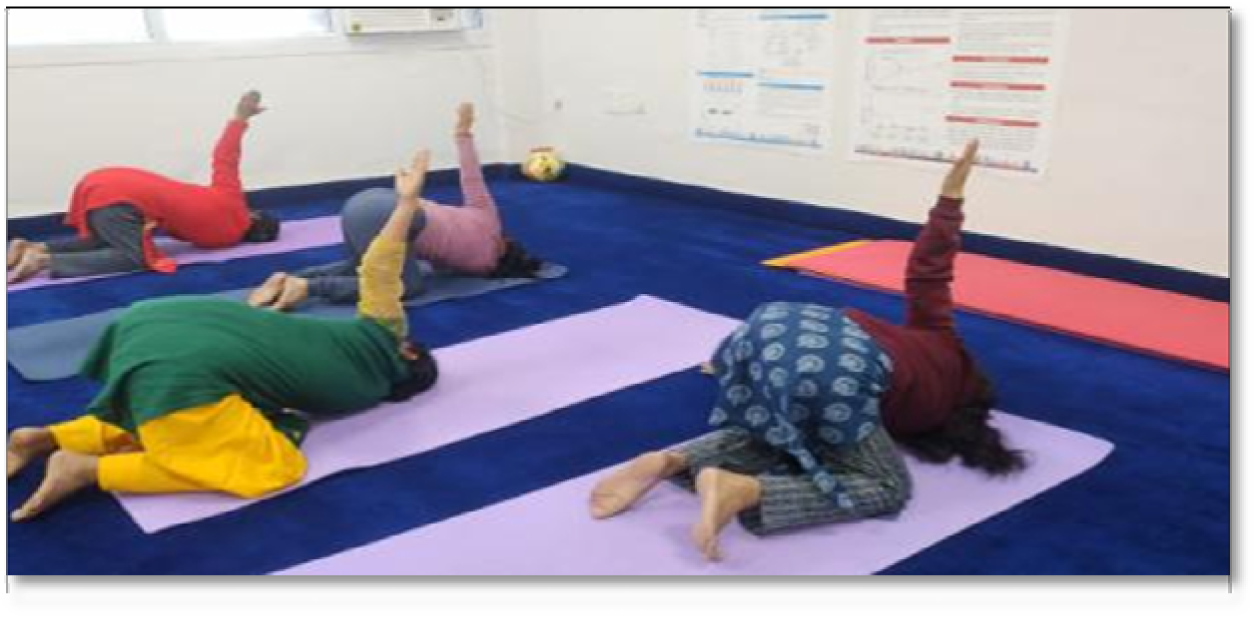
Representative photograph of supervised yogic intervention performed by the fibromyalgia patients in the Integral Health and Wellness Clinic

**Table 1:** Yogic Intervention Protocol for Fibromyalgia Patients Stndrdised in the Pain Research and TMS Labortory.

| S. No. | Yoga Practices | Frequency/duration |
| --- | --- | --- |
| 1. | **Prayer** | 2 minutes |
| 2. | **Loosening exercises/Sukshma Vyayama**<br>Griva Shakti Vikasaka/Neck Movement (Rotation – clockwise and anti-clockwise; up and down; sideways)<br>Bhujia Valli Shakti Vikasaka Purna Bhujia Shakti Vikasaka/Shoulder Movement<br>(Rotation clockwise-anti-clockwise, Elbows fold in-out, Rotate arms alternatively)<br>Kati Shakti Vikasaka/Trunk Movement (Twist body along waist, Side bending)<br>Jangha Shakti Vikasaka (II-A&B)/ Thigh Movement (chair pose, kick with alternate leg)<br>Janu Shakti Yikasaka/ Knee movement (Rotate knees clockwise, anti-clockwise)<br>Pada-mula shakti Vikasaka/Ankle movement (Toes pointing outwards-up, down, sideways)<br>Gulpha-pada-pristha-pada-tala shakti Vikasaka/Ankle movement (Rotate ankle clockwise and anti-clockwise) | 3 times each |
| 3. | **Standing Asanas**<br>Tadasana<br>Ardha Chakrasana<br>Samkonasana<br>Katichakrasana | 3 times each with regulated breathing |
| 4. | **Sitting Asanas**<br>Shithildandasana<br>Ardhushtasana<br>Shashankasana<br>Marjariasana | 3 times each with regulated breathing |
| 5. | **Prone Asanas**<br>Makarasana<br>Bhujangasana<br>Shalabhasana | 3 times each with regulated breathing` |
| 6. | **Supine Asanas**<br>Supt-pawanmuktasana<br>Viparitarani<br>Setubandhasana<br>Markatasana | 3 times each with regulated breathing |
| 7. | **Pranayam**<br>Bhramari<br>Anulom vilom | 10 minutes |
| 8. | **Relaxation technique/Shavasana**<br>Voluntarily relaxing each part of body by focussing attention (toes to the head) | 5 minutes |
| 9. | **Prayer** | 2 minutes |

### Data processing and statistical analysis

Data was stored in the Microsoft Excel Sheet. Data analysis was performed using Stata 13 software tool. Graphs were plotted with MedCalc, GraphPad Prism and MS Excel software Inc. Gaussian distribution (normality test) for the data was performed using Shapiro-Wilk test. Parametric data was expressed in Mean ± SEM while non parametric in Median (Q1, Q3). Intra-group comparisons for the efficacy of interventions were performed using ‘Paired t-test’ and ‘Mann-Whitney U test’ for parametric and non-parametric data respectively. Parametric data was expressed in Mean ± SEM while non parametric in Median (Q1, Q3). Intergroup comparison between post therapy time-points were performed using ‘Two sample independent t-test’ and ‘Wilcoxon signed rank sum (Mann-Whitney U) test’ for parametric and non-parametric data respectively. Adjustment for the baseline significant data was done using ANCoVA while comparison for post intervention time-points. Significance level for all the results was set at 0.05. Probabilities below 5% were considered statistically significant. The sample size was determined with power calculation formula using MedCalc Software tool based on a comparable case-control study by Williams et al., 2009, anticipating minimally clinical relevant difference (MCRD) of 1 in the VAS score between the groups, with combined standard deviation of 3.2 units; the estimated sample size was calculated as 99 participants. [34] The assumed power and level of significance were 80% and 5% respectively and assuming 20% loss-to-follow up, the study thus included 120 chronic low back pain patients. Ethical clearance for 180 participants (60 each group) were taken and achieved the committed sample size for biomarkers quantification.

## Results

Result section comprises of 1. Results of Case Control Study (n = 120); 2. Results of Interventional Study (n = 60), and 3. Results of Randomized Controlled Trials (n = 120). Data was primarily collected for the randomized controlled trial (RCT) in 120 fibromyalgia patients undergoing either yogic intervention or standard therapy recommended by the medical specialists. Consort flow chart for the study is given in Figure 5.

**Figure 5:**
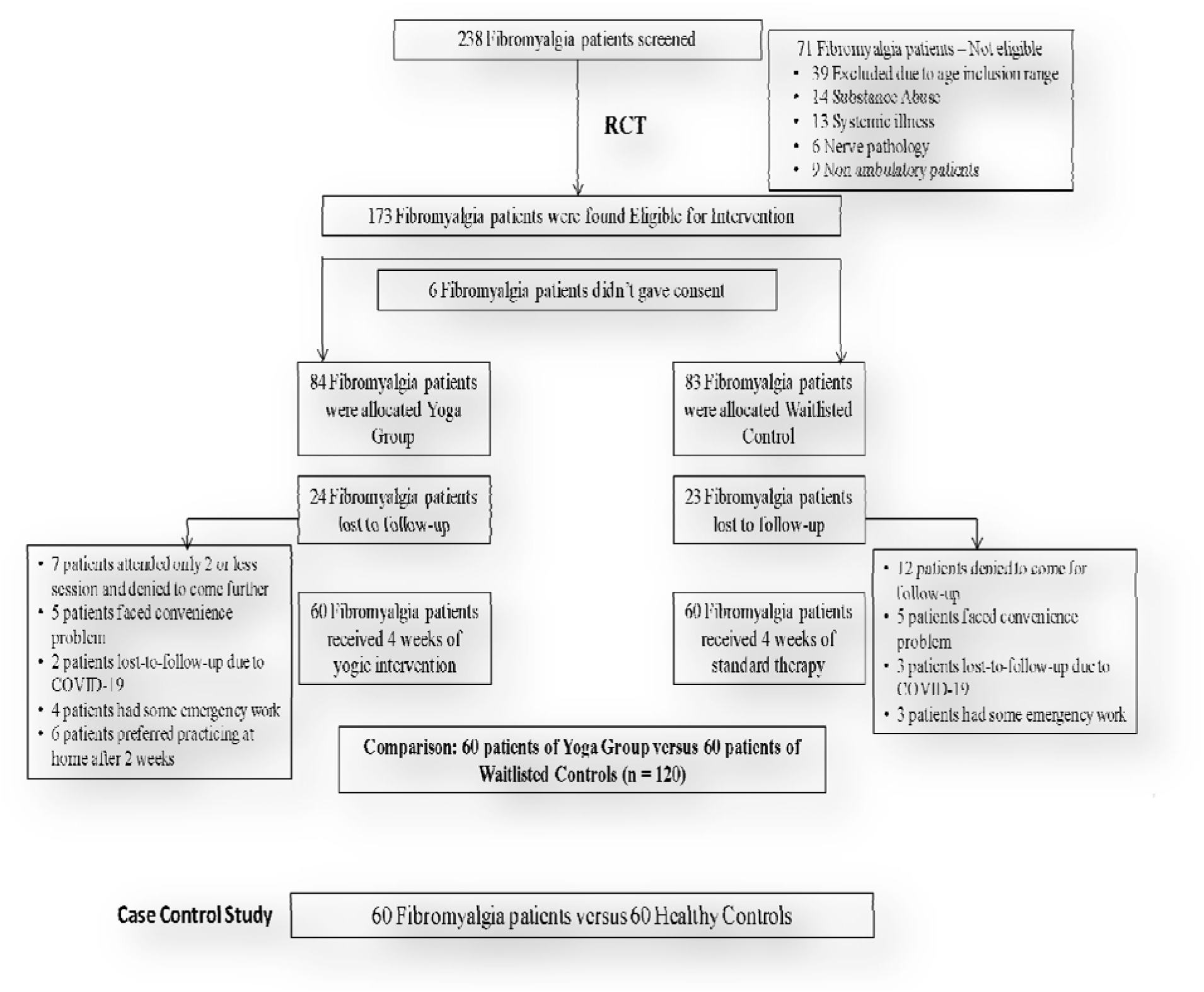
Consort flow chart for the study.

### Results of the Case Control Study (n = 120)

Primary objective was to compare the measures of pain status, quality of life and sleep, flexibility and range of motion, cortical excitability, and levels of blood biomarkers (β-Endorphin, Cortisol, Glutamate, Serotonin and Substance P) in 60 fibromyalgia patients with age and gender matched healthy controls.

### Physiological characteristics, quality of life and sleep quality of fibromyalgia patients (n = 60) and healthy controls (n = 60)

Average weight and BMI of the fibromyalgia patients (FM) were 63.04 ± 1.34 Kg and 25.70 ± 0.51 Kg/m^2^ respectively; which were not significantly different from that of healthy controls 65.96 ± 1.63 Kg and 25.67 ± 0.52 Kg/m^2^ (p-value = 0.166 and 0.976 respectively) at baseline. Heart rate of the fibromyalgia patients on an average was 83.97 ± 1.79 beats/minute and that of healthy controls was 80.25 ± 1.39 beats/minute; which was non-significantly different (p-value = 0.068), although they tend reach the significance level. Systolic and diastolic blood pressure of healthy controls were found significantly lower than that of fibromyalgia patients (Systolic: p-value = 0.0002 and Diastolic: p-value <0.0001) represented in Table 2 and Figure 6a-b.

**Figure 6.**
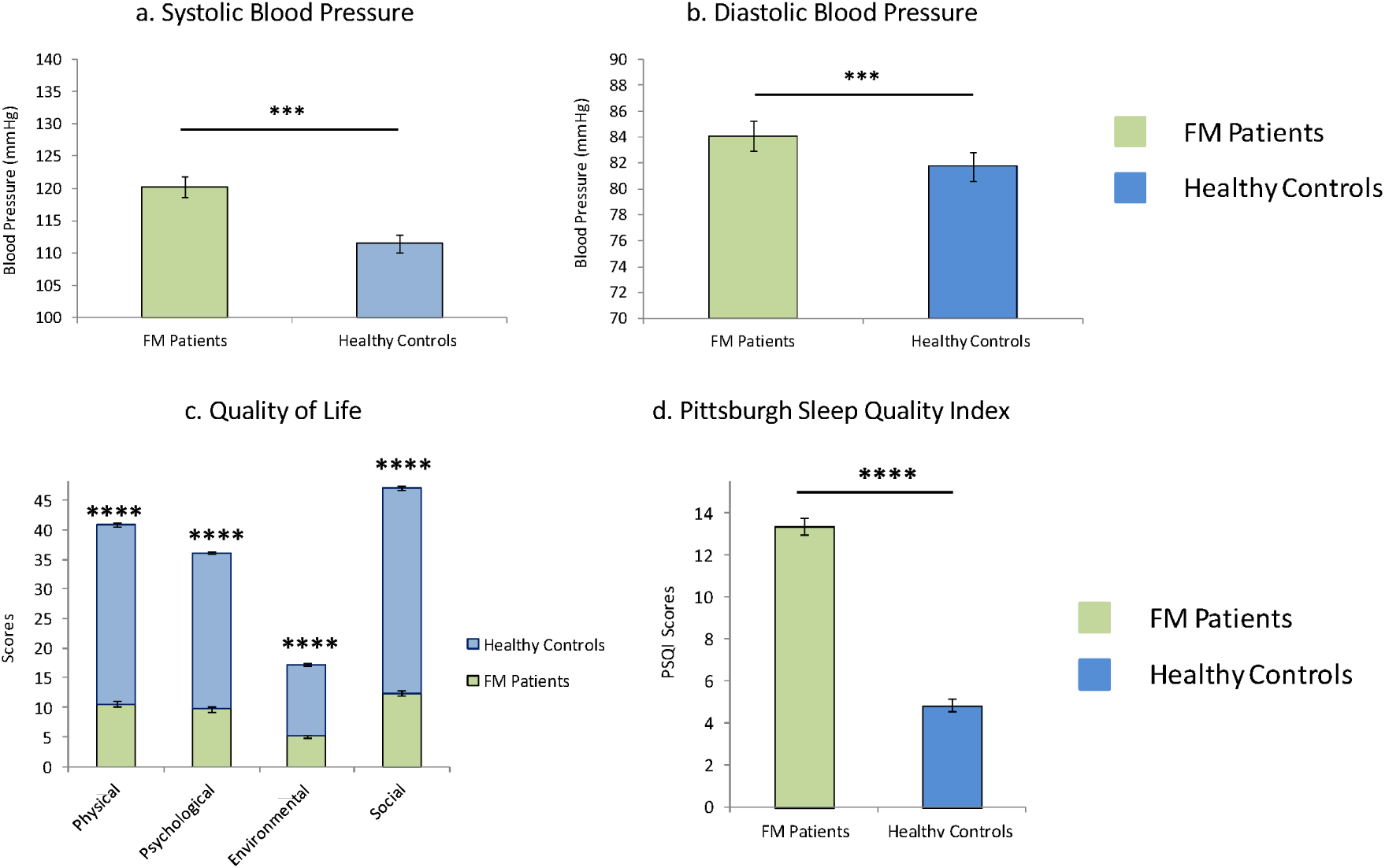
Physiological characteristics, quality of life and sleep quality of fibromyalgia patients and healthy controls: a) Systolic Blood pressure, b) Diastolic Blood Pressure, c) WHO-Quality of Life and d) Sleep Quality Index. Data was normally distributed (Gaussian distribution) – ‘Shapiro-Wilk test’; expressed in Mean ± SEM. Two sample (Independent) t-test was performed for comparison between fibromyalgia patients and healthy controls. p-value < 0.05 was considered significant. Asterisk (*) depicts a significant difference between the arms. “****” represents p<0.0001. WHOQOL – World Health Organisation Quality of Life; PSQI – Pittsburgh Sleep Quality Index.

**Table 2:**
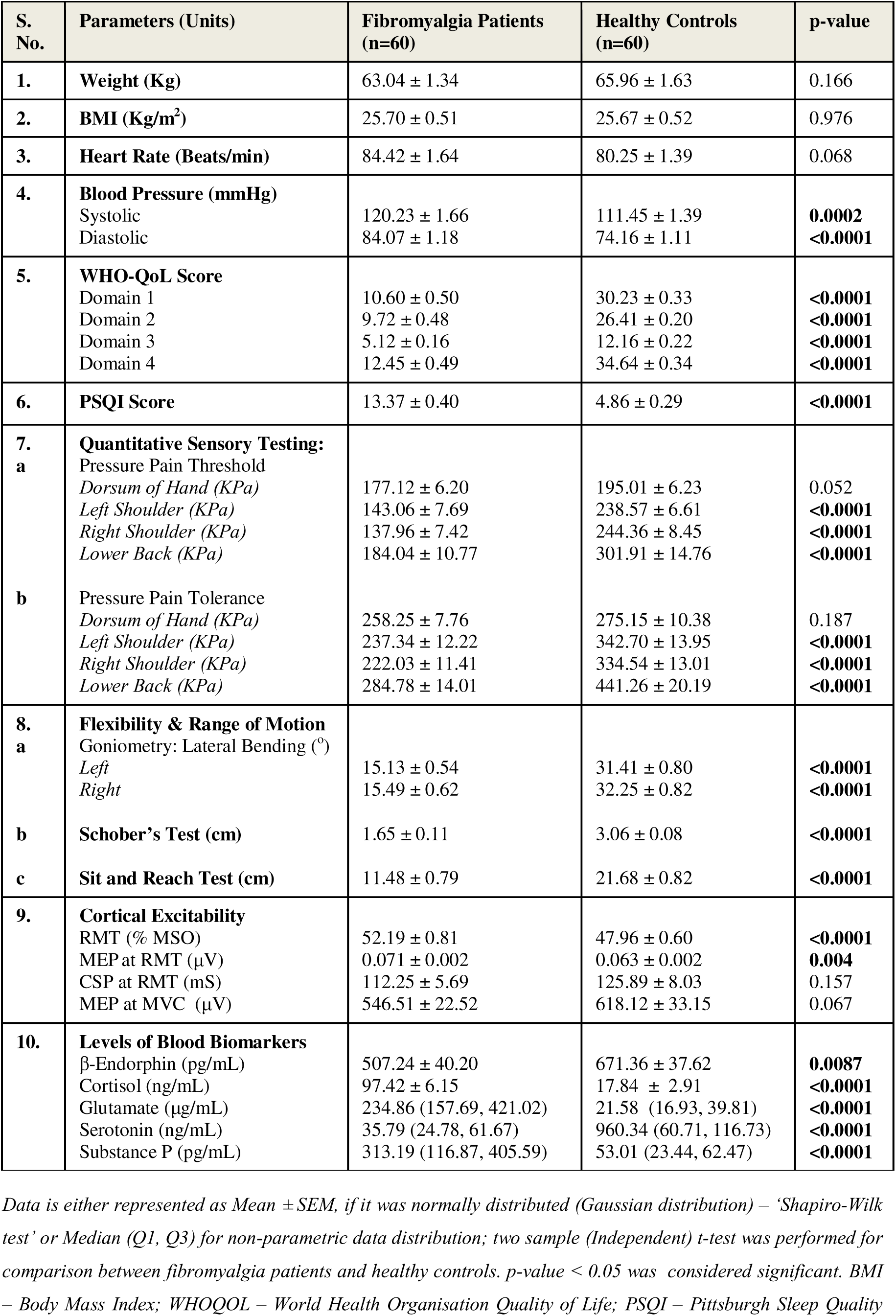
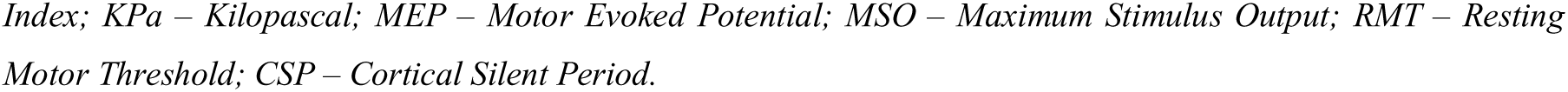
Comparison of study parameters of fibromyalgia patients and healthy controls in the case-control study.

Quality of life of fibromyalgia patients (FM) and healthy controls when assessed using WHO-QOL-BREF questionnaire for all the four domains: physical, psychological, environmental and social; a significant retardation in it was reported in fibromyalgia patients as compared to healthy cohort in all domains – physical domain (Fibromyalgia patients: 10.60 ± 0.50 and Healthy Controls: 30.23 ± 0.33; p<0.0001), psychological domain (Fibromyalgia patients: 9.72 ± 0.48 and Healthy Controls: 26.41 ± 0.20; p<0.0001), environmental domain (Fibromyalgia patients: 5.12 ± 0.16 and Healthy Controls: 12.16 ± 0.22; p<0.0001), and social domain (Fibromyalgia patients: 12.45 ± 0.49 and Healthy Controls: 34.64 ± 0.34; p<0.0001). (Table 2 & Figure 6c). PSQI measures sleep impairment, if any, in subjects suffering from disturbed sleep due to psychological distress or chronic pain. Sleep quality in fibromyalgia patients was found significantly poorer than the healthy controls of our study (Fibromyalgia patients: 13.37 ± 0.40 and Healthy Controls: 4.86 ± 0.29; p<0.0001) (Table 2 & Figure 6d).

### Sensory Testing of Fibromyalgia Patients (n = 59) and Healthy Controls (n = 59)

Objective assessment of pain was done using Quantitative Sensory Testing (QST) machine with the pressure modality by the digital algometer around three localized tender points (left shoulder, right shoulder and lower back) of fibromyalgia and one reference site (dorsum of hand). Both the pressure pain threshold and pressure pain tolerance were higher at the reference site as compared to the left and right shoulders at baseline and post-intervention. Pressure pain threshold at dorsum of hand (Fibromyalgia patients = 177.12 ± 6.20 kPa and Healthy Controls = 195.01 ± 6.23 kPa; p = 0.052), left shoulder (Fibromyalgia patients = 143.06 ± 7.69 kPa and Healthy Controls = 238.57 ± 6.61 kPa; p<0.0001), right shoulder (Fibromyalgia patients = 137.96 ± 7.42 kPa and Healthy Controls = 244.36 ± 8.45 kPa; p<0.0001) and lower back (Fibromyalgia patients = 184.04 ± 10.77 kPa and Healthy Controls = 301.91 ± 14.76 kPa; p p<0.0001) of healthy controls were significantly higher than that of the fibromyalgia patients (Table 2 & Figure 7a). Though pressure pain thresholds of fibromyalgia patients and healthy controls at dorsum of hand were not significantly different, but it has been achieving the set significance level. Moreover, pressure pain tolerance, except for the dorsum of hand (Fibromyalgia patients = 258.25 ± 7.76 kPa and Healthy Controls = 275.15 ± 10.38 kPa; p = 0.187) was found significantly higher at left shoulder (Fibromyalgia patients = 237.34 ± 12.22 kPa and Healthy Controls = 342.70 ± 13.95 kPa; p<0.0001), right shoulder (Fibromyalgia patients = 222.03 ± 11.41 kPa and Healthy Controls = 334.54 ± 13.01 kPa; p<0.0001) and lower back (Fibromyalgia patients = 284.78 ± 14.01 kPa and Healthy Controls = 441.26 ± 20.19 kPa; p<0.0001) of healthy controls as compared to the fibromyalgia patients of our cohort (Table 2 & Figure 7b).

**Figure 7.**
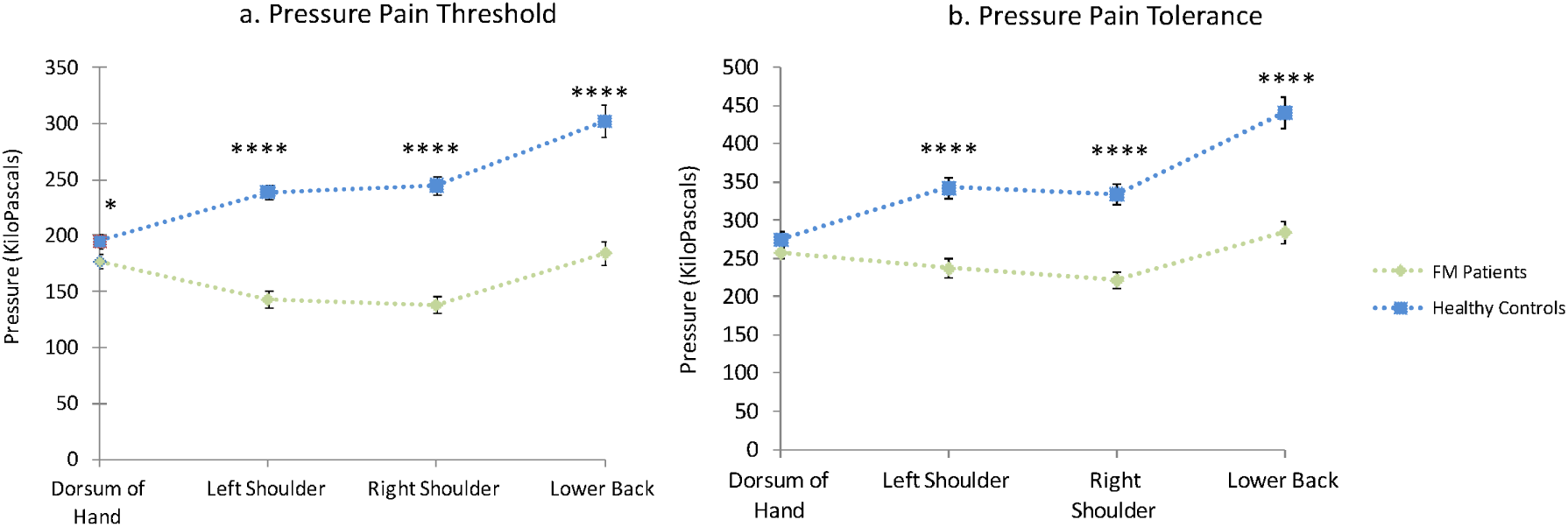
Assessment of pressure pain parameters using quantitative sensory testing (QST) at dorsum of hand, shoulders and lower back of fibromyalgia patients and healthy controls. a) Pressure pain threshold and b) Pressure pain tolerance were measured at the same site in continuation. Data was normally distributed (Gaussian distribution) – ‘Shapiro-Wilk test’; expressed in Mean ± SEM. Two sample (Independent) t-test was performed for comparison between fibromyalgia patients and healthy controls. p-value < 0.05 was considered significant. Asterisk (*) depicts a significant difference between the arms. “****” represents p<0.0001.

### Flexibility and range of motion of fibromyalgia patients (n = 60) and healthy controls (n = 60)

A significantly greater flexibility and range of motion was reported in healthy controls than that of fibromyalgia patients in both the left (Fibromyalgia Patients = 15.13 ± 0.54 and Healthy Controls = 31.41 ± 0.80 ; p<0.0001) and right (Fibromyalgia Patients = 15.49 ± 0.62 and Healthy Controls = 32.25 ± 0.82 ; p<0.0001) directions (Table 2 & Figure 8a). Moreover, lumbar flexibility was found significantly better in healthy controls than the fibromyalgia patients of same age group (Fibromyalgia Patients = 11.48 ± 0.79 cm and Healthy Controls = 21.68 ± 0.82 cm; p<0.0001) when assessed using Sit and Reach box (Table 2 & Figure 8b). Lumbar flexion and range of motion in patients with fibromyalgia and age and gender-matched healthy individuals were assessed using modified Schober’s test, Lateral Goniometry and Sit and Reach box. We found a significantly higher flexibility and range of motion in healthy controls as compared to fibromyalgia patients of our cohort on assessment using Modified Schober’s test (Fibromyalgia Patients = 1.65 ± 0.11 cm and Healthy Controls = 3.06 ± 0.08 cm; p<0.0001) (Table 2 & Figure 8c).

**Figure 8.**
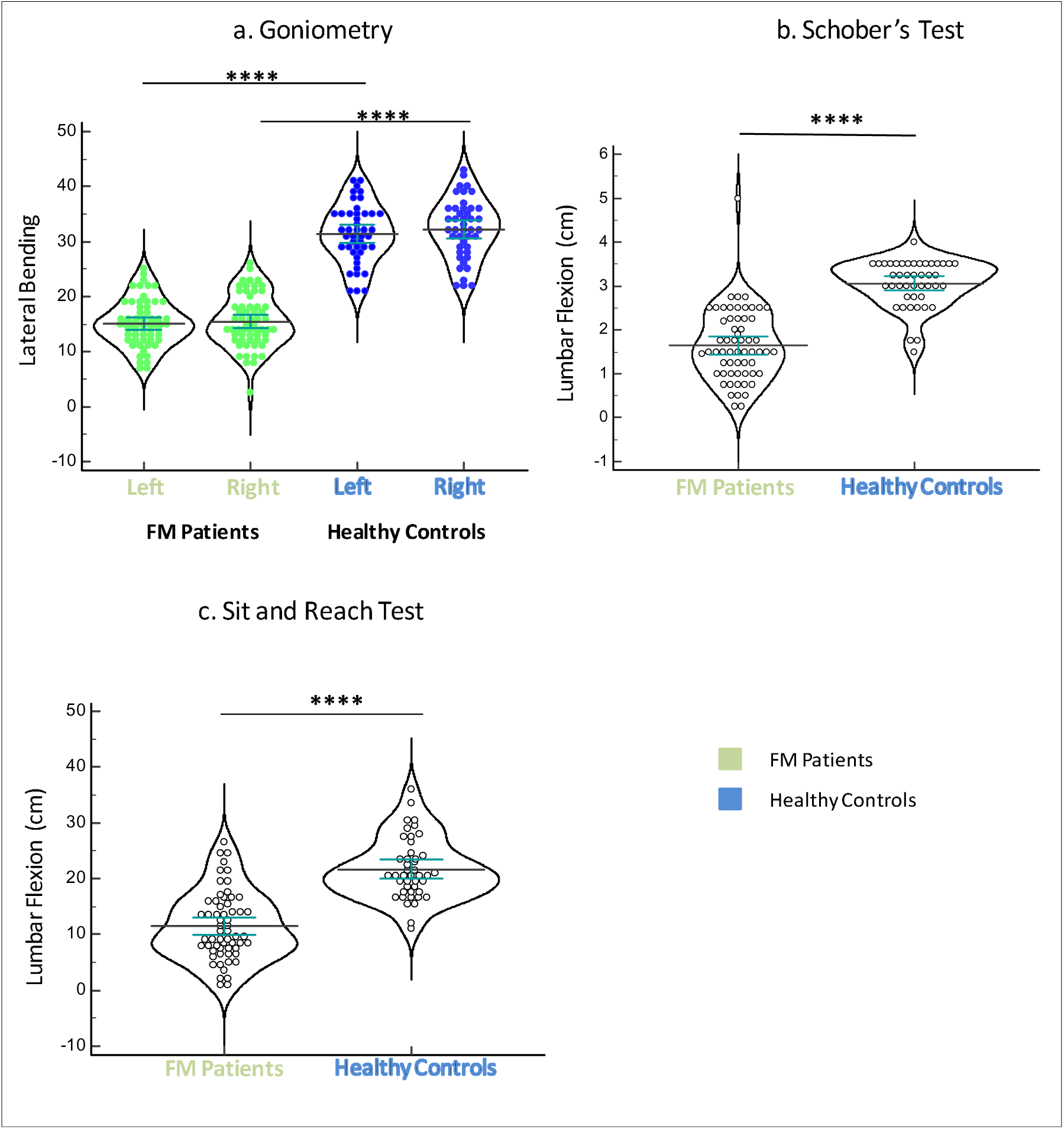
Comparison of pre-yoga and post-yoga flexibility and range of motion in fibromyalgia patients: a) Goniometry; b) Schober’s Test and, c) Sit and Reach test. Data was normally distributed (Gaussian distribution) – ‘Shapiro-Wilk test’; expressed in Mean ± SEM. Two sample (Independent) t-test was performed for comparison between fibromyalgia patients and healthy controls. p-value < 0.05 was considered significant. Asterisk (*) depicts significant difference between the arms. “****” represents p<0.0001.

### Cortical excitability of fibromyalgia patients (n = 60) and healthy controls (n = 60)

Cortical excitability of 60 fibromyalgia patients and 60 healthy controls were measured using Transcranial Magnetic Stimulation (TMS) at primary motor cortex (M1). Resting Motor Threshold (RMT) of fibromyalgia patients was found significantly higher (Fibromyalgia Patients = 52.19 ± 0.81 % MSO and Healthy Controls = 47.96 ± 0.60 % MSO; p<0.0001). We have also found a significantly lower Motor Evoked Potential (MEP) at RMT (Fibromyalgia Patients = 0.071 ± 0.002 mV and Healthy Controls = 0.063 ± 0.002 mV; p = 0.004) in healthy controls as compared to fibromyalgia patients. Cortical Silent Period (CSP) of the fibromyalgia patients was not different from those of the healthy controls (Fibromyalgia Patients = 112.25 ± 5.69 ms and Healthy Controls = 125.89 ± 8.03 ms; p = 0.157). (Table 2 & Figure 9a-b)

**Figure 9.**
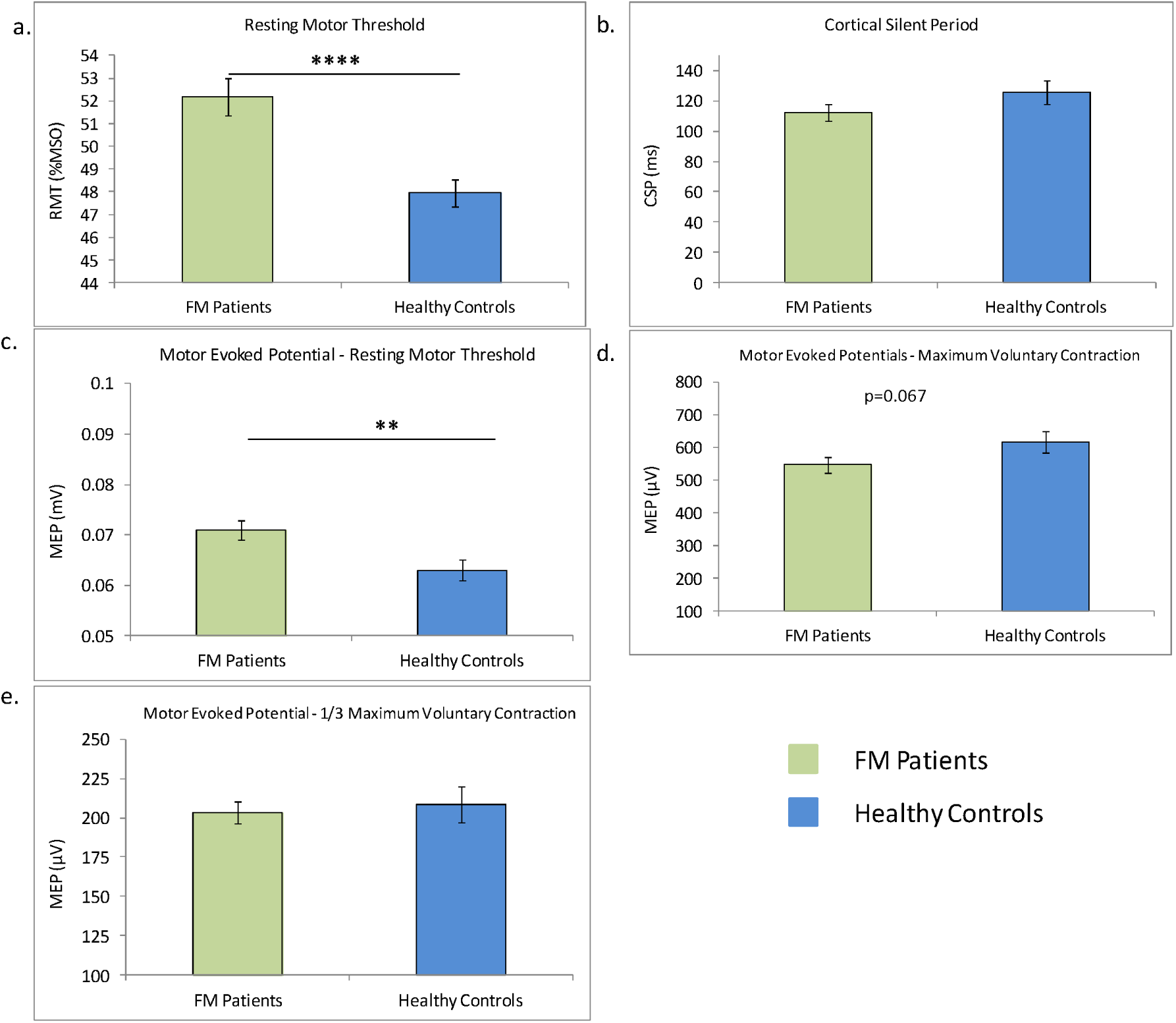
Changes in cortical excitability parameters in fibromyalgia patients and healthy controls: a) Resting Motor Threshold (p<0.0001); b) Cortical Silent Period (p=0.157); c) Motor Evoked Potential (p=0.004); d) Motor Evoked Potential at maximum voluntary contraction (p=0.067), and e) Motor Evoked Potential at 1/3 ‘maximum voluntary contraction (p=0.684). Data was normally distributed (Gaussian distribution) – ‘Shapiro-Wilk test’; expressed in Mean ± SEM. Two sample (Independent) t-test was performed for comparison between fibromyalgia patients and healthy controls. p-value < 0.05 was considered significant. Asterisk (*) depicts a significant difference between the arms. “*” represents p<0.05; “**” represents p<0.01. “****” represents p<0.0001.

Motor activity of fibromyalgia patients and healthy controls were measured by recording electromyographic response and peak-to-peak amplitude of MEP at Maximum Voluntary Contraction (MVC) and one-third of the MVC; we have not found any significant difference between the groups for both the MEP amplitude, at MVC (Fibromyalgia Patients = 546.51 ± 22.52 μV and Healthy Controls = 618.12 ± 33.15 μV; p = 0.067) and at 1/3 of MVC (Fibromyalgia Patients = 203.42 ± 7.31 μV and Healthy Controls = 208.72 ± 11.50 μV; p = 0.684). (Table 2 & Figure 9c-d) A recruitment curve for the Motor Evoked Potential (MEP) and Cortical Silent Period (CSP) for the fibromyalgia patients and healthy controls were plotted from their respective individual data values, ranging 90 % to 150 % of the RMT with a step-up of 10 % MSO of the RMT (Table 2). We did not found any significant difference between the MEP values of fibromyalgia patients and healthy controls at 90 % (Fibromyalgia Patients = 0.041 ± 0.004 mV and Healthy Controls = 0.030 ± 0.002 mV; p = 0.048) and RMT (Fibromyalgia Patients = 0.106 ± 0.013 mV and Healthy Controls = 0.065 ± 0.002 mV; p = 0.106) 110 % (Fibromyalgia Patients = 0.324 ± 0.056 mV and Healthy Controls = 0.339 ± 0.061mV; p = 0.180), 120 % (Fibromyalgia Patients = 0.581 ± 0.065 mV and Healthy Controls = 1.048 ± 0.71 mV; p = 0.961), 130 % (Fibromyalgia Patients = 1.013 ± 0.107 mV and Healthy Controls = 1.048 ± 0.71 mV; p = 0.801), 140 % (Fibromyalgia Patients = 1.418 ± 0.160 mV and Healthy Controls = 1.706 ± 0.148 mV; p = 0.206) and 150 % (Fibromyalgia Patients = 2.103 ± 0.238 mV and Healthy Controls = 2.000 ± 0.134 mV; p = 0.767) of the RMT (Table 2 & Figure 10a).

**Figure 10.**
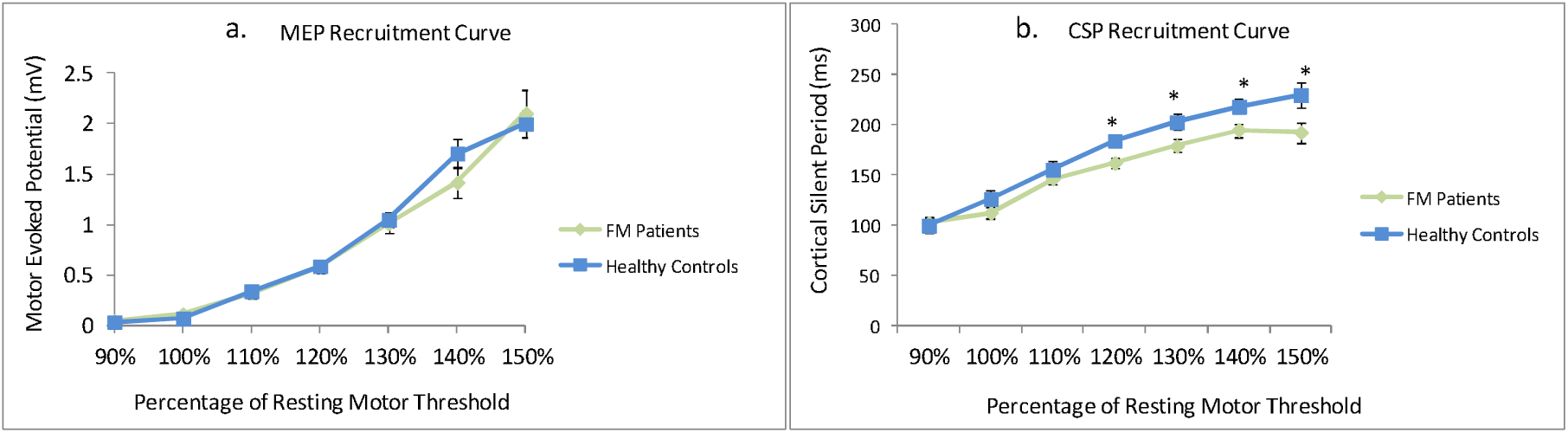
Recruitment curves of fibromyalgia patients and healthy controls: a) Motor Evoked Potential, and b) Cortical Silent Period. Data was normally distributed (Gaussian distribution) – ‘Shapiro-Wilk test’; expressed in Mean ± SEM. Two sample (Independent) t-test was performed for comparison between fibromyalgia patients and healthy controls. p-value < 0.05 was considered significant. Asterisk (*) depicts a significant difference between the arms. “*” represents p<0.05.

Cortical Silent Period (CSP) data for fibromyalgia patients and healthy controls were recorded for MSO values varying from 90 % to 150 % of the RMT (n= 60 each). Silent periods at 90 % (Fibromyalgia Patients = 102.05 ± 4.97 ms and Healthy Controls = 99.22 ± 7.95 ms; p = 0.751), 100 % (Fibromyalgia Patients = 112.25 ± 5.69 ms and Healthy Controls = 125.89 ± 8.03 ms; p = 0.157) and 110 % (Fibromyalgia Patients = 146.06 ± 5.81 ms and Healthy Controls = 155.31 ± 8.44 ms; p = 0.352) of the RMT have not shown any significant change between the fibromyalgia patients and healthy controls. For CSP values at 120 % (Fibromyalgia Patients = 161.74 ± 5.85 ms and Healthy Controls = 183.91 ± 6.97 ms; p = 0.017), 130 % (Fibromyalgia Patients = 178.92 ± 6.57 ms and Healthy Controls = 202.67 ± 8.25 ms; p = 0.025), 140 % (Fibromyalgia Patients = 194.89 ± 6.66 ms and Healthy Controls = 218.23 ± 7.37 ms; p = 0.024) and 150 % (Fibromyalgia Patients = 192.50 ± 10.54 ms and Healthy Controls = 229.56 ± 12.84 ms; p = 0.049) of the RMT, we reported a significantly higher silent period, on comparison between the respective data sets of fibromyalgia patients and age and gender-matched healthy controls (Table 2 & Figure 10b). Recruitment curves for the MEP and CSP for the respective data sets are depicted in Figure 10 for the fibromyalgia patients and healthy controls.

### Quantification of blood biomarkers in fibromyalgia patients and healthy controls (n = 82)

Concentration of Endorphin (Fibromyalgia Patients = 507.24 ± 40.20 pg/mL and Healthy Controls = 671.36 ± 37.62 pg/mL; p = 0.0087) and Cortisol (Fibromyalgia Patients = 97.42 ± 6.15 ng/mL and Healthy Controls = 17.84 ± 2.91 ng/mL; p<0.0001) were significantly different in fibromyalgia patients from the healthy controls. Endorphin level was quite lower in serum samples of fibromyalgia patients whereas the level of cortisol was lower in healthy controls, indicating poor quality of life and higher stress level in patients with fibromyalgia. Glutamate level was found significantly elevated in fibromyalgia patients (Fibromyalgia Patients = 234.86 (157.69, 421.02) ng/mL and Healthy Controls = 21.58 (16.93, 39.81) ng/mL; p<0.0001). (Table 2 & Figure 11a-c) Concentration of Serotonin (Fibromyalgia Patients = 35.79 (24.78, 61.67) ng/mL and Healthy Controls = 960.34 (60.71, 116.73) ng/mL; p<0.0001) and Substance P (Fibromyalgia Patients = 313.19 (116.87, 405.59) pg/mL and Healthy Controls = 53.01 (23.44, 62.47) pg/mL; p<0.0001) were reported significantly lower in fibromyalgia patients and healthy controls respectively. (Table 2 & Figure 11d-e)

**Figure 11.**
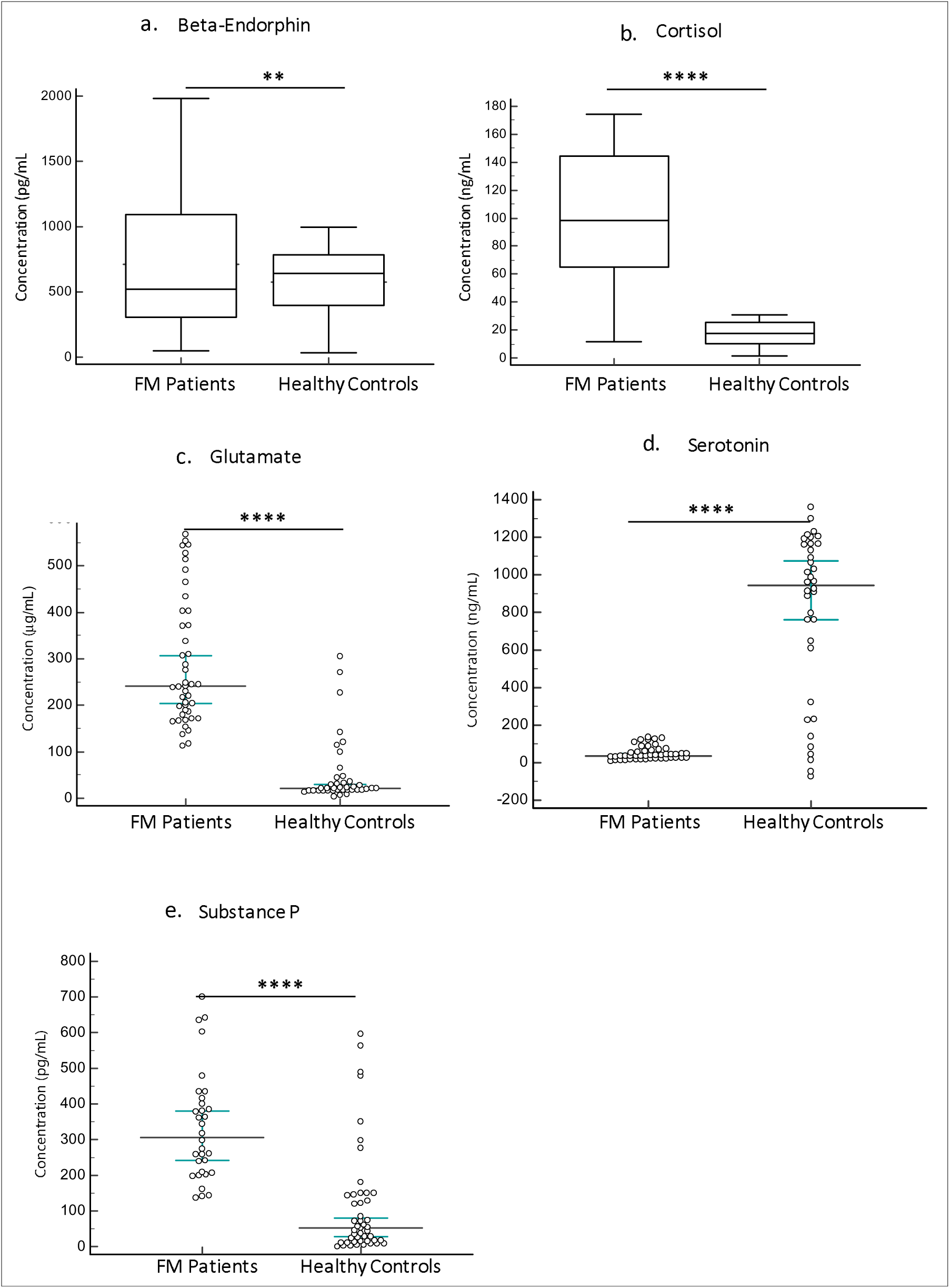
Comparison of levels of pain and related biomarkers in fibromyalgia patients and Healthy Controls: a) β-Endorphin (p=0.008); b) Cortisol (p<0.0001); c) Glutamate (p<0.0001); d) Serotonin (p<0.0001), and e) Substance P (p<0.0001). Data was normally distributed (Gaussian distribution) – ‘Shapiro-Wilk test’; expressed in Mean ± SEM otherwise expressed in Median (Q1, Q3). Two sample (Independent) t-test was performed for comparison between fibromyalgia patients and healthy controls. p-value < 0.05 was considered significant. Asterisk (*) depicts significant difference between the arms. “****” represents p<0.0001.

### Results of the Interventional Study (n = 60)

Primary objective was to study the effect of yogic intervention on the following aspects in 60 fibromyalgia patients (a) Pain status, (b) Quality of life, (c) Flexibility and range of motion, (d) Cortical excitability, and (e) Levels of blood biomarkers (β-Endorphin, Cortisol, Glutamate, Serotonin and Substance P). There were only two left-handed fibromyalgia patients (1 male and 1 female).

### Physiological characteristics of fibromyalgia patients of interventional study (n = 60)

At baseline average weight and BMI of the fibromyalgia patients were 62.47 ± 1.28 Kg and 25.79 ± 0.48 Kg/m^2^ respectively; which after 28 days of yogic intervention was reported 62.14 ± 1.22 Kg and 25.62 ± 0.45 Kg/m^2^, suggesting no significant change (p-value = 0.734 and 0.707 respectively). Heart rate of the patients enrolled in the yoga program was 83.97 ± 1.79 beats/minute at baseline which thereafter intervention was 84.31 ± 1.74 beats/minute, with non-significant difference (p-value = 0.861) (Table 3). Blood pressure of fibromyalgia patients was also measured pre- and post-yoga; systolic and diastolic blood pressure was 120.70 ± 1.84 mmHg and 83.69 ± 1.27 mmHg respectively. We found significant reduction in the systolic blood pressure (116.59 ± 1.45 mmHg; p-value = 0.025) after yogic intervention whilst the diastolic blood pressure remains significantly unchanged (81.74 ± 1.20 mmHg; p-value = 0.174) as represented in Table 3 and Figure 12a-b.

**Figure 12.**
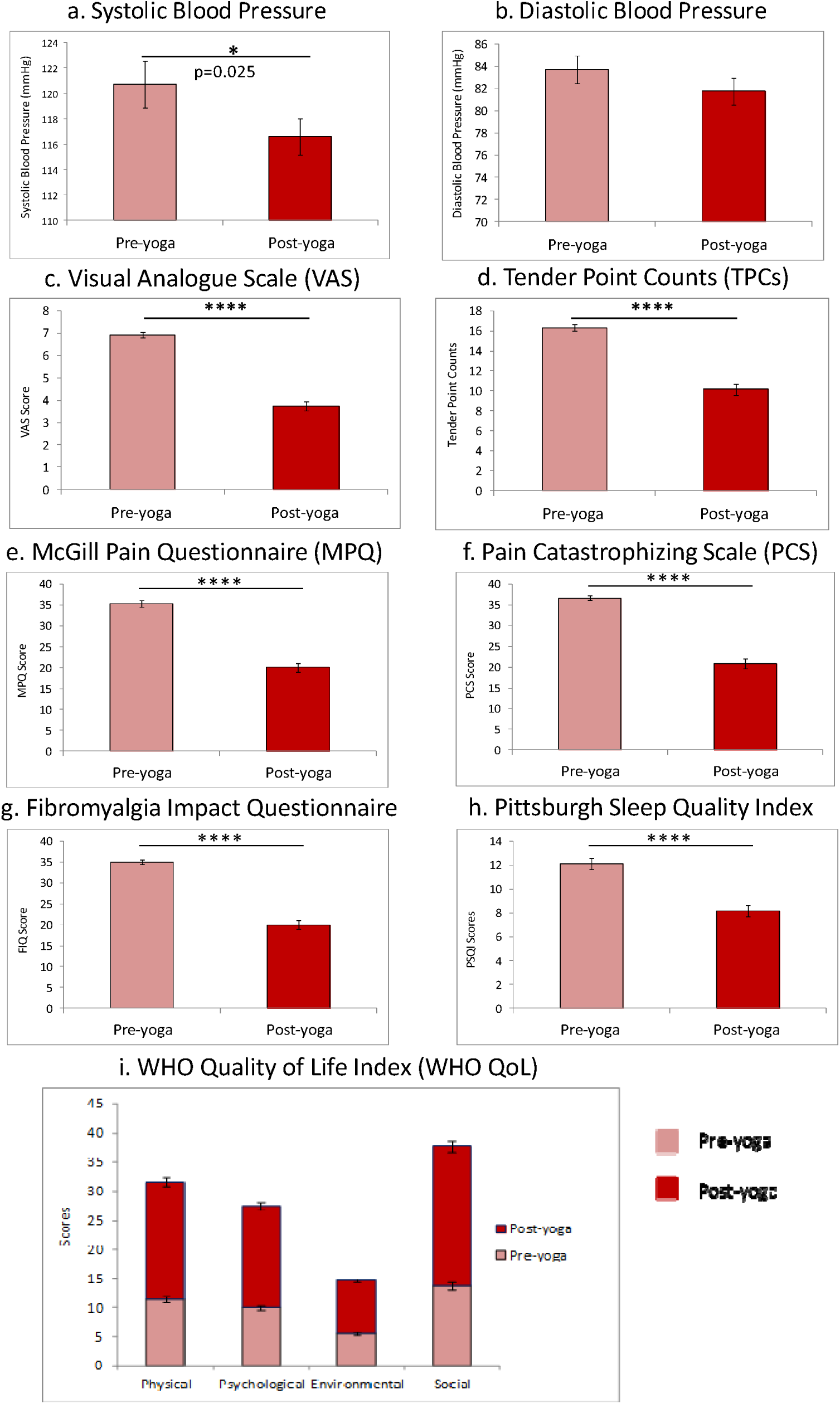
Effect of yogic intervention on pain and associated symptoms of fibromyalgia: a) Systolic Blood pressure; b) Diastolic Blood pressure of fibromyalgia patients; c) Visual Analog Scale (VAS) score; d) Tender Point Counts; e) McGill Pain Questionnaire; f) Pain Catastrophizing Scale; g) Fibromyalgia Impact Questionnaire; h) Pittsburgh Sleep Quality Index; and i) Various domains of WHO-Quality of Life. Data was normally distributed (Gaussian distribution) – ‘Shapiro-Wilk test’; expressed in Mean ± SEM. Paired t-test was performed for comparison between pre-yoga and post-yoga assessment parameters. p-value < 0.05 was considered significant. Asterisk (*) depicts significant differences post therapy. “****” represents p<0.0001. MPQ - McGill Pain Questionnaire; PSQI – Pittsburgh Sleep Quality Index; FIQ – Fibromyalgia Impact Questionnaire; PCS – Pain Catastrophizing Scale.

**Table 3:** Study parameters of fibromyalgia patients at baseline and post yoga in the interventional study.

| S. No. | Parameters (Units) | Pre-yoga | Post-yoga | p-value |
| --- | --- | --- | --- | --- |
| 1. | **Weight (Kg)** | 62.47 ± 1.28 | 62.14 ± 1.22 | 0.734 |
| 2. | **BMI (Kg/m<sup>2</sup>)** | 25.79 ± 0.48 | 25.62 ± 0.45 | 0.707 |
| 3. | **Heart Rate (Beats/min)** | 83.97 ± 1.79 | 84.31 ± 1.74 | 0.861 |
| 4. | **Blood Pressure (mmHg)**<br>Systolic<br>Diastolic | 120.70 ± 1.84<br>83.69 ± 1.27 | 116.59 ± 1.45<br>81.74 ± 1.20 | **0.025**<br>0.174 |
| 5. | **VAS Score** | 6.92 ± 0.12 | 3.73 ± 0.21 | **<0.0001** |
| 6. | **Tender Points Counts** | 16.33 ± 0.32 | 10.15 ± 0.57 | **<0.0001** |
| 7. | **MPQ Score** | 35.31 ± 0.77 | 20.07 ± 1.11 | **<0.0001** |
| 8. | **PCS Score** | 36.62 ± 0.57 | 20.84 ± 1.13 | **<0.0001** |
| 9. | **FIQ Score** | 35.05 ± 0.53 | 19.99 ± 1.11 | **<0.0001** |
| 10. | **WHO-QoL Score**<br>Physical<br>Psychological<br>Environmental<br>Social | 11.38 ± 0.54<br>10.02 ± 0.38<br>5.51 ± 0.24<br>13.81 ± 0.63 | 20.20 ± 0.71<br>17.43 ± 0.65<br>9.16 ± 0.35<br>23.85 ± 0.85 | **<0.0001**<br>**<0.0001**<br>**<0.0001**<br>**<0.0001** |
| 11. | **PSQI Score** | 12.11 ± 0.49 | 8.15 ± 0.46 | **<0.0001** |
| 12.<br>a. | **Quantitative Sensory Testing**<br>Pressure Pain Threshold<br>*Dorsum of Hand (KPa)*<br>*Left Shoulder (KPa)*<br>*Right Shoulder (KPa)*<br>*Lower Back (KPa)* | 166.96 ± 6.59<br>132.19 ± 7.48<br>132.31 ± 7.79<br>178.90 ± 9.23 | 159.91 ± 6.50<br>149.54 ± 7.26<br>149.85 ± 6.89<br>202.88 ± 8.20 | 0.443<br>0.064<br>0.062<br>**0.009** |
| b. | Pressure Pain Tolerance<br>*Dorsum of Hand (KPa)*<br>*Left Shoulder (KPa)*<br>*Right Shoulder (KPa)*<br>*Lower Back (KPa)* | 246.75 ± 10.36<br>211.84 ± 9.94<br>204.82 ± 10.18<br>266.79 ± 11.74 | 249.27 ± 8.32<br>228.01 ± 9.22<br>234.05 ± 10.59<br>301.47 ± 11.92 | 0.844<br>0.216<br>**0.037**<br>**0.014** |
| 13.<br>a.<br>b.<br>c. | **Flexibility & Range of Motion**<br>Schober's Test (cm)<br>Goniometry Lateral Bending (°)<br>*Left*<br>*Right*<br>Sit and Reach Test (cm) | 1.76 ± 0.11<br>15.07 ± 0.48<br>15.72 ± 0.50<br>12.29 ± 0.72 | 2.96 ± 0.14<br>21.87 ± 0.53<br>22.84 ± 0.52<br>17.86 ± 0.86 | **<0.0001**<br>**<0.0001**<br>**<0.0001**<br>**<0.0001** |
| 14.<br>a.<br>b.<br>c.<br>d.<br>e. | **Cortical Excitability**<br>RMT (% MSO)<br>MEP at RMT (mV)<br>CSP at RMT (ms)<br>MEP at MVC (μV)<br>MEP at 1/3 MVC (μV) | 53.60 ± 0.96<br>0.082 ± 0.01<br>117.29 ± 6.42<br>547.02 ± 19.94<br>194.08 ± 7.50 | 50.45 ± 0.96<br>0.073 ± 0.01<br>107.35 ± 4.88<br>597.81 ± 23.28<br>210.43 ± 7.78 | **0.004**<br>**0.012**<br>0.246<br>**0.05**<br>**0.043** |
| 15. | **Levels of Blood Biomarkers** |  |  |  |
| a. | β-endorphin (pg/mL) | 637.24 ± 40.20 | 719.90 ± 42.99 | 0.145 |
| b. | Cortisol (ng/mL) | 97.42 ± 6.15 | 112.60 ± 10.13 | 0.157 |
| c. | Glutamate (μg/mL) | 237.67 (167.96, 279.87) | 206.49 (162.84, 320.58) | 0.605 |
| d. | Serotonin (ng/mL) | 38.55 (23.43, 85.30) | 38.88 (29.40, 64.58) | 0.388 |
| e. | Substance P (pg/mL) | 72.94 (20.90, 149.07) | 40.23 (12.45, 107.23) | 0.097 |

*Data is represented as Mean ± SEM. Data was normally distributed (Gaussian distribution) – ‘Shapiro-Wilk test’; expressed in Mean ± SEM. Paired t-test was performed for comparison between pre-yoga and post-yoga assessment parameters. Glutamate, Serotonin and Substance P data were non-parametric – ‘Shapiro-Wilk test’; expressed in Median (Q1, Q3). Paired t-test and Signed rank sum test (Mann-Whitney U test) was performed for comparison between pre-yoga and post-yoga assessment for parametric data and non-parametric data, respectively. VAS – Visual Analog Scale; MPQ – McGill Pain Questionnaire; WHOQOL – World Health Organisation Quality of Life; PSQI – Pittsburgh Sleep Quality Index; FIQ – Fibromyalgia Impact Questionnaire; PCS – Pain Catastrophizing Scale; KPa – KiloPascal; MEP - Motor Evoked Potential; MVC - Maximum Voluntary Contraction.*

### Subjective assessment of pain and associated symptoms of fibromyalgia of interventional study (n = 60)

Average pain experienced by the patient on the numerical pain rating scale, Visual Analogue Scale (VAS) score was found significantly reduced after yogic intervention (Pre-yoga: 6.92 ± 0.12 and Post-yoga: 3.73 ± 0.21; p<0.0001). Fibromyalgia patients had a total of 16.33 ± 0.32 tender points at baseline which has significantly reduced to 10.15 ± 0.57 (p<0.0001) after four weeks of yogic intervention. Subjective assessment of pain was performed using short form McGill Pain Questionnaire (SF-MPQ) and Pain Catastrophizing Scale (PCS); both MPQ and PCS scores were found significantly reduced after the intervention (MPQ Score: Pre-yoga = 35.31 ± 0.77 and Post-yoga = 20.07 ± 1.11; PCS scores: Pre-yoga = 36.62 ± 0.57 and Post-yoga = 20.84 ± 1.13; p-value for both were <0.0001). (Table 3 & Figure 12c-g)

We assessed fibromyalgia impact using Fibromyalgia Impact Questionnaire (FIQ) and found a lesser impact on patients post-yoga (Pre-yoga = 35.05 ± 0.53; Post-yoga = 19.99 ± 1.11; p<0.0001). Pittsburgh Sleep Quality Index (PSQI) measures sleep quality in patients suffering from disturbed sleep due to psychological distress or chronic pain. Sleep quality in fibromyalgia patients was found significantly improved after four weeks of yogic intervention (Pre-yoga = 12.11 ± 0.49 and Post-yoga = 8.15 ± 0.46; p<0.0001). (Figure 12h) Quality of life of fibromyalgia patients when assessed using WHO-QOL-BREF questionnaire for all the four domains: physical, psychological, environmental and social; a significant improvement was reported – physical domain (Pre-yoga = 11.38 ± 0.54 and Post-yoga = 20.20 ± 0.71; p<0.0001), psychological domain (Pre-yoga = 10.02 ± 0.38 and Post-yoga = 17.43 ± 0.65; p<0.0001), environmental domain (Pre-yoga = 5.51 ± 0.24 and Post-yoga = 9.16 ± 0.35; p<0.0001) and social domain (Pre-yoga = 13.81 ± 0.63 and Pot-yoga = 23.85 ± 0.85; p<0.0001). (Figure 12i)

### Objective assessment of pain using quantitative sensory testing (n = 59)

Both the pressure pain threshold and pressure pain tolerance were higher at the reference site as compared to the left and right shoulders at baseline and post-intervention. Pressure pain threshold at dorsum of hand (Pre-yoga = 166.96 ± 6.59 kPa and Post-yoga = 159.91 ± 6.50 kPa; p = 0.443), left shoulder (Pre-yoga = 132.19 ± 7.48 kPa and Post-yoga = 149.54 ± 7.26 k Pa; p = 0.064) and right shoulder (Pre-yoga = 132.31 ± 7.79 kPa and Post-yoga = 149.85 ± 6.89 kPa; p = 0.062) didn’t show any significant change after four weeks of yogic intervention; but the lower back pressure pain threshold was found significantly increased (Pre-yoga = 178.90 ± 9.23 kPa and Post-yoga = 202.88 ± 8.20 kPa; p = 0.009) after the yogic intervention (Table 3, Figure 13a). Although pressure pain thresholds for left and right shoulders were found harnessed by yoga practices for four weeks, but they didn’t achieve set significance level. Dorsum of hand (Pre-yoga = 246.75 ± 10.36 kPa and Post-yoga = 249.27 ± 8.32 kPa; p = 0.844) and left shoulder’s (Pre-yoga = 211.84 ± 9.94 kPa and Post-yoga = 228.01 ± 9.22 kPa; p = 0.216) pressure pain tolerance remain unaltered after four weeks of yogic intervention in fibromyalgia patients; but for right shoulder (Pre-yoga = 204.82 ± 10.18 kPa and Post-yoga = 234.05 ± 10.59 kPa; p = 0.037) and lower back (Pre-yoga = 266.79 ± 11.74 kPa and Post-yoga = 301.47 ± 11.92 kPa; p = 0.014), tolerance was found significantly increased (Figure 13b).

**Figure 13.**
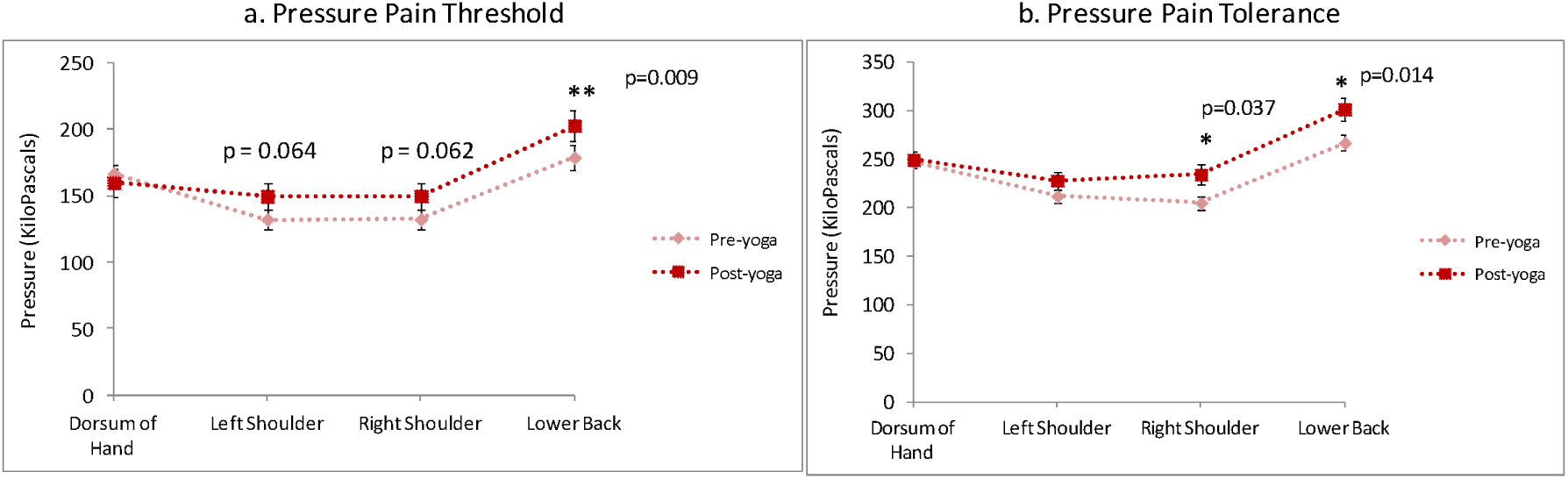
Assessment of pressure pain parameters using quantitative sensory testing (QST) at dorsum of hand, shoulders and lower back of fibromyalgia patients. a) Pressure pain threshold and b) Pressure pain tolerance were measured at the same site in continuation. Data was normally distributed (Gaussian distribution) – ‘Shapiro-Wilk test’; expressed in Mean ± SEM. Paired t-test was performed for comparison between pre-yoga and post-yoga assessment parameters. p-value < 0.05 was considered significant. Asterisk (*) depicts significant difference post therapy. “*” represents p<0.05; “**” represents p<0.01.

### Assessment of flexibility and range of motion of fibromyalgia patients of the interventional study (n = 60)

Lumbar flexion and range of motion in patients with fibromyalgia (n = 60) were assessed using modified Schober’s test, Goniometry and Sit and Reach box. After four weeks of regular yoga practice participants showed an increase in all the measures of lumbar flexion and range of motion (Table 3). Schober’s test showed significant elevation in the flexion (Pre-yoga = 1.76 ± 0.11 cm and Post-yoga = 2.96 ± 0.14 cm; p<0.0001). Lateral goniometry was used to measure left-right bending before and after yogic intervention in fibromyalgia patients (Figure 14a). We found a significant increase in flexibility and range of motion in both left (Pre-yoga = 15.07 ± 0.48 and Post-yoga = 21.87 ± 0.53 ; p<0.0001) and right (Pre-yoga = 15.72 ± 0.50 and Post-yoga = 22.84 ± 0.52 ; p<0.0001) directions. (Figure 14b) Furthermore, lumbar flexibility was found significantly increased (Pre-yoga = 12.29 ± 0.72 cm and Post-yoga = 17.86 ± 0.86 cm; p<0.0001) after yoga when assessed using Sit and Reach box. (Figure 14c)

**Figure 14.**
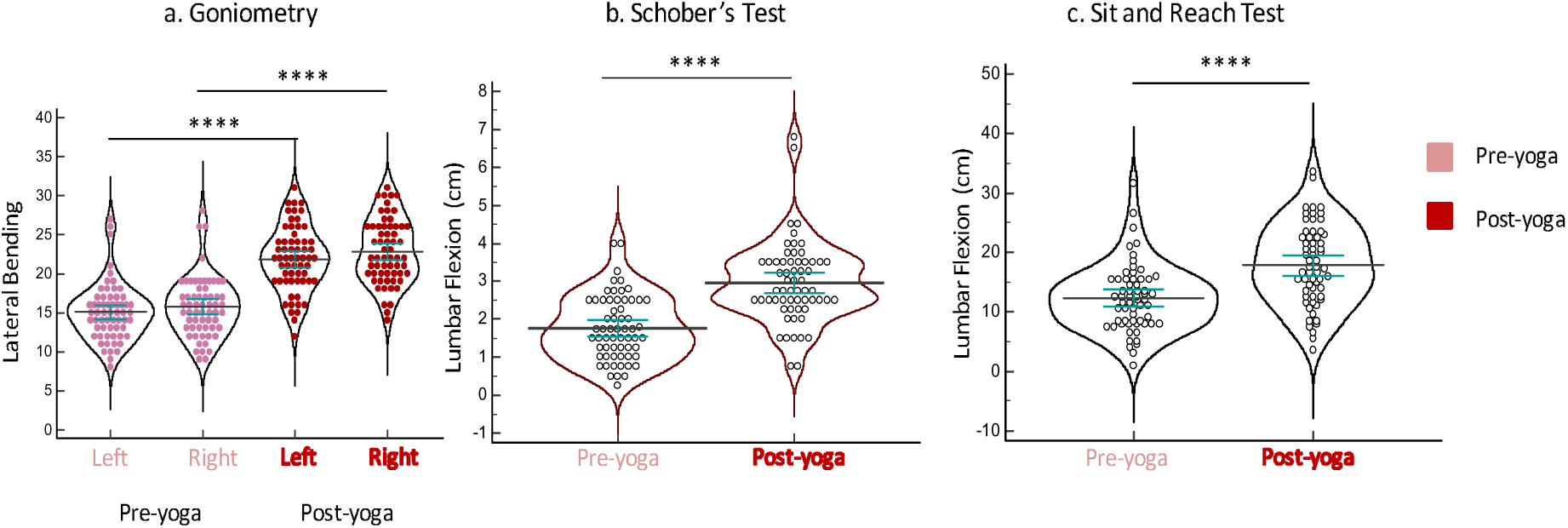
Comparison of pre-yoga and post-yoga flexibility and range of motion in fibromyalgia patients: a) Goniometry; b) Schober’s Test and, c) Sit and Reach test. Data was normally distributed (Gaussian distribution) – ‘Shapiro-Wilk test’; expressed in Mean ± SEM. Paired t-test was performed for comparison between pre-yoga and post-yoga assessment parameters. p-value < 0.05 was considered significant. Asterisk (*) depicts significant differences post therapy. “****” represents p<0.0001.

### Assessment of cortical excitability of fibromyalgia patients of interventional study (n = 60)

Resting Motor Threshold (RMT) of fibromyalgia patients after four weeks of yogic intervention was found significantly reduced (Pre-yoga = 53.60 ± 0.96 % MSO and Post-yoga = 50.45 ± 0.96 % MSO; p = 0.004) (Figure 15a). We have also found a significant decrease in Motor Evoked Potential (MEP) at RMT (Pre-yoga = 0.082 ± 0.01 mV and Post-yoga = 0.073 ± 0.01 mV; p = 0.012) after the intervention (Figure 15b). Cortical Silent Period (CSP) of the patients after the intervention didn’t differ significantly from the baseline (Pre-yoga = 117.29 ± 6.42 ms and Post-yoga = 107.35 ± 4.88 ms; p = 0.246) (Figure 15c). (Table 3) Motor activity of fibromyalgia patients were measured by recording electromyographic response and peak-to-peak amplitude of MEP at Maximum Voluntary Contraction (MVC) and one-third of the MVC; a significant increase in the MEP amplitude at MVC (Pre-yoga = 547.02 ± 19.94 μV and Post-yoga = 597.81 ± 23.28 μV; p = 0.05) and 1/3 of MVC (Pre-yoga = 194.08 ± 7.50 μV and Post-yoga = 210.43 ± 7.78 μV; p = 0.043) was observed after four weeks of supervised yoga practices (Figure 15d-e). A recruitment curve for the Motor Evoked Potential (MEP) and Cortical Silent Period (CSP) for the fibromyalgia patients were plotted from the respective individual data values, ranging 90 % to 150 % of the RMT with an escalation of 10 %, at baseline and after four weeks of yogic intervention (Table 3).

**Figure 15.**
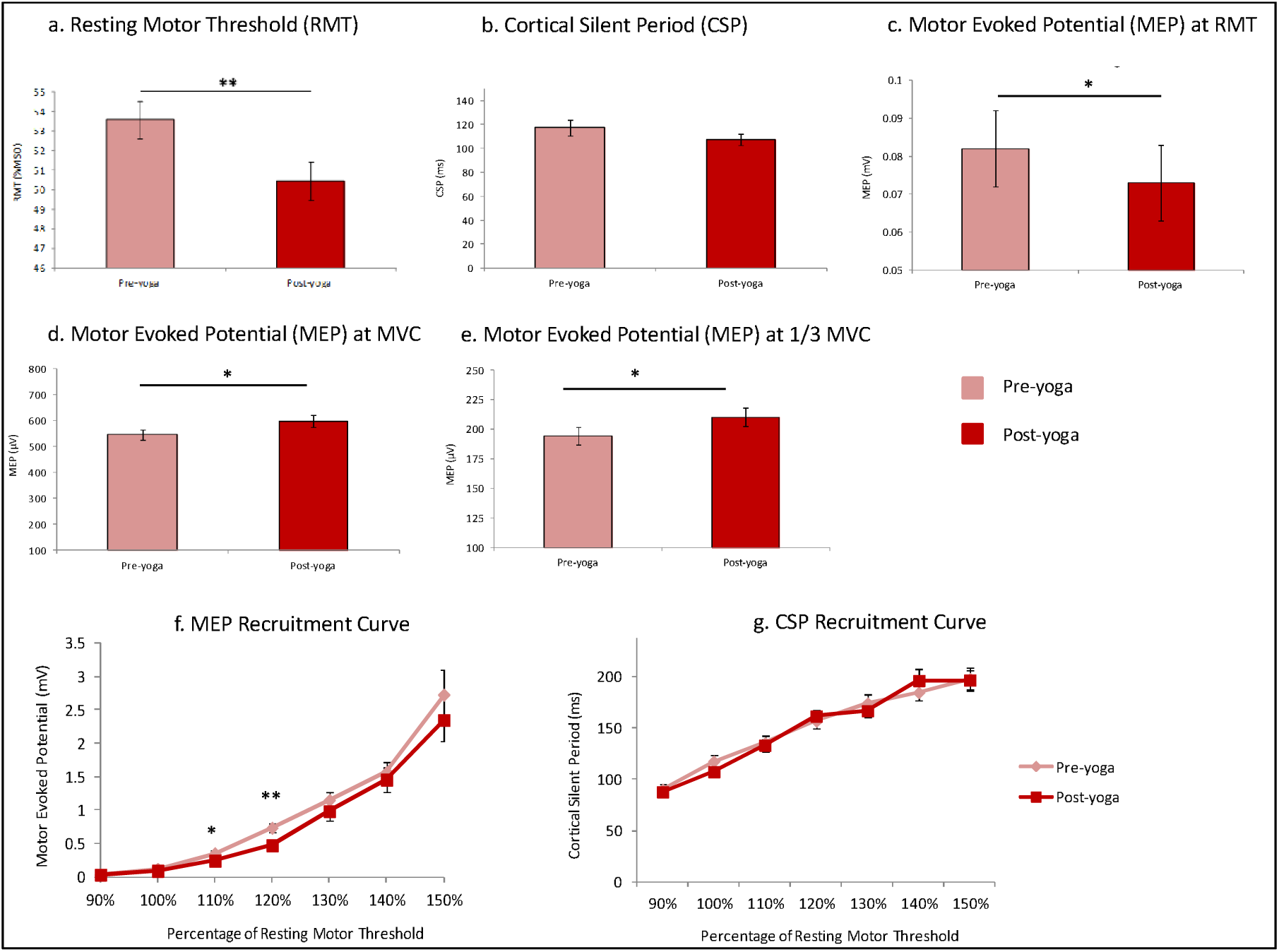
Changes in cortical excitability parameters in fibromyalgia patients post-yoga: a) Resting Motor Threshold (p=0.004); b) Cortical Silent Period (p=0.246); c) Motor Evoked Potential (p=0.012); d) Motor Evoked Potential at maximum voluntary contraction (p=0.05); e) Motor Evoked Potential at 1/3 ‘maximum voluntary contraction (p=0.043); f) Motor Evoked Potential (MEP) Recruitment curve; and g) Cortical Silent Period (CSP) Recruitment curve of fibromyalgia patients pre- and post-yoga. Data was normally distributed (Gaussian distribution) – ‘Shapiro-Wilk test’; expressed in Mean ± SEM. Paired t-test was performed for comparison between pre-yoga and post-yoga assessment parameters. p-value < 0.05 was considered significant. Asterisk (*) depicts significant differences post therapy.

We found a significant change in the MEP values at 110 % (Pre-yoga = 0.354 ± 0.038 mV and Post-yoga = 0.248 ± 0.022 mV; p = 0.021) and 120 % (Pre-yoga = 0.736 ± 0.070 mV and Post-yoga = 0.476 ± 0.051 mV; p = 0.002) of RMT whereas the MEP values remain statistically non-significant at 90 % (Pre-yoga = 0.038 ± 0.002 mV and Post-yoga = 0.036 ± 0.003 mV; p = 0.827), RMT (Pre-yoga = 0.120 ± 0.014 mV and Post-yoga = 0.094 ± 0.003 mV; p = 0.082), 130 % (Pre-yoga = 1.149 ± 0.113 mV and Post-yoga = 0.986 ± 0.141 mV; p = 0.280), 140 % (Pre-yoga = 1.570 ± 0.153 mV and Post-yoga = 1.457 ± 0.188 mV; p = 0.580) and 150 % (Pre-yoga = 2.725 ± 0.372 mV and Post-yoga = 2.345 ± 0.324 mV; p = 0.332) after four weeks of yogic intervention in fibromyalgia patients (Table 3 & Figure 15f). Cortical Silent Period (CSP) data were recorded for MSO values varying from 90 % to 150 % of the RMT, both at baseline (Pre-therapy) and after four weeks of yogic intervention (Post-therapy) in fibromyalgia patients (n= 60). Silent periods at 90 % (Pre-yoga = 89.95 ± 4.73 ms and Post-yoga = 87.76 ± 4.22 ms; p = 0.701), 100 % (Pre-yoga = 117.29 ± 6.42 ms and Post-yoga = 107.35 ± 4.88 ms; p = 0.246), 110 % (Pre-yoga = 135.19 ± 7.45 ms and Post-yoga = 133.06 ± 6.08 ms; p = 0.828), 120 % (Pre-yoga = 156.44 ± 6.94 ms and Post-yoga = 161.52 ± 5.95 ms; p = 0.513), 130 % (Pre-yoga = 174.03 ± 8.04 ms and Post-yoga = 166.49 ± 6.77 ms; p = 0.474), 140 % (Pre-yoga = 184.27 ± 7.28 ms and Post-yoga = 195.99 ± 10.84 ms; p = 0.383) and 150 % (Pre-yoga = 197.70 ± 10.40 ms and Post-yoga = 196.22 ± 10.32 ms; p = 0.911); on comparison between the time points, found statistically unchanged for all the data sets assessed at stepped-up percentages of MSO (Table 3 & Figure 15g). Recruitment curves for the MEP and CSP for the respective data sets are depicted in Figure 15f-g for the fibromyalgia patients at baseline and four weeks of regular yogic intervention.

### Assessment of blood biomarkers in fibromyalgia patients of the interventional study (n = 54)

Levels of pain and related blood biomarkers were quantified in isolated serum samples of the fibromyalgia patients at baseline and after four weeks of yogic intervention using Enzyme Linked Immunosorbent Assay (ELISA) (Table 3). Concentration of Β-Endorphin (Pre-yoga = 637.24 ± 40.20 pg/mL and Post-yoga = 719.90 ± 42.99 pg/mL; p = 0.145), Cortisol (Pre-yoga = 97.42 ± 6.15 ng/mL and Post-yoga = 112.60 ± 10.13 ng/mL; p = 0.157), Glutamate (Pre-yoga = 237.67 (167.96, 279.87) μg/mL and Post-yoga = 206.49 (162.84, 320.58) μg/mL; p = 0.605), Serotonin (Pre-yoga = 38.55 (23.43, 85.30) ng/mL and Post-yoga = 38.88 (29.40, 64.58) ng/mL; p = 0.388) and Substance P (Pre-yoga = 72.94 (20.90, 149.07) pg/mL and Post-yoga = 40.23 (12.45, 107.23) pg/mL; p = 0.097) didn’t show any significant change after the supervised yogic intervention (Table 3).

### Results of the Randomized Controlled Trial (n = 120)

Primary objective (Randomized Controlled Trial in 120 fibromyalgia patients) was to compare the measures of pain status and cortical excitability of fibromyalgia patients of yoga group and waitlisted controls. Participants of the yoga group were administered with four weeks of supervised yogic intervention whilst that of waitlisted controls were intervened with the standard medical therapy suggested/recommended/prescribed by the medical experts of the field for the same time period. Moreover, yoga group participants were on the ongoing pharmaceutical drugs prescribed by the medical practitioners.

### Demographic and clinical features of fibromyalgia patients of the randomized controlled trial (n = 120)

During medical history reporting, participants were asked about the demographic details such as their geographical location and socioeconomic status. There were five left-handed fibromyalgia patients. The prevalence of female fibromyalgia patients was found to be 0.81 in our cohort, and was affected in a ratio of around 1:5. Approximately, 72% fibromyalgia patients (n = 87) were married having 2.17 ± 1.4 children on average. Amongst fibromyalgia patients of our cohort, nearly half of the patients (49%) were non-working (mostly housewives). (Table 4) One-third (33 %) of the patients were having educational qualifications equal to or less than XII level and 40% of them have discontinued their academics beyond VIII level. Approximately half of the fibromyalgia patients (49%) were holding bachelor’s level of educational qualification. Masters above or equivalent level of academic qualifications were reported in around 18% of the patients of our cohort. (Figure 16d)

**Figure 16.**
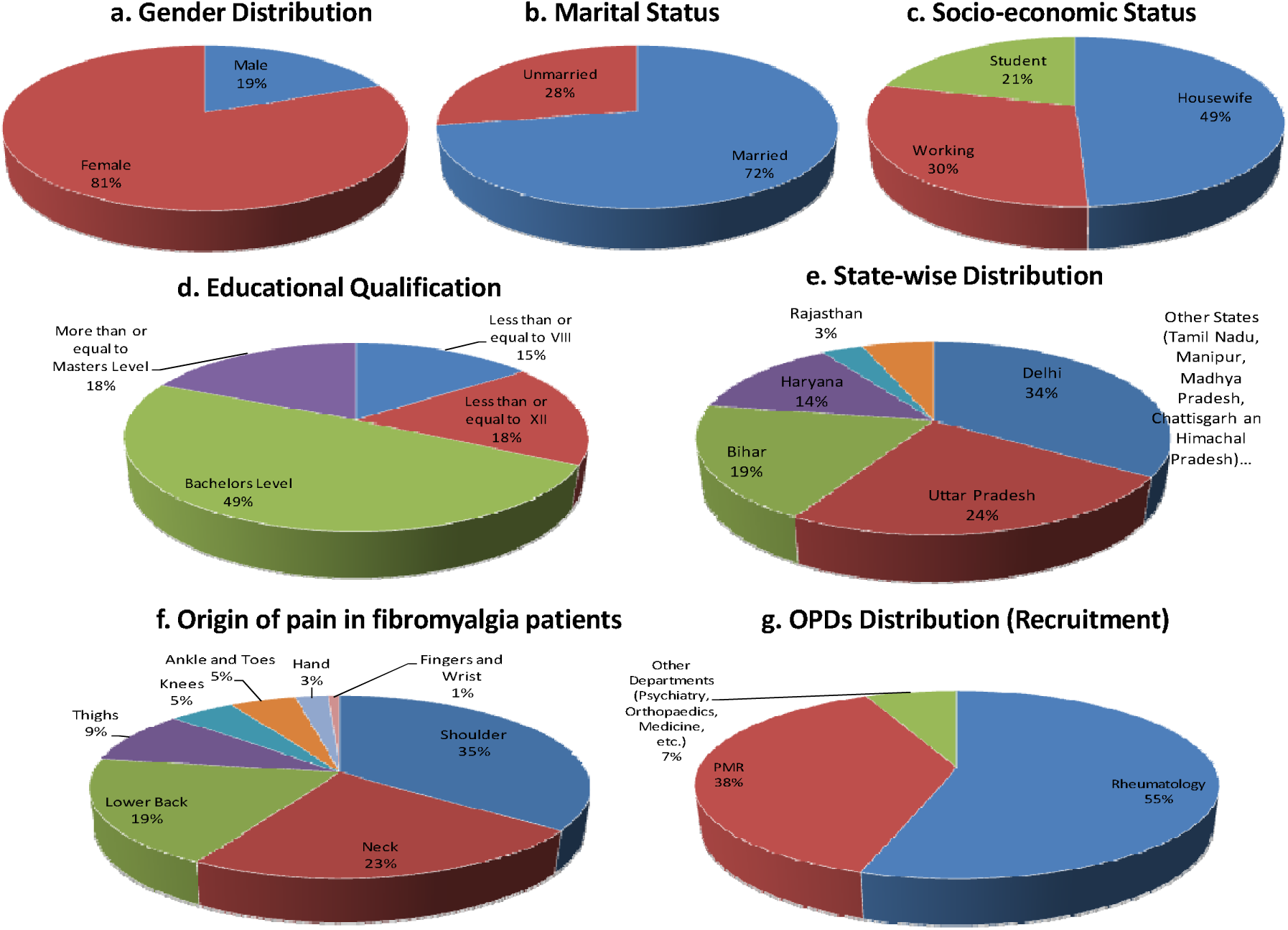
Demographic and clinical features of fibromyalgia patients a) Gender Distribution, b) Marital Status, c) Socio-economic status, d) Educational Qualifications and e) State-wise distribution; f) Origin of Pain and g) Patients referred from Out-patients Departments.

**Table 4:** Demographic and clinical characteristics of fibromyalgia patients of RCT at baseline.

| S. No. | Characteristics | Number of Patients | Proportion of Patients |
| --- | --- | --- | --- |
| **1.** | **Sex** |  |  |
|  | Male | 23 | 0.19 |
|  | Female | 97 | 0.81 |
| **2.** | **Marital Status** |  |  |
|  | Married* | 87 | 0.72 |
|  | Unmarried | 33 | 0.28 |
| **3.** | **Socioeconomic Status** |  |  |
|  | Housewife | 59 | 0.49 |
|  | Working | 36 | 0.30 |
|  | Student | 25 | 0.21 |
| **4.** | **Education** |  |  |
|  | Less than or equal to VIII | 18 | 0.15 |
|  | Less than or equal to XII | 21 | 0.18 |
|  | Bachelors Level | 59 | 0.49 |
|  | More than or equal to Masters Level | 22 | 0.18 |
| **5.** | **State-wise patients visiting OPDs** |  |  |
|  | Delhi | 41 | 0.35 |
|  | Uttar Pradesh | 29 | 0.24 |
|  | Bihar | 23 | 0.19 |
|  | Haryana | 16 | 0.13 |
|  | Rajasthan | 4 | 0.03 |
|  | Other States | 7 | 0.06 |
| **1.** | **Clinical Manifestations** |  |  |
|  | Genetic History | 32 | 0.27 |
|  | Sleep Problems | 64 | 0.53 |
|  | Post-Partum Pain Onset | 24 | 0.20 |
|  | Morning Stiffness | 76 | 0.63 |
| **2.** | **Origin of Pain** |  |  |
|  | Shoulder | 42 | 0.35 |
|  | Neck | 28 | 0.23 |
|  | Lower Back | 23 | 0.19 |
|  | Thighs | 11 | 0.09 |
|  | Knees | 6 | 0.05 |
|  | Hand | 6 | 0.05 |
|  | Ankle and Toes | 3 | 0.03 |
|  | Fingers and Wrist | 1 | 0.01 |
| **3.** | **OPDs** |  |  |
|  | Rheumatology | 66 | 0.55 |
|  | PMR | 46 | 0.38 |
|  | Other Departments | 8 | 0.07 |

**\*Denotes married patients bearing an average of ..... children.**

Three of every four fibromyalgia patients (75%) came from the northern part of India (Delhi – 35%, Uttar Pradesh – 24%, Haryana – 13%, and Rajasthan – 3%). Moreover approximately, 20.5% of patients visiting our OPDs were from eastern states (predominantly Bihar – 19% and Manipur – 0.9%). Patients from other regions such as central, (Madhya Pradesh and Chhattisgarh), western (Maharashtra) and southern (Telangana, Tamil Nadu) part of India consisted nearly 6%. Geographical distribution of patients visiting our clinic is given in Table 4 and Figure 16e.

We reported a genetic history of arthritis and other musculoskeletal symptoms in the parental or grand parental lineage in almost 27% (n=32) of the fibromyalgia patients. Sleep problems and morning stiffness were observed in 53% and 63% of the fibromyalgia patients respectively. Postpartum pain onset was subjectively reported in one-fifth of the patients of our cohort (n=24), which is nearly 25% of the female fibromyalgia patients and one-third of the married woman with fibromyalgia. Unilateral pain was reported in approximately one-fifth of the fibromyalgia patients. In most of the patients (77%) the origin of pain was reported from the neck to the hip (shoulder – 35%, neck – 23%, and lower back – 19%). Only 22% of the patients felt first pain in either upper or lower extremities (Leg: thighs: – 9%, Knees – 5% and ankle-toes – 3%; Hand: Hand as whole – 5%; Fingers and wrist – 1%) (Table 4 & Figure 16f). Most of the fibromyalgia patients (93%) in our study were referred either from the OPDs of department of Rheumatology (65%) or Physical Medicine and Rehabilitation (38%) of the same tertiary care hospital. Only 7% fibromyalgia patients were referred from other departments such as Psychiatry, Orthopaedics, and Medicine for clinical assessments (Table 4 & Figure 16g).

### Physiological characteristics of fibromyalgia patients of the randomized controlled trial (n=120)

Physiological characteristics of fibromyalgia patients such as weight, Body Mass Index, heart rate and blood pressure were taken at baseline and after four weeks of the therapy (yogic intervention in case of *Yoga Group* patients and standard therapy in *Waitlisted Control* patients). Average weight of the fibromyalgia patients of both arms were comparable at baseline (Pre-therapy: Yoga Group = 62.47 ± 1.28 Kg and Waitlisted Controls = 63.04 ± 1.34 Kg; p = 0.759); which were not significantly different at four weeks of interventions (Post-therapy: Yoga Group = 62.14 ± 1.23 Kg and Waitlisted Controls = 63.68 ± 1.43 Kg; p = 0.416). Neither the yoga group nor waitlisted control patients showed any significant change after four weeks of yogic intervention (Yoga Group: Pre-therapy = 62.47 ± 1.28 Kg and Post-therapy = 62.14 ± 1.23 Kg; p = 0.734) and standard therapy (Post-therapy: Yoga Group = 63.04 ± 1.34 Kg and Waitlisted Controls = 63.68 ± 1.43 Kg; p = 0.311) respectively (Table 5). Body mass Indices (BMIs) of fibromyalgia patients of both the study groups were comparable at the baseline (Pre-therapy: Yoga Group = 25.79 ± 0.48 Kg/m^2^ and Waitlisted Controls = 25.70 ± 0.51 Kg/m^2^; p = 0.895); which were not significantly different at four weeks of interventions (Post-therapy: Yoga Group = 25.62 ± 0.45 Kg/m^2^ and Waitlisted Controls = 25.90 ± 0.54 Kg/m^2^; p = 0.691). Both yoga group and waitlisted control patients didn’t show any significant change after four weeks of yogic intervention (Yoga Group: Pre-therapy = 25.79 ± 0.48 Kg/m^2^ and Post-therapy = 25.62 ± 0.45 Kg/m^2^; p = 0.707) and standard therapy (Post-therapy: Yoga Group = 25.70 ± 0.51 Kg/m^2^ and Waitlisted Controls = 25.90 ± 0.54 Kg/m^2^; p = 0.364) respectively (Table 5).

**Table 5:**
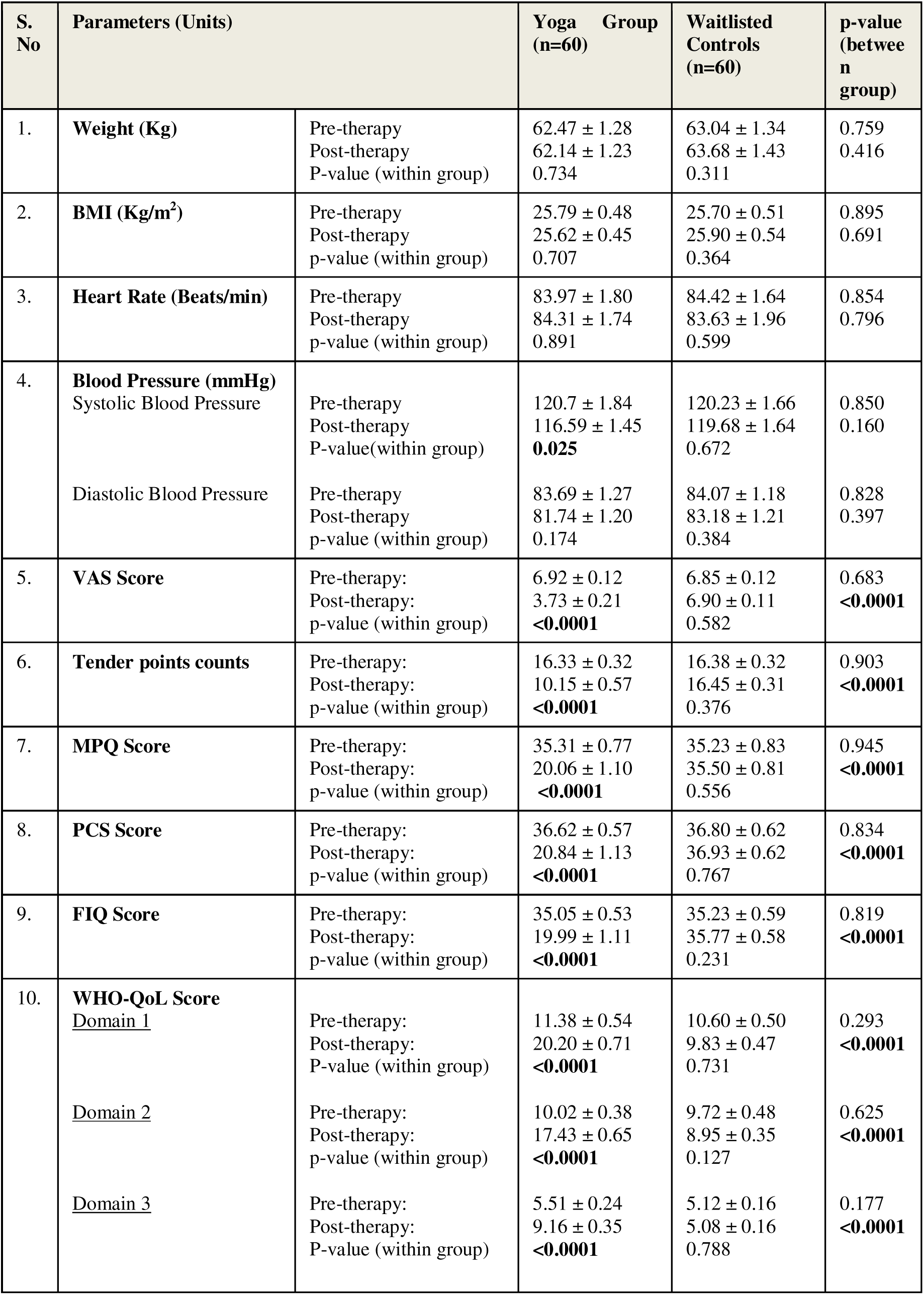
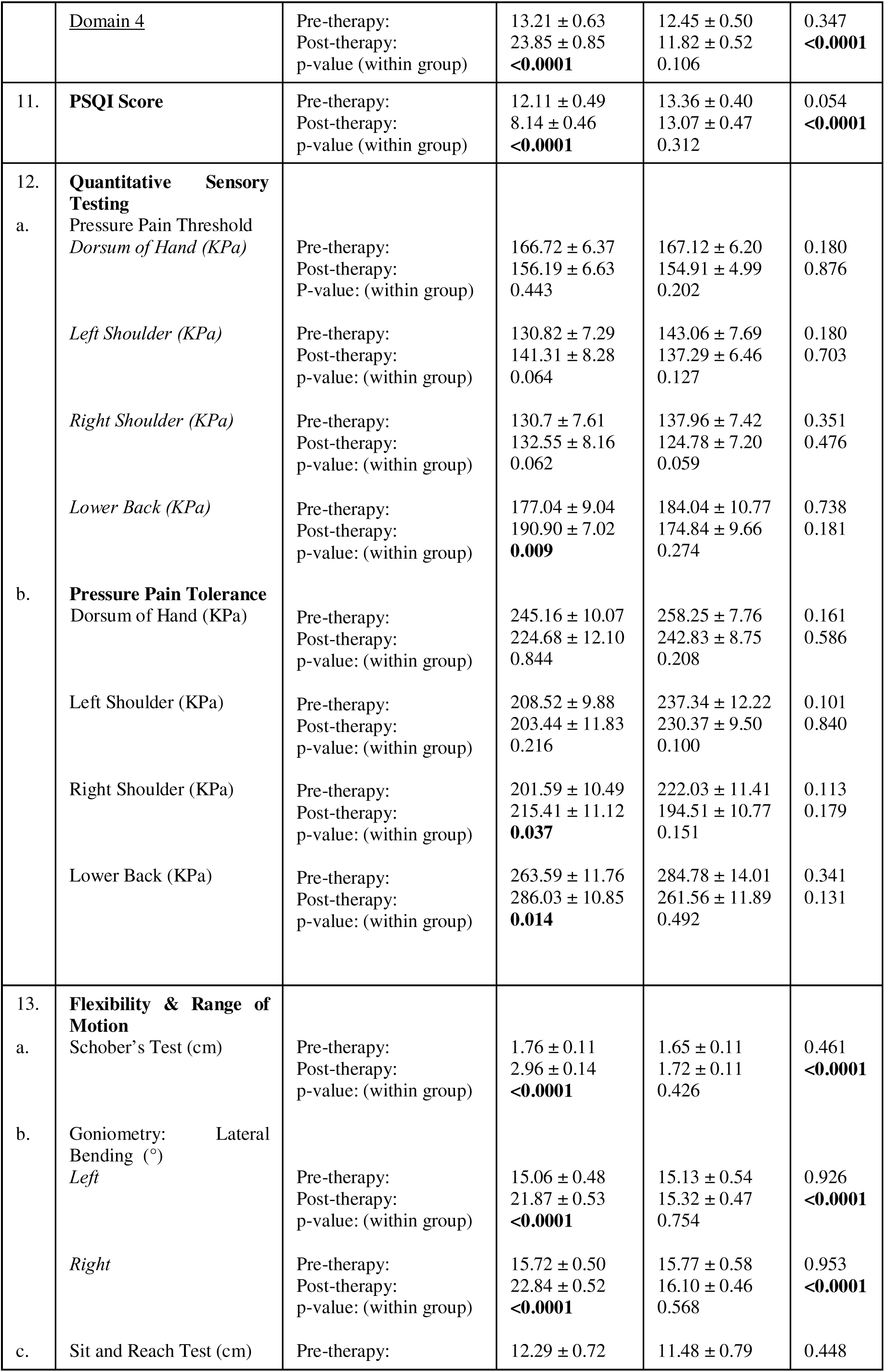
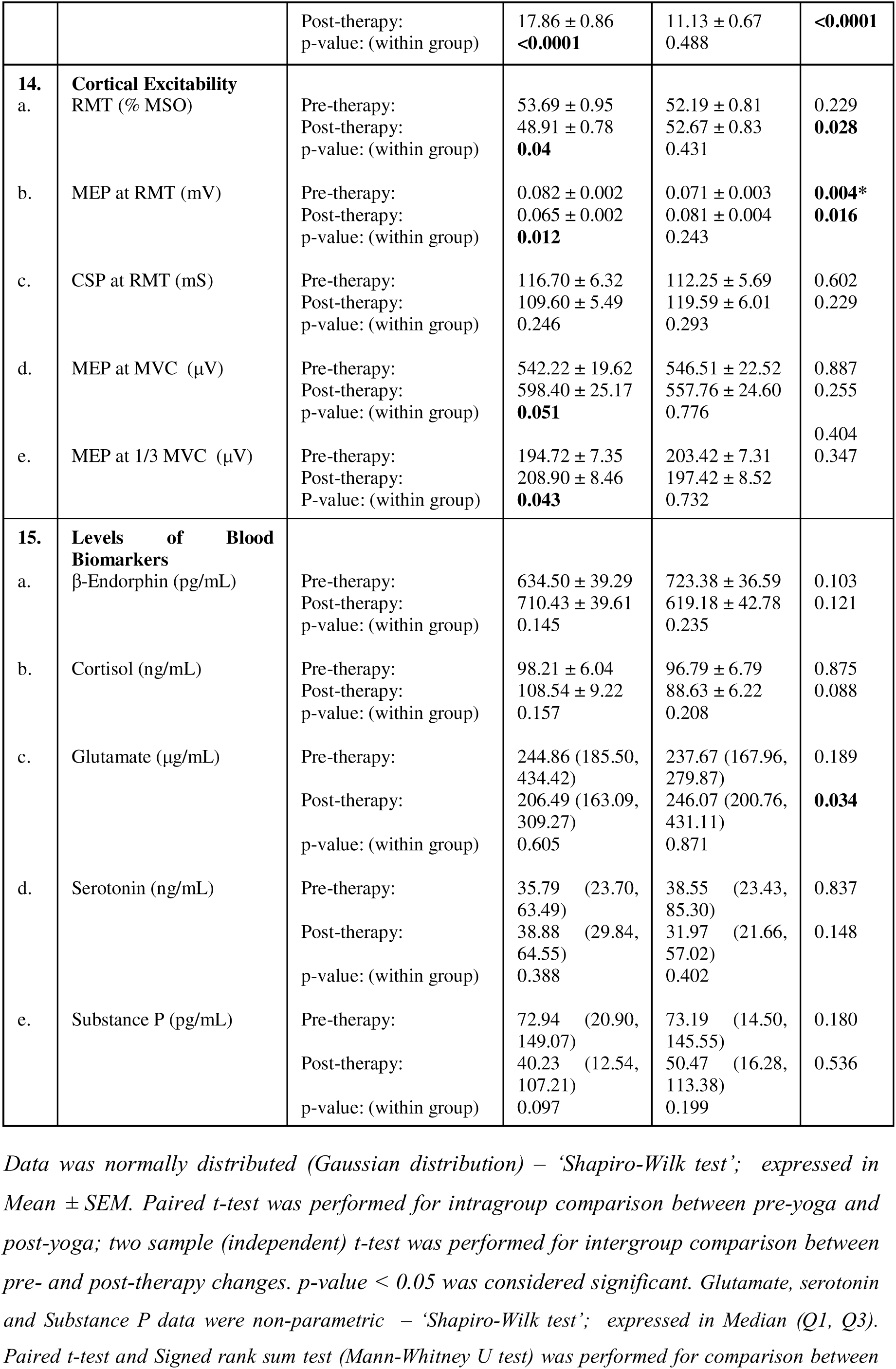
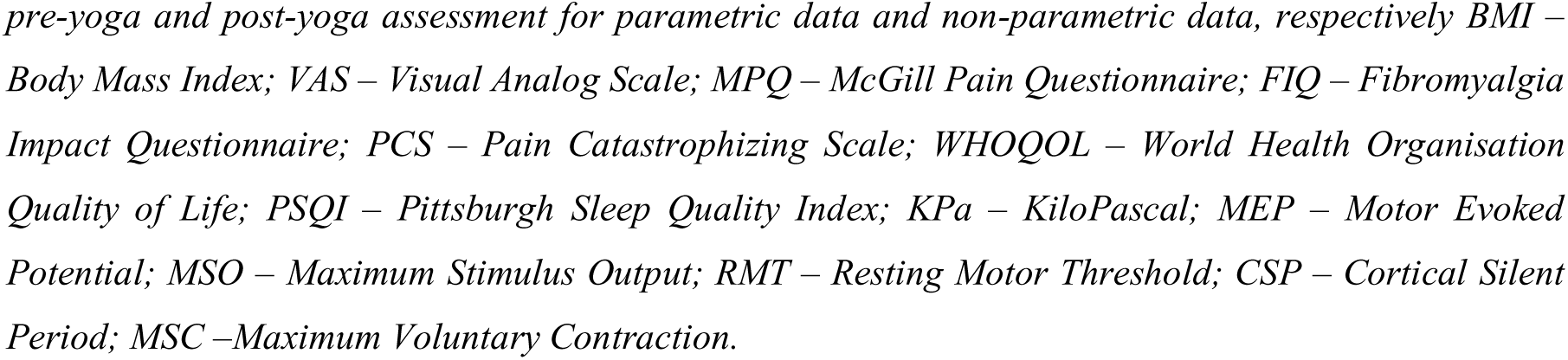
Effect of interventions on various study parameters of fibromyalgia patients of yoga group and waitlisted controls at baseline and post-therapy of the randomized controlled trial.

No significant difference was found at baseline measures of heart rates between the two study groups (Pre-therapy: Yoga Group = 83.97 ± 1.80 bpm and Waitlisted Controls = 84.42 ± 1.64 bpm; p = 0.854); Post-therapy heart rate of yoga group was also not different from that of waitlisted controls (Yoga Group = 84.31 ± 1.74 bpm and Waitlisted Controls = 83.63 ± 1.96 bpm; p = 0.796). Yogic intervention as well as standard therapy had no effect on heart rates of fibromyalgia patients (Yoga Group: Pre-therapy = 83.97 ± 1.80 bpm and Post-therapy = 84.31 ± 1.74; p = 0.891; Waitlisted Controls: Pre-therapy = 84.42 ± 1.64 bpm and Post-therapy = 83.63 ± 1.96 bpm; p = 0.599) (Table 5).

Both mean systolic (Pre-therapy: Yoga Group = 120.7 ± 1.84 mmHg and Waitlisted Controls = 120.23 ± 1.66 mmHg; p = 0.850) as well as diastolic blood pressures (Pre-therapy: Yoga Group = 83.69 ± 1.27 mmHg and Waitlisted Controls = 84.07 ± 1.18 mmHg; p = 0.828) were comparable at the baseline between fibromyalgia patients of both the study groups. On comparison between post-therapy systolic (Post-therapy: Yoga Group = 116.59 ± 1.45 mmHg and Waitlisted Controls = 119.68 ± 1.64 mmHg; p = 0.160) and diastolic blood pressures (Post-therapy: Yoga Group = 81.74 ± 1.20 mmHg and Waitlisted Controls = 83.18 ± 1.21 mmHg; p = 0.397) of yoga group and waitlisted controls, no significant difference was found between the arms. Systolic blood pressure of yoga group patients was found reduced significantly (Pre-therapy = 120.7 ± 1.84 mmHg and Post-therapy =116.59 ± 1.45; p = 0.025) after the intervention while we have not reported any change after standard therapy in waitlisted controls (Pre-therapy = 120.23 ± 1.66 mmHg and Post-therapy = 119.68 ± 1.64; p = 0.672). Yogic intervention as well as standard therapy had no effect on diastolic blood pressure of fibromyalgia patients (Yoga Group: Pre-therapy - 83.69 ± 1.27 mmHg and Post-therapy - 81.74 ± 1.20 mmHg; p = 0.174; Waitlisted Controls: Pre-therapy = 84.07 ± 1.18 mmHg and Post-therapy = 83.18 ± 1.21 mmHg; p = 0.384). (Figure 17a-b).

**Figure 17.**
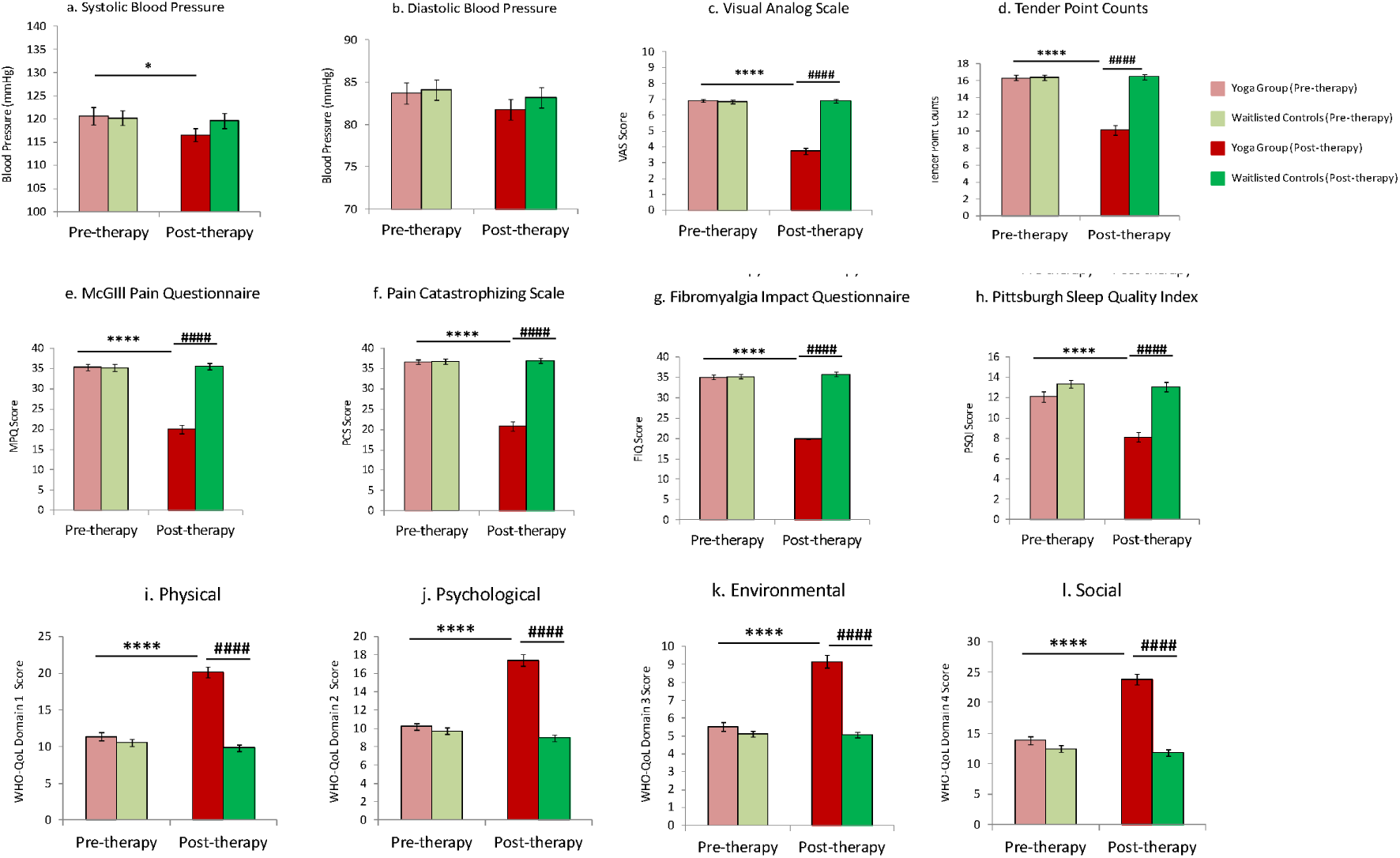
Effect of yogic intervention and standard therapy on a) Systolic Blood Pressure; b) Diastolic Blood Pressure; c) Visual Analog Scale; d) Tender Point Counts; e) McGill Pain Questionnaire; f) Pain Catastrophizing; g) Fibromyalgia Impact; h) Piitsburgh Sleep Quality Index in yoga group; i-l) WHO-Quality of Life: i) Physical; j) Psychological; k) Environmental, and l) Social domain, in yoga group and waitlisted controls of fibromyalgia patients respectively. Data was normally distributed (Gaussian distribution) – ‘Shapiro-Wilk test’; expressed in Mean ± SEM. Paired t-test was performed for intragroup comparison between pre-yoga and post-yoga; two sample (independent) t-test was performed for intergroup comparison between pre- and post-therapy changes. p-value < 0.05 was considered significant. Asterisk (*) depicts intragroup significant differences post therapy. “*” represents p<0.05; “****” represents p<0.0001. Hashtag (#) depicts an intergroup significant difference between pre- or post therapy. “####” represents p<0.0001.

### Subjective assessment of pain and associated symptoms of fibromyalgia of the randomized controlled trial (n = 120)

Intensity of pain experienced by fibromyalgia patients was measured using a numerical pain rating scale - Visual Analogue Scale (VAS) score. The VAS score of fibromyalgia patients of both the study groups were comparable at baseline (Yoga Group: Pre-yoga - 6.92 ± 0.12 and Waitlisted Controls: Pre-yoga - 6.85 ± 0.12; p = 0.683). Four weeks of yogic intervention and standard therapy in fibromyalgia patients showed a significant difference at post intervention time point (Yoga Group: Post-yoga - 3.73 ± 0.21 and Waitlisted Controls: Post-yoga - 6.90 ± 0.11; p<0.0001). Only yoga group showed significant improvement on the VAS (Yoga Group: Pre-therapy - 6.92 ± 0.12 and Post-therapy - 3.73 ± 0.21; p<0.0001; Waitlisted Controls: Pre-therapy = 6.85 ± 0.12 and Post-therapy = 6.90 ± 0.11; p = 0.582). (Table 5 & Figure 17c) Fibromyalgia patients of yoga group and waitlisted controls had a total of 16.33 ± 0.32 and 16.38 ± 0.32 tender points respectively at baseline, which was comparable (p = 903). Tender points in fibromyalgia patients of yoga group was found significantly lower in number than that of the waitlisted controls post-therapy (Yoga Group: Post-therapy = 10.15 ± 0.57 and Waitlisted Controls: Post-therapy = 16.45 ± 0.31; p<0.0001). On comparison of effect of both the therapy individually, we found a significant reduction in tender point counts after four weeks of yogic intervention (Pre-therapy - 16.33 ± 0.32 and Post-therapy – 10.15 ± 0.57; p<0.0001) whereas standard therapy (Pre-therapy = 16.38 ± 0.32 and Post-therapy = 16.45 ± 0.31; p = 0.376) didn’t show any effect on fibromyalgia patients of our cohort. (Table 5 & Figure 17d)

Subjective assessment of pain was performed using short form McGill Pain Questionnaire (SF-MPQ) and Pain Catastrophizing Scale (PCS). The MPQ scores of fibromyalgia patients of both the study groups were comparable at the baseline (Yoga Group = 35.31 ± 0.77; Waitlisted Controls = 35.23 ± 0.83; p = 0.945); which after four weeks of interventions differed significantly (Yoga Group = 20.06 ± 1.10; Waitlisted Controls = 35.50 ± 0.81; p<0.0001) from each other. Fibromyalgia patients of yoga group showed a significant improvement in pain score (Pre-therapy = 35.31 ± 0.77 and Post-therapy = 20.06 ± 1.10; p<0.0001) after regular yoga practice, while waitlisted patients taking standard medications (Pre-therapy = 35.23 ± 0.83 and Post-therapy = 35.50 ± 0.81; p = 0.556) effect remain unaffected (Table 5 & Figure 17e). Catastrophization due to pain in fibromyalgia (PCS score) were comparable initially in yoga group and waitlisted controls (Yoga Group: Pre-therapy = 36.62 ± 0.57; Waitlisted Controls: Pre-therapy = 36.80 ± 0.62; p = 0.834). PCS scores were found significantly lower in yoga group after four weeks of yogic intervention as compared to that in standard therapy (Yoga Group: Post-therapy = 20.84 ± 1.13; Waitlisted Controls: Pre-therapy = 36.93 ± 0.62; p<0.0001). Four weeks of yogic intervention significantly reduced catastrophizing pain of fibromyalgia (Pre-therapy = 36.62 ± 0.57 and Post-therapy = 20.84 ± 1.13; p<0.0001). Standard therapy had no significant role in pain relief from fibromyalgia pain (Pre-therapy = 36.80 ± 0.62 and Post-therapy = 36.93 ± 0.62; p = 767) when assessed using PCS questionnaire (Table 5 & Figure 17f).

Assessment of disease impact and quality of life of fibromyalgia patients were performed using Fibromyalgia Impact Questionnaire (FIQ) and WHO Quality of life (Brief) questionnaire (WHO-QOL-BREF) once at baseline and then after four weeks of interventions. We found a similar impact of fibromyalgia in both the study groups at baseline (Yoga Group: Pre-therapy = 35.05 ± 0.53; Waitlisted Controls: Pre-therapy = 35.23 ± 0.59; p = 0.819); but at four-week (post-therapy) impact was significantly lower in yoga group patients as compared to those of waitlisted controls (Yoga Group: Pre-therapy = 19.99 ± 1.11; Waitlisted Controls: Pre-therapy = 35.77 ± 0.58; p<0.0001). Lesser impact on patients due to fibromyalgia was observed post yoga (Pre-therapy = 35.05 ± 0.53 and Post-therapy = 19.99 ± 1.11; p<0.0001). Four weeks of standard therapy had no effect on fibromyalgia impact (Pre-therapy = 35.23 ± 0.59 and Post-therapy = 35.77 ± 0.58; p = 231) (Table 5 & Figure 17g).

Pittsburgh Sleep Quality Index (PSQI) measures sleep quality in patients suffering from impaired sleep due to psychological distress or chronic pain. Sleep quality in fibromyalgia were found impaired and were comparable between yoga group and waitlisted control patients (Yoga Group: Pre-therapy = 12.11 ± 0.49; Waitlisted Controls: Pre-therapy = 13.36 ± 0.40; p = 0.054). After four weeks of yogic intervention and standard therapy, two arms varied significantly (Yoga Group: Post-therapy = 8.14 ± 0.46; Waitlisted Controls: Pre-therapy = 13.07 ± 0.47; p<0.0001); patients practicing four weeks of supervised yoga had lower sleep impairment than others taking standard therapy. Yogic intervention significantly improves sleep (Pre-therapy = 12.11 ± 0.49 and Post-therapy = 8.14 ± 0.46; p<0.0001) in fibromyalgia patients while similar course of standard therapy had no significant (Pre-therapy = 13.36 ± 0.40 and Post-therapy = 13.07 ± 0.47; p = 312) role as such (Table 5 & Figure 17h).

Quality of life of fibromyalgia patients was assessed using WHO-QOL-BREF questionnaire for all the four domains. At baseline, measures of quality of life for all the domains such as physical (Yoga Group: Pre-therapy = 11.38 ± 0.54; Waitlisted Controls: Pre-therapy = 10.60 ± 0.50; p = 0. 0.293), psychological (Yoga Group: Pre-therapy = 10.02 ± 0.38; Waitlisted Controls: Pre-therapy = 9.72 ± 0.48; p = 0. 0.625), environmental (Yoga Group: Pre-therapy = 5.51 ± 0.24; Waitlisted Controls: Pre-therapy = 5.12 ± 0.16; p = 0. 0.177) and social (Yoga Group: Pre-therapy = 13.21 ± 0.63; Waitlisted Controls: Pre-therapy = 12.45 ± 0.50; p = 0. 0.347) were comparable between the study groups. These measures of quality of life showed a significantly higher scores after four weeks of interventions in the yoga group than the waitlisted controls: physical (Yoga Group: Post-therapy = 20.20 ± 0.71; Waitlisted Controls: Post-therapy = 9.83 ± 0.47; p<0.0001), psychological (Yoga Group: Post-therapy = 17.43 ± 0.65; Waitlisted Controls: Post-therapy = 8.95 ± 0.35; p<0.0001), environmental (Yoga Group: Post-therapy = 9.16 ± 0.35; Waitlisted Controls: Post-therapy = 5.08 ± 0.16; p<0.0001) and social (Yoga Group: Post-therapy = 23.85 ± 0.85; Waitlisted Controls: Post-therapy = 11.82 ± 0.52; p<0.0001). A significant improvement in quality of life was reported in yoga group after four weeks of regular supervised yogic intervention not in the patients undergoing four weeks of standard therapy in the waitlisted controls. (Table 5 & Figure 17i-l).

### Objective assessment of pain using quantitative sensory testing in fibromyalgia patients of the randomized controlled trial (n = 117)

Objective assessment of pain was performed using Quantitative Sensory Testing (QST) machine with pressure modality. Mechanical pressure was applied by the digital algometer device on the four dermatomes – three affected region (left shoulder, right shoulder and lower back) and one reference site (dorsum of hand) to assess the tenderness and associated pain sensitivity in fibromyalgia patients. Pressure pain threshold and pressure pain tolerances were measured at all the above-mentioned sites four times and averaged. Both the pressure pain threshold and pressure pain tolerance were higher at the reference sites as compared to the left and right shoulders at baseline (Pre-therapy) and post intervention (Post-therapy) in the study groups (Table 5).

Pressure pain threshold at dorsum of hand was comparable between yoga group and waitlisted control fibromyalgia patients at baseline (Yoga Group: Pre-therapy = 166.72 ± 6.37 kPa; Waitlisted Controls: Pre-therapy = 167.12 ± 6.20 kPa; p = 0.180) and post-therapy (Yoga Group: Post-therapy = 156.19 ± 6.63; Waitlisted Controls: Post-therapy = 154.91 ± 4.99 kPa; p = 0.876). There were no effect of four weeks of yogic intervention (Pre-therapy = 166.72 ± 6.37 kPa and Post-therapy = 156.19 ± 6.63 kPa; p = 0.443) and standard therapy (Pre-therapy = 167.12 ± 6.20 kPa and Post-therapy = 154.91 ± 4.99 kPa; p = 0.202) on pressure pain threshold at dorsum of hand of fibromyalgia patients (Table 5 & Figure 18a). Left shoulder pressure pain threshold was also comparable between yoga group and waitlisted control patients at baseline (Yoga Group: Pre-therapy = 130.82 ± 7.29 kPa; Waitlisted Controls: Pre-therapy = 143.06 ± 7.69 kPa; p = 0.180). Threshold at left shoulder post-therapy (Yoga Group: Post-therapy = 131.31 ± 8.28; Waitlisted Controls: Post-therapy = 137.29 ± 6.46 kPa; p = 0.703) did not showed any significant difference after four weeks between the study groups. There were no effect of four weeks of yogic intervention (Pre-therapy = 130.82 ± 7.29 kPa and Post-therapy = 141.31 ± 8.28 kPa; p = 0.064) as well as standard therapy (Pre-therapy = 143.06 ± 7.69 kPa and Post-therapy = 137.29 ± 6.46 kPa; p = 0.127) on pressure pain threshold at left shoulder of fibromyalgia patients when post-therapy versus baseline comparison was done. Although pressure pain threshold at left shoulder post yogic intervention showed a trend of increase, it didn’t reach the set significance level (Table 5 & Figure 18b). Pressure pain threshold at baseline at the right shoulder was comparable between yoga group and waitlisted control patients (Yoga Group: Pre-therapy = 130.7 ± 7.61 kPa; Waitlisted Controls: Pre-therapy = 137.96 ± 7.42 kPa; p = 0.351). Pain threshold at right shoulder post-therapy (Yoga Group: Post-therapy = 132.55 ± 8.16; Waitlisted Controls: Post-therapy = 124.78 ± 7.20 kPa; p = 0.476) did not showed any significant difference after four weeks between these two study groups. There were no effect of four weeks of yogic intervention (Pre-therapy = 130.7 ± 7.61 kPa and Post-therapy = 132.55 ± 8.16 kPa; p = 0.062) and standard therapy (Pre-therapy = 137.96 ± 7.42 kPa and Post-therapy = 124.78 ± 7.20 kPa; p = 0.059) on pressure pain threshold at right shoulder of fibromyalgia patients when post-therapy versus baseline comparison was done (Table 5 & Figure 18c). Lower back pressure pain threshold of yoga group patients at baseline was not significantly different from that of waitlisted control (Yoga Group: Pre-therapy = 177.04 ± 9.04 kPa; Waitlisted Controls: Pre-therapy = 184.04 ± 10.77 kPa; p = 0.738). Pressure pain threshold at lower back of fibromyalgia patients of yoga were not different from waitlisted controls post-therapy (Yoga Group: Post-therapy = 190.90 ± 7.02; Waitlisted Controls: Post-therapy = 174.84 ± 9.66 kPa; p = 0.181). We found a significant enhancement in the pain threshold after four weeks of yogic intervention (Pre-therapy = 177.04 ± 9.04 kPa and Post-therapy = 190.90 ± 7.02 kPa; p = 0.009). Waitlisted controls, on standard therapy (Pre-therapy = 184.04 ± 10.77 kPa and Post-therapy = 174.84 ± 9.66 kPa; p = 0.274) did not have any significant change in pressure pain threshold at lower back of fibromyalgia patients when post-therapy versus baseline comparison was done (Table 5 & Figure 18d).

**Figure 18.**
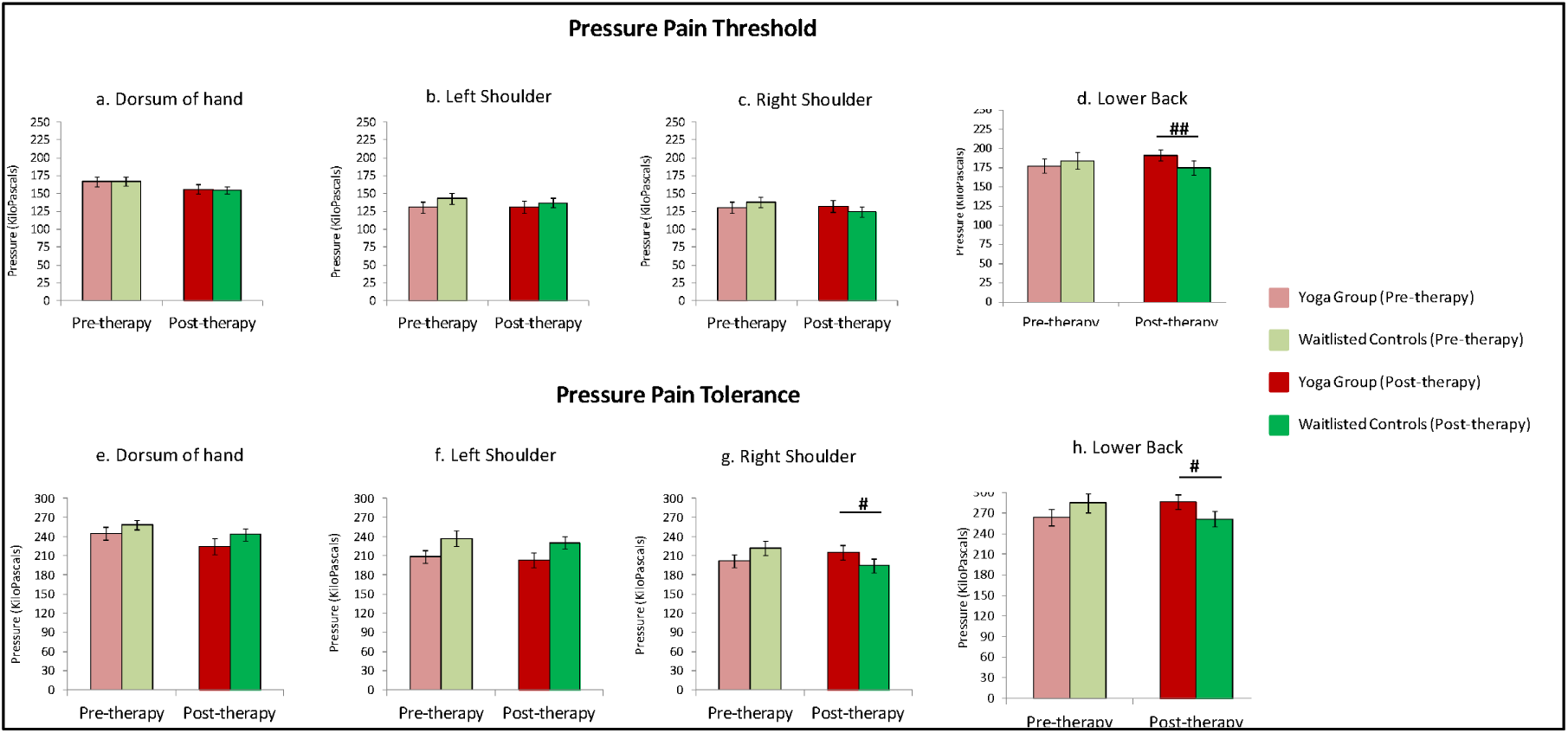
Quantitative Sensory Testing. Effect of yogic intervention and standard therapy on pressure pain threshold at: a) Dorsum of Hand; b) Left Shoulder; c) Right Shoulder, and d) Lower Back, in fibromyalgia patients of yoga group and waitlisted controls. And pressure pain tolerance at: e) Dorsum of Hand; f) Left Shoulder; g) Right Shoulder, and h) Lower Back, in fibromyalgia patients of yoga group and waitlisted controls Data was normally distributed (Gaussian distribution) – ‘Shapiro-Wilk test’; expressed in Mean ± SEM. Paired t-test was performed for intragroup comparison between pre-yoga and post-yoga; two sample (independent) t-test was performed for intergroup comparison between pre- and post-therapy changes. p-value < 0.05 was considered significant. Asterisk (*) depicts intragroup significant differences post therapy. “*” represents p<0.05; “****” represents p<0.0001. Hashtag (#) depicts an intergroup significant difference between pre- or post therapy. “#” represents p<0.05; “##” represents p<0.01; “####” represents p<0.0001.

Pressure pain tolerance at dorsum of hand was comparable between fibromyalgia patients of yoga group and waitlisted control at baseline (Yoga Group: Pre-therapy = 245.16 ± 10.07 kPa; Waitlisted Controls: Pre-therapy = 258.25 ± 7.76 kPa; p = 0.161) and post-therapy (Yoga Group: Post-therapy = 224.68 ± 12.10; Waitlisted Controls: Post-therapy = 242.83 ± 8.75 kPa; p = 0.586). There were no effect of four weeks of yogic intervention (Pre-therapy = 245.16 ± 10.07 kPa and Post-therapy = 224.68 ± 12.10 kPa; p = 0.844) and standard therapy (Pre-therapy = 258.25 ± 7.76 kPa and Post-therapy = 242.83 ± 8.75 kPa; p = 0.208) on pressure pain tolerance at dorsum of hand of fibromyalgia patients (Table 5 & Figure 18e). Left shoulder pressure pain tolerance was also comparable between yoga group and waitlisted control patients at baseline (Yoga Group: Pre-therapy = 208.52 ± 9.88 kPa; Waitlisted Controls: Pre-therapy = 237.34 ± 12.22 kPa; p = 0. 0.101). Pain tolerance at left shoulder post-therapy four weeks (Yoga Group: Post-therapy = 203.44 ± 11.83; Waitlisted Controls: Post-therapy = 230.37 ± 9.50 kPa; p = 0.840) did not showed any significant difference between the study groups. There were no effect of four weeks of yogic intervention (Pre-therapy = 208.52 ± 9.88 kPa and Post-therapy = 203.44 ± 11.83 kPa; p = 0.216) as well as standard therapy (Pre-therapy = 237.34 ± 12.22 kPa and Post-therapy = 230.37 ± 9.50 kPa; p = 0.100) on pressure pain tolerance at left shoulder of fibromyalgia patients when post-therapy versus baseline comparison was done (Table 5 & Figure 18f). Pressure pain tolerance at baseline at the right shoulder was comparable between yoga group and waitlisted control patients (Yoga Group: Pre-therapy = 201.59 ± 10.49 kPa; Waitlisted Controls: Pre-therapy = 222.03 ± 11.41 kPa; p = 0.113). Pain tolerance at right shoulder post-therapy (Yoga Group: Post-therapy = 215.41 ± 11.12; Waitlisted Controls: Post-therapy = 194.51 ± 10.77 kPa; p = 0.179) did not showed any significant difference after four weeks between these two study groups. There was a significant increase in the pain tolerance of fibromyalgia patients after four weeks of yogic intervention (Pre-therapy = 201.59 ± 10.49 kPa and Post-therapy = 215.41 ± 11.12 kPa; p = 0.037). Standard therapy (Pre-therapy = 222.03 ± 11.41 kPa and Post-therapy = 194.51 ± 10.77 kPa; p = 0.151) had no such effect on pressure pain tolerance at the right shoulder of fibromyalgia patients (Table 5 & Figure 18g). Lower back pressure pain tolerance of yoga group patients at baseline was not significantly different from that of waitlisted control (Yoga Group: Pre-therapy = 263.59 ± 11.76 kPa; Waitlisted Controls: Pre-therapy = 284.78 ± 14.01 kPa; p = 0.341). Pressure pain tolerance at lower back of fibromyalgia patients of yoga were not different from waitlisted controls post-therapy (Yoga Group: Post-therapy = 286.03 ± 10.85; Waitlisted Controls: Post-therapy = 261.56 ± 11.89 kPa; p = 0.131). There was a significant enhancement in the pain tolerance after four weeks of yogic intervention (Pre-therapy = 263.59 ± 11.76 kPa and Post-therapy = 286.03 ± 10.85 kPa; p = 0.014). Waitlisted controls, on standard therapy (Pre-therapy = 284.78 ± 14.01 kPa and Post-therapy = 261.56 ± 11.89 kPa; p = 0.492) did not have any significant change in pressure pain tolerance at lower back of fibromyalgia patients when post-therapy versus baseline comparison was done (Table 5 & Figure 18h).

### Assessment of flexibility and range of motion of fibromyalgia patients of the randomized controlled trial (n = 120)

Lumbar flexion and range of motion in patients with fibromyalgia (n = 120) were assessed using modified Schober’s test, Goniometry and Sit and Reach box before and after four weeks of yogic intervention and standard therapy in yoga group and waitlisted controls respectively (Table 5). Lateral goniometry was used to measure left-right flexion and rage of motion before and after yogic intervention and standard therapy in yoga group and waitlisted control fibromyalgia patients. Baseline measures of left goniometry were comparable between the two study groups (Yoga Group: Pre-therapy = 15.06 ± 0.48 ; Waitlisted Controls: Pre-therapy = 15.13 ± 0.54 ; p = 0.926). Post intervention, range of motion toward left in fibromyalgia patients of yoga group and waitlisted controls differed significantly (Yoga Group: Post-therapy = 21.87 ± 0.53 ; Waitlisted Controls: Post-therapy = 15.32 ± 0.47 ; p<0.0001) from the waitlisted controls; fibromyalgia patients undergoing four weeks of yogic intervention showed a significantly higher range of motion (Pre-therapy = 15.06 ± 0.48 and Post- therapy = 21.87 ± 0.53 ; p<0.0001). Left side range of motion of participants allocated for standard therapy for the same course in waitlisted controls arm (Pre-therapy = 15.13 ± 0.54 and Post-therapy = 15.32 ± 0.47 ; p = 0.754) didn’t significantly changed from baseline. Furthermore, range of motion toward right was also comparable between the groups (Yoga Group: Pre-therapy = 15.72 ± 0.50 ; Waitlisted Controls: Pre-therapy = 15.77 ± 0.58 ; p = 0.953). Post intervention, range of motion toward right in fibromyalgia patients of yoga group and waitlisted controls also differed significantly (Yoga Group: Post-therapy = 22.84 ± 0.52 ; Waitlisted Controls: Post-therapy = 16.10 ± 0.46 ; p<0.0001) from the waitlisted controls; fibromyalgia patients undergoing four weeks of yogic intervention showed a significantly higher range of motion (Pre-therapy = 15.72 ± 0.50 and Post- therapy = 22.84 ± 0.52 ; p<0.0001). Right side range of motion of participants allocated for standard therapy for the same course in waitlisted controls arm (Pre-therapy = 15.77 ± 0.58 and Post-therapy = 16.10 ± 0.46 ; p = 0.568) didn’t significantly changed from baseline (Table 5 & Figure 19a-b).

**Figure 19.**
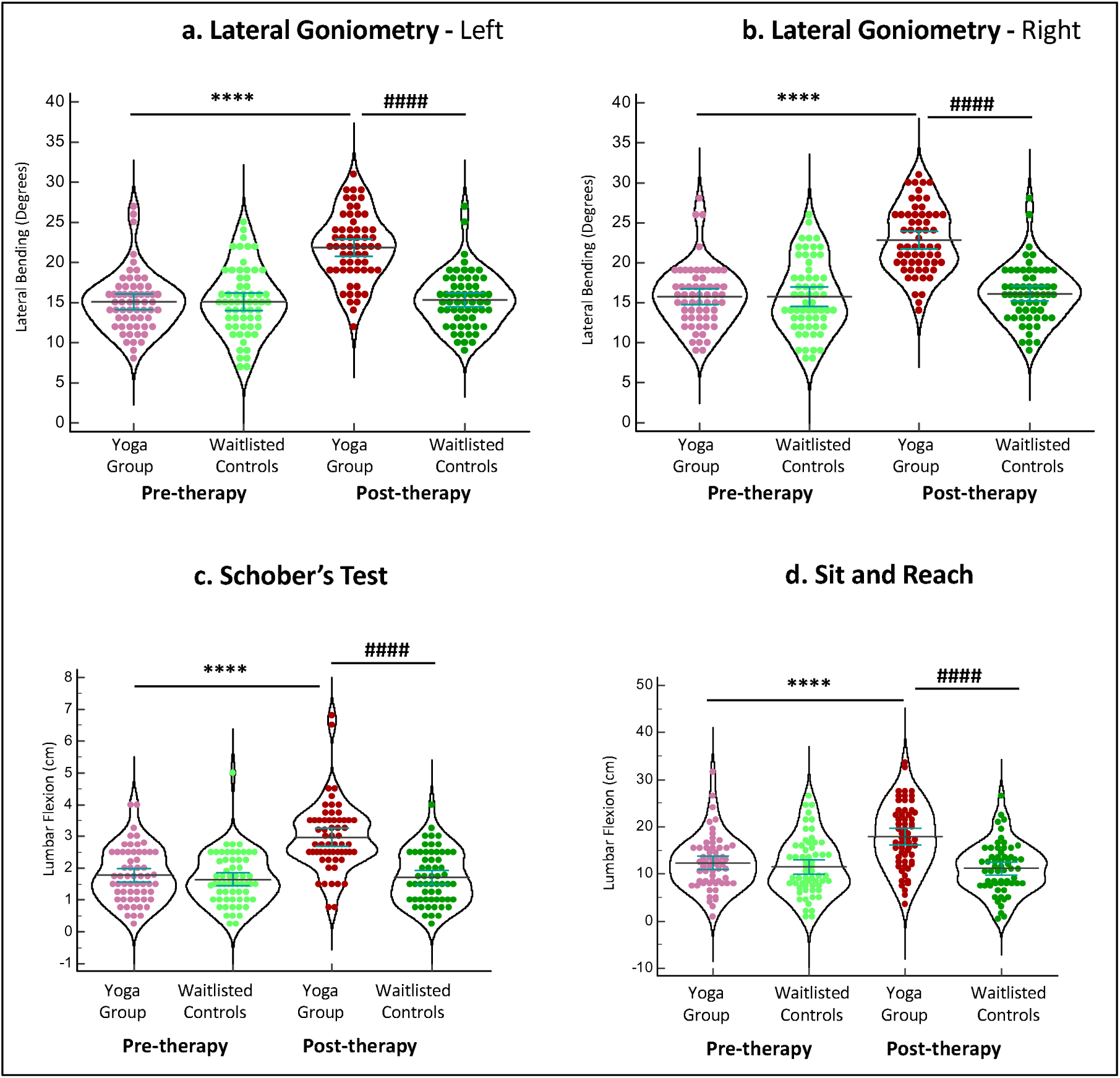
Effect of yogic intervention and standard therapy on range of motion: a) Left Goniometry; b) Right Goniometry; c) Schober’s Test; and d) Sit and Reach test, in fibromyalgia patients of yoga group and waitlisted controls. Data was normally distributed (Gaussian distribution) – ‘Shapiro-Wilk test’; expressed in Mean ± SEM. Paired t-test was performed for intra-group comparison between pre-yoga and post-yoga; two sample (independent) t-test was performed for intergroup comparison between pre- and post-therapy changes. p-value < 0.05 was considered significant. Asterisk (*) depicts intra-group significant differences post therapy. “****” represents p<0.0001. Hashtag (#) depicts an intergroup significant difference between pre- or post therapy. “#” represents p<0.05; “####” represents p<0.0001.

At baseline, lumbar flexibility and range of motion (measured using Schober’s test) in fibromyalgia patients of yoga group and waitlisted controls were comparable (Yoga Group: Pre-therapy = 1.76 ± 0.11 cm; Waitlisted Controls: Pre-therapy = 1.65 ± 0.11 cm; p = 0.461). After regular yoga practice, fibromyalgia patients of yoga group showed a significantly larger lumbar flexibility and range of motion than the waitlisted controls (Yoga Group: Post-therapy = 2.96 ± 0.14 cm; Waitlisted Controls: Post-therapy = 1.72 ± 0.11 cm; p<0.0001). Four weeks of yogic intervention in fibromyalgia patients significantly improved (Pre-therapy = 1.76 ± 0.11 cm and Post-therapy = 2.96 ± 0.14 cm; p<0.0001) lumbar flexibility and range of motion; standard therapy for the same course in waitlisted controls (Pre-therapy = 1.65 ± 0.11 cm and Post-therapy = 1.72 ± 0.11 cm; p = 0.426) didn’t have role in such flexibility enhancement (Table 5 & Figure 19c). Sit and reach box was used to measure lumbar flexibility and range of motion in fibromyalgia patients of yoga group and waitlisted controls. Baseline measures were comparable between the yoga group and waitlisted control patients (Yoga Group: Pre-therapy = 12.29 ± 0.72 cm; Waitlisted Controls: Pre-therapy = 11.48 ± 0.79 cm; p = 0.448). After regular yoga practice, fibromyalgia patients of yoga group showed a significantly larger lumbar flexibility and range of motion than the waitlisted controls (Yoga Group: Post-therapy = 17.86 ± 0.86 cm; Waitlisted Controls: Post-therapy = 11.13 ± 0.67 cm; p<0.0001). Four weeks of yogic intervention in fibromyalgia patients significantly improved (Pre-therapy = 12.29 ± 0.72 cm and Post-therapy = 17.86 ± 0.86 cm; p<0.0001) lumbar flexibility and range of motion; standard therapy for the same course in waitlisted controls (Pre-therapy = 11.48 ± 0.79 cm and Post-therapy = 11.13 ± 0.67 cm; p = 0.488) didn’t have role in such improvement in flexibility and range of motion (Table 5 & Figure 19d).

### Assessment of cortical excitability of fibromyalgia patients of the randomized controlled trial (n = 120)

Cortical excitability was measured using the Transcranial Magnetic Stimulation (TMS) machine; it was performed at primary motor cortex (M1) with the help of figure-of-8 coil. Resting Motor Threshold (RMT) is the measure of minimum percentage of MSO required to get a twitch of MEP amplitude ≥50 μV in half of the trial. Baseline measures of RMT were comparable between the fibromyalgia patients of yoga group and waitlisted controls (Yoga Group: Pre-therapy = 53.69 ± 0.95 % MSO; Waitlisted Controls: Pre-therapy = 52.19 ± 0.81 % MSO; p = 0.229). After regular yoga practice, fibromyalgia patients of yoga group showed a significantly lower RMT than that of the waitlisted controls (Yoga Group: Post-therapy = 48.91 ± 0.78 % MSO; Waitlisted Controls: Post-therapy = 52.67 ± 0.83 % MSO; p = 0.028). After four weeks of yogic intervention, RMT of the fibromyalgia patients significantly reduced (Pre-therapy = 53.69 ± 0.95 % MSO and Post-therapy = 48.91 ± 0.78 % MSO; p<0.0001) lumbar flexibility and range of motion; standard therapy for the same course in waitlisted controls (Pre-therapy = 52.19 ± 0.81 % MSO and Post-therapy = 52.67 ± 0.83 % MSO; p = 0.431) didn’t have role in such improvement in flexibility and range of motion (Table 5 & Figure 20a).

**Figure 20.**
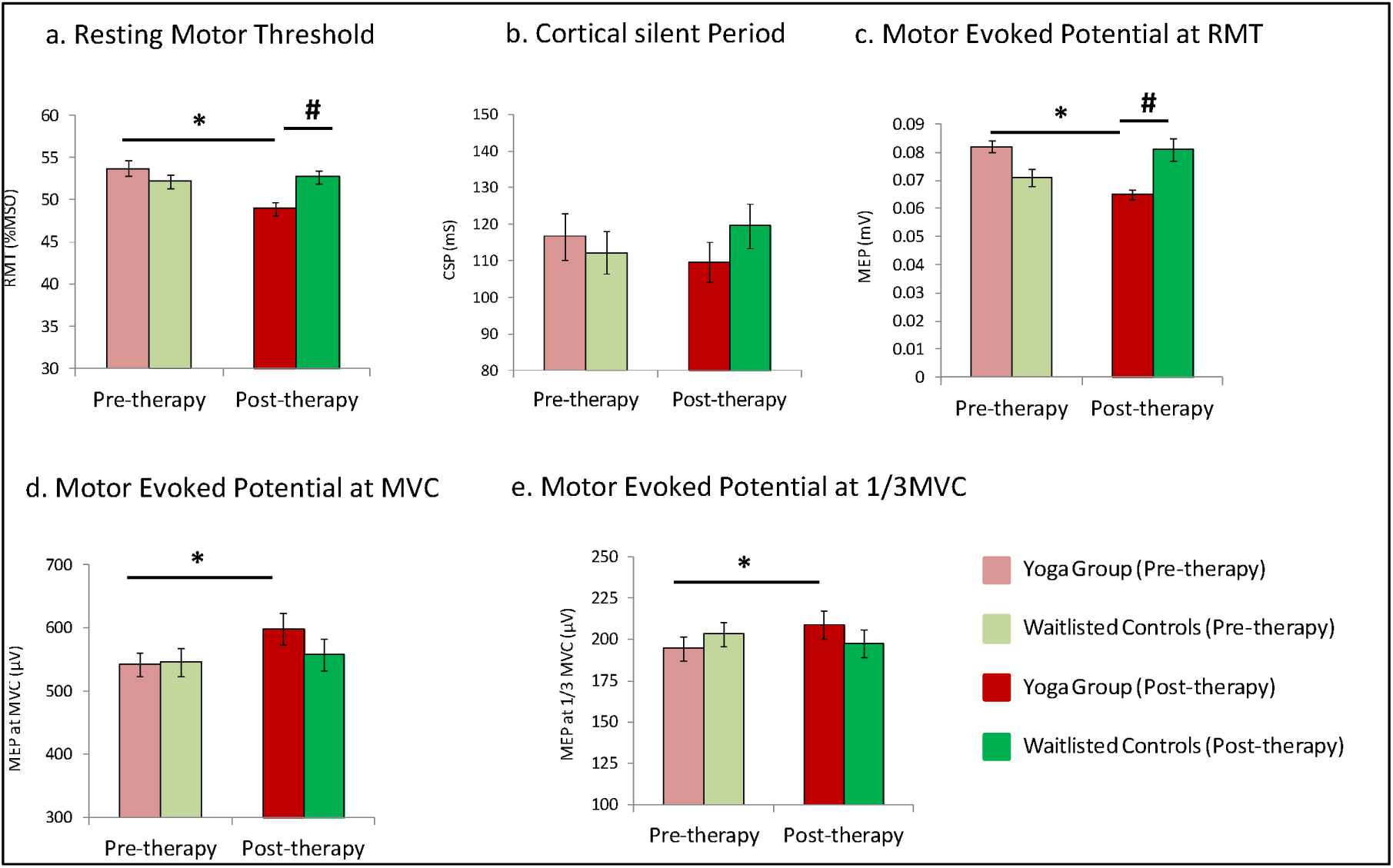
Effect of yogic intervention and standard therapy on cortical excitability parameters: a) Resting Motor Threshold; b) Cortical Silent Period; c) Motor Evoked Potential; d) Motor Evoked Potential at maximum voluntary contraction, and e) Motor Evoked Potential at 1/3 ‘ maximum voluntary contraction in fibromyalgia patients of yoga group and waitlisted controls. Data was normally distributed (Gaussian distribution) – ‘Shapiro-Wilk test’; expressed in Mean ± SEM. Paired t-test was performed for intragroup comparison between pre-yoga and post-yoga; two sample (independent) t-test was performed for intergroup comparison between pre- and post-therapy changes. p-value < 0.05 was considered significant. Asterisk (*) depicts intragroup significant differences post therapy. “*” represents p<0.05. Hashtag (#) depicts an intergroup significant difference between pre- or post-therapy. “#” represents p<0.05.

Corticomotor activity of the fibromyalgia patients were measured by recording peak-to-peak amplitude of the Motor Evoked Potential (MEP) at RMT, Maximum Voluntary Contraction (MVC) and one-third of the MVC in electromyographic response. At baseline MEP at RMT were significantly different between the fibromyalgia patients of yoga group and waitlisted controls (Yoga Group: Pre-therapy = 0.082 ± 0.002 mV; Waitlisted Controls: Pre-therapy = 0.071 ± 0.003 mV; p = 0.004*). Post-therapy, MEP of fibromyalgia patients of yoga group showed a significantly lower values than that of the waitlisted controls (Yoga Group: Post-therapy = 0.065 ± 0.002 mV; Waitlisted Controls: Post-therapy = 0.081 ± 0.004 mV; p = 0.016). After four weeks of yogic intervention, MEP of the fibromyalgia patients had significantly reduced from the baseline (Pre-therapy = 0.082 ± 0.002 mV and Post-therapy = 0.065 ± 0.002 mV; p = 0.012); standard therapy for the same course in waitlisted controls (Pre-therapy = 0.071 ± 0.003 mV and Post-therapy = 0.081 ± 0.004 mV; p = 0.243) didn’t have any significant effect on MEP (Table 5 & Figure 20b).

Cortical Silent Period (CSP) of the fibromyalgia patients were measured as the time period of interruption of ongoing muscle activity (generally one-third of MVC) by the Transcranial magnetic stimulation on M1. CSP were not significantly varied between the fibromyalgia patients of yoga group and waitlisted controls at baseline (Yoga Group: Pre-therapy = 116.70 ± 6.32 ms; Waitlisted Controls: Pre-therapy = 112.25 ± 5.69 ms; p = 0.602). We have not found any significant difference in CSP of fibromyalgia patients of yoga group and waitlisted controls post-therapy also (Yoga Group: Post-therapy = 109.60 ± 5.49 ms; Waitlisted Controls: Post-therapy = 119.59 ± 6.01 ms; p = 0.229). Neither four weeks of yogic intervention in yoga group (Pre-therapy = 116.70 ± 6.32 ms and Post-therapy = 109.60 ± 5.49 ms; p = 0.246) nor the standard therapy for same course in waitlisted controls (Pre-therapy = 112.25 ± 5.69 ms and Post-therapy = 119.59 ± 6.01 ms; p = 0.293) had any significant effect on CSP of fibromyalgia patients in our cohort (Table 5 & Figure 20c).

Motor Evoked Potential (MEP) at Maximum Voluntary Contraction (MVC) in fibromyalgia patients at baseline were comparable different fibromyalgia patients of yoga group and waitlisted controls (Yoga Group: Pre-therapy = 542.22 ± 19.62 μV; Waitlisted Controls: Pre-therapy = 546.51 ± 22.52 μV; p = 0.887). Post-therapy also, evoked potential at MVC in fibromyalgia patients of yoga group didn’t show significant difference from waitlisted controls (Yoga Group: Post-therapy = 598.40 ± 25.17 μV; Waitlisted Controls: Post-therapy = 557.76 ± 24.60 μV; p = 0.255). After four weeks of yogic intervention, MEP of the fibromyalgia patients had significantly increased from the baseline (Pre-therapy = 542.22 ± 19.62 μV and Post-therapy = 598.40 ± 25.17 μV; p = 0.05); standard therapy for the same course in waitlisted controls (Pre-therapy = 546.51 ± 22.52 μV and Post-therapy = 557.76 ± 24.60 μV; p = 0.776) didn’t have significant effect on MEP at MVC (Table 5 & Figure 20d).

Motor Evoked Potential (MEP) at one-third of Maximum Voluntary Contraction (MVC) in fibromyalgia patients at baseline were comparable between the fibromyalgia patients of yoga group and waitlisted controls (Yoga Group: Pre-therapy = 194.72 ± 7.35 μV; Waitlisted Controls: Pre-therapy = 203.42 ± 7.31 μV; p = 0.404). Post-therapy measures of MEP at MVC in fibromyalgia patients of yoga group was not different from that of waitlisted controls (Yoga Group: Post-therapy = 208.90 ± 8.46 μV; Waitlisted Controls: Post-therapy = 197.42 ± 8.52 μV; p = 0.347). After four weeks of yogic intervention, MEP at 1/3 MVC of the fibromyalgia patients had significantly increased from the baseline (Pre-therapy = 194.72 ± 7.35 μV and Post-therapy = 208.90 ± 8.46 μV; p = 0.043); standard therapy for the same course in waitlisted controls (Pre-therapy = 203.42 ± 7.31 μV and Post-therapy = 197.42 ± 8.52 μV; p = 0.732) had no significant effect on MEP at 1/3 MVC (Table 5 & Figure 20e).

A recruitment curve for the Motor Evoked Potential (MEP) and Cortical Silent Period (CSP) for the fibromyalgia patients of both the study groups were plotted from the respective individual data values, ranging 90 % to 150 % of the RMT with an escalation of 10 %, at baseline and after four weeks of yogic intervention and standard therapy in yoga group and waitlisted controls, respectively (Table 5). Average peak-to-peak amplitude of MEP at baseline in fibromyalgia patients of yoga group and waitlisted controls at 90% (Yoga Group: Pre-therapy = 0.037 ± 0.003 mV; Waitlisted Controls: Pre-therapy = 0.041 ± 0.004 mV; p = 0.462), 100 % (Yoga Group: Pre-therapy = 0.102 ± 0.013 mV; Waitlisted Controls: Pre-therapy = 0.106 ± 0.014 mV; p = 0.476), 110 % (Yoga Group: Pre-therapy = 0.350 ± 0.038 mV; Waitlisted Controls: Pre-therapy = 0.324 ± 0.041 mV; p = 0.625), 120 % (Yoga Group: Pre-therapy = 0.731 ± 0.068 mV; Waitlisted Controls: Pre-therapy = 0.581 ± 0.065 mV; p = 0.117), 130 % (Yoga Group: Pre-therapy = 1.12 ± 0.11 mV; Waitlisted Controls: Pre-therapy = 1.01 ± 0.11 mV; p = 0.509), 140 % (Yoga Group: Pre-therapy = 1.52 ± 0.15 mV; Waitlisted Controls: Pre-therapy = 1.42 ± 0.16 mV; p = 0.625) and 150 % (Yoga Group: Pre-therapy = 2.51 ± 0.32 mV; Waitlisted Controls: Pre-therapy = 2.10 ± 0.23 mV; p = 0.308) of the RMT were comparable. MEP at 90 % (Yoga Group: Post-therapy = 0.036 ± 0.001 mV; Waitlisted Controls: Post-therapy = 0.031 ± 0.002 mV; p = 0.023), 100 % (Yoga Group: Post-therapy = 0.055 ± 0.004 mV; Waitlisted Controls: Post-therapy = 0.087 ± 0.004 mV; p = 0.027), 110 % (Yoga Group: Post-therapy = 0.369 ± 0.036 mV; Waitlisted Controls: Post-therapy = 0.324 ± 0.033 mV; p = 0.024) and 120 % (Yoga Group: Post-therapy = 0.653 ± 0.060 mV; Waitlisted Controls: Post-therapy = 0.605 ± 0.063 mV; p = 0.029) of RMT at four weeks of interventions were significantly different between the study groups, whereas at 130 % (Yoga Group: Post-therapy = 0.972 ± 0.15 mV; Waitlisted Controls: Post-therapy = 0.977 ± 0.11 mV; p = 0.976), 140 % (Yoga Group: Post-therapy = 1.46 ± 0.19 mV; Waitlisted Controls: Post-therapy = 1.32 ± 0.14 mV; p = 0.554) and 150 (Yoga Group: Post-therapy = 2.10 ± 0.31 mV; Waitlisted Controls: Post-therapy = 2.06 ± 0.25 mV; p = 0.723) of RMT it was not different from each other (Table 5 & Figure 21a).

**Figure 21.**
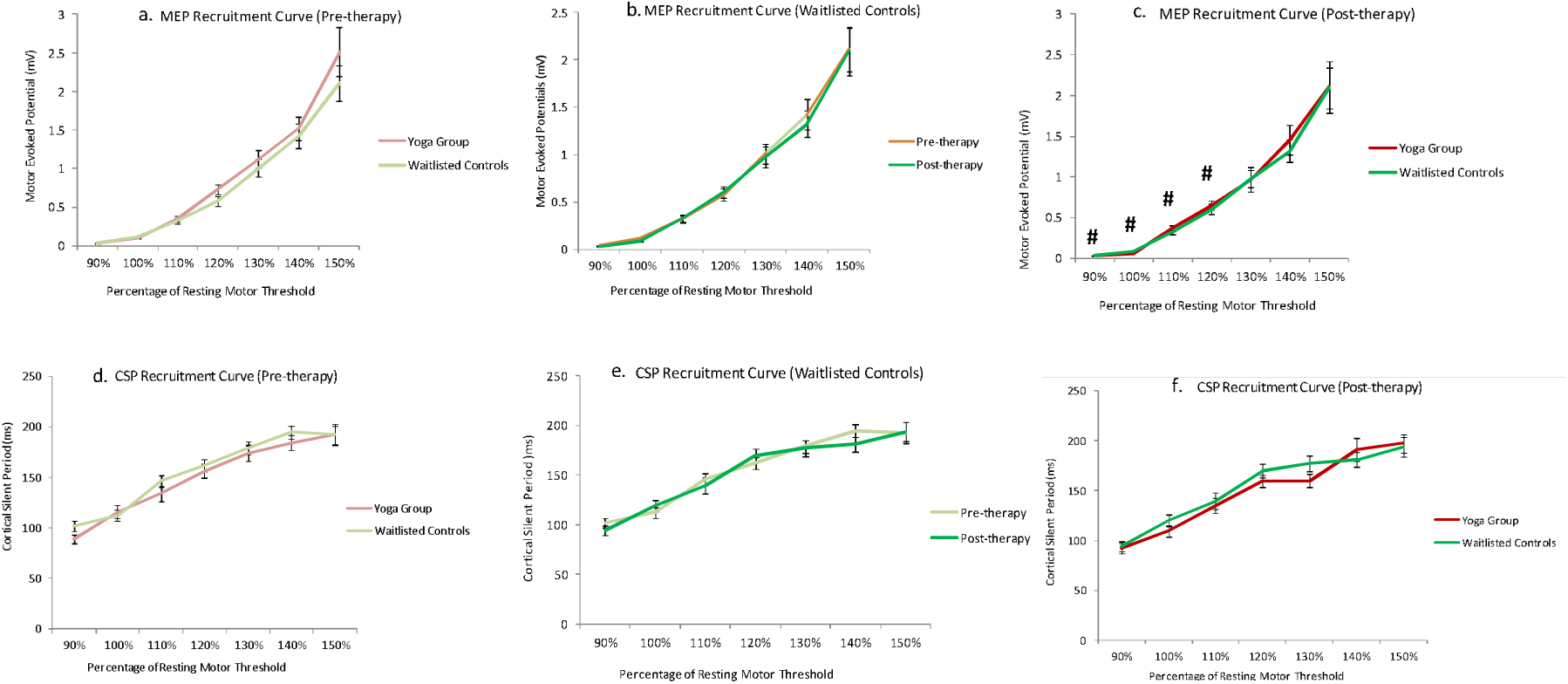
Recruitment Curves: Motor Evoked Potential (MEP) - a) Pre-therapy; b) Waitlisted Controls – pre- and post-therapy changes; c) Post-therapy. Cortical Silent Period (CSP) - d) Pre-therapy; e) Waitlisted Controls – pre- and post-therapy changes; f) Post-therapy, in fibromyalgia patients of yoga group and waitlisted controls. Data was normally distributed (Gaussian distribution) – ‘Shapiro-Wilk test’; expressed in Mean ± SEM. Paired t-test was performed for intragroup comparison between pre-yoga and post-yoga; two sample (independent) t-test was performed for intergroup comparison between pre- and post-therapy changes. p-value < 0.05 was considered significant. Asterisk (*) depicts intragroup significant difference post therapy. “*” represents p<0.05; “****” represents p<0.0001. Hashtag (#) depicts an intergroup significant difference between pre- or post-therapy. “#” represents p<0.05.

After four weeks of yogic intervention, MEP of the fibromyalgia patients had changed significantly at 110 % (Pre-therapy = 0.350 ± 0.038 mV and Post-therapy = 0.369 ± 0.036 mV; p = 0.021) and 120 % (Pre-therapy = 0.731 ± 0.068 mV and Post-therapy = 0.653 ± 0.060 mV; p = 0.020) of the RMT from the baseline while MEP at 90 % (Pre-therapy = 0.037 ± 0.003 mV and Post-therapy = 0.036 ± 0.001 mV; p = 0.827), 100 % (Pre-therapy = 0.102 ± 0.013 mV and Post-therapy = 0.055 ± 0.004 mV; p = 0.082), 130 % (Pre-therapy = 1.12 ± 0.11 mV and Post-therapy = 0.972 ± 0.15 mV; p = 0.280), 140% (Pre-therapy = 1.52 ± 0.15 mV and Post-therapy = 1.46 ± 0.19 mV; p = 0.580) and 150 % (Pre-therapy = 2.51 ± 0.32 mV and Post-therapy = 2.10 ± 0.31 mV; p = 0.332) of RMT it remain unchanged (Table 5 & Figure 21b). Standard therapy for the same course in waitlisted controls didn’t have any significant effect on MEP at 90 % (Pre-therapy = 0.041 ± 0.004 mV and Post-therapy = 0.031 ± 0.002 mV; p = 0.089), 100 % (Pre-therapy = 0.106 ± 0.014 mV and Post-therapy = 0.087 ± 0.004 mV; p = 0.667), 110 % (Pre-therapy = 0.324 ± 0.041 mV and Post-therapy = 0.324 ± 0.033 mV; p = 0.875), 120 % (Pre-therapy = 0.581 ± 0.065 mV and Post-therapy = 0.605 ± 0.063 mV; p = 0.451), 130 % (Pre-therapy = 1.01 ± 0.11 mV and Post-therapy = 0.977 ± 0.11 mV; p = 0.776), 140 % (Pre-therapy = 1.42 ± 0.16 mV and Post-therapy = 1.32 ± 0.14 mV; p = 0.327) and 150 % (Pre-therapy = 2.10 ± 0.23 mV and Post-therapy = 2.06 ± 0.25 mV; p = 0.998) of the RMT. (Table 5 & Figure 21c)

We have not observed any significant changes in comparison between baseline and post intervention data for yoga group as well as waitlisted controls. Cortical Silent Periods (CSPs) at baseline in fibromyalgia patients of yoga group and waitlisted controls at 90% (Yoga Group: Pre-therapy = 88.81 ± 4.44 ms; Waitlisted Controls: Pre-therapy = 102.05 ± 4.97 ms; p = 0.051), 100 % (Yoga Group: Pre-therapy = 116.70 ± 6.32 ms; Waitlisted Controls: Pre-therapy = 112.25 ± 5.69 ms; p = 0.602), 110 % (Yoga Group: Pre-therapy = 133.99 ± 7.41 ms; Waitlisted Controls: Pre-therapy = 146.06 ± 5.81 ms; p = 0.198), 120 % (Yoga Group: Pre-therapy = 156.17 ± 6.82 ms; Waitlisted Controls: Pre-therapy = 161.74 ± 5.85 ms; p = 0.534), 130 % (Yoga Group: Pre-therapy = 174.09 ± 7.89 ms; Waitlisted Controls: Pre-therapy = 178.92 ± 6.57 ms; p = 0.637), 140 % (Yoga Group: Pre-therapy = 184.21 ± 7.14 ms; Waitlisted Controls: Pre-therapy = 194.89 ± 6.66 ms; p = 0.276) and 150 % (Yoga Group: Pre-therapy = 192.04 ± 9.31 ms; Waitlisted Controls: Pre-therapy = 192.50 ± 10.54 ms; p = 0.974) of RMT were all comparable (Table 5 & Figure 21d).

Post-therapy CSP of yoga group and waitlisted controls patients at 90 % (Yoga Group: Post-therapy = 92.43 ± 4.95 ms; Waitlisted Controls: Post-therapy = 94.34 ± 4.82 ms; p = 0.785), 100 % (Yoga Group: Post-therapy = 109.60 ± 5.49 ms; Waitlisted Controls: Post-therapy = 119.59 ± 6.01 ms; p = 0.229), 110 % (Yoga Group: Post-therapy = 135.18 ± 7.01 ms; Waitlisted Controls: Post-therapy = 139.82 ± 8.03 ms; p = 0.670 ), 120 % (Yoga Group: Post-therapy = 159.84 ± 6.31 ms; Waitlisted Controls: Post-therapy = 169.51 ± 6.81 ms; p = 0.305), 130 % (Yoga Group: Post-therapy = 160.12 ± 6.87 ms; Waitlisted Controls: Post-therapy = 177.41 ± 7.80 ms; p = 0.103), 140 % (Yoga Group: Post-therapy = 191.19 ± 11.24 ms; Waitlisted Controls: Post-therapy = 181.22 ± 7.48 ms; p = 0.452) and 150 (Yoga Group: Post-therapy = 197.25 ± 9.47 ms; Waitlisted Controls: Post-therapy = 193.86 9.69 ms; p = 0.803) of RMT after four weeks of interventions, were not significantly different from each other (Table 5 & Figure 21e). After four weeks of yogic intervention, CSPs of the fibromyalgia patients of yoga group did not changed at 90 % (Pre-therapy = 88.81 ± 4.44 ms and Post-therapy = 92.43 ± 4.95 ms; p = 0.701), 100 % (Pre-therapy = 116.70 ± 6.32 ms and Post-therapy = 109.60 ± 5.49 ms; p = 0.246), 110 % (Pre-therapy = 133.99 ± 7.41 ms and Post-therapy = 135.18 ± 7.01 ms; p = 0.828), 120 % (Pre-therapy = 156.17 ± 6.82 ms and Post-therapy = 159.84 ± 6.31 ms; p = 0.513), 130 % (Pre-therapy = 174.09 ± 7.89 ms and Post-therapy = 160.12 ± 6.87 ms; p = 0.474), 140% (Pre-therapy = 184.21 ± 7.14 ms and Post-therapy = 191.19 ± 11.24 ms; p = 0.383) and 150 % (Pre-therapy = 192.04 ± 9.31 ms and Post-therapy = 197.25 ± 9.47 ms; p = 0.911) of RMT; remain unchanged for all datasets (Table 5 & Figure 21e). Standard therapy for the same course in waitlisted controls didn’t have any significant effect on CSP at 90 % (Pre-therapy = 102.05 ± 4.97 ms and Post-therapy = 94.34 ± 4.82 ms; p = 0.244), 100 % (Pre-therapy = 112.25 ± 5.69 ms and Post-therapy = 119.59 ± 6.01 ms; p = 0.293), 110 % (Pre-therapy = 146.06 ± 5.81 ms and Post-therapy = 139.82 ± 8.03 ms; p = 0.496), 120 % (Pre-therapy = 161.74 ± 5.85 ms and Post-therapy = 169.51 ± 6.81 ms; p = 0.394), 130 % (Pre-therapy = 178.92 ± 6.57 ms and Post-therapy = 177.41 ± 7.80 ms; p = 0.776), 140 % (Pre-therapy = 194.89 ± 6.66 ms and Post-therapy = 181.22 ± 7.48 ms; p = 0.211) and 150 % (Pre-therapy = 192.50 ± 10.54 ms and Post-therapy = 193.86 9.69 ms; p = 0.766) of the RMT values (Table 5 & Figure 21f).

### Assessment of blood biomarkers in fibromyalgia patients of the randomized controlled trial (n = 103)

Level of β-Endorphins in yoga group patients at baseline was comparable to that of waitlisted control (Yoga Group: Pre-therapy = 634.50 ± 39.29 pg/mL; Waitlisted Controls: Pre-therapy = 723.38 ± 36.59 pg/mL; p = 0.103). Concentration of β-Endorphins in fibromyalgia patients of yoga group were not different from that in waitlisted controls even post-therapy (Yoga Group: Post-therapy = 710.43 ± 39.61 pg/mL; Waitlisted Controls: Post-therapy = 619.18 ± 42.78 pg/mL; p = 0.121). We have not found any significant difference in the levels of β-Endorphins either after four weeks of yogic intervention in yoga group (Pre-therapy = 634.50 ± 39.29 pg/mL and Post-therapy = 710.43 ± 39.61 pg/mL; p = 0.145) or standard therapy in waitlisted controls (Pre-therapy = 723.38 ± 36.59 pg/mL and Post-therapy = 619.18 ± 42.78 pg/mL; p = 0.235) when post-therapy versus pre-therapy concentrations were compared (Table 5 & Figure 22a). Concentrations of cortisol in yoga group patients at baseline was comparable to that of waitlisted control (Yoga Group: Pre-therapy = 98.21 ± 6.04 ng/mL; Waitlisted Controls: Pre-therapy = 96.79 ± 6.79 ng/mL; p = 0.875). Concentration of cortisol in fibromyalgia patients of yoga group were not different from that in waitlisted controls even post-therapy (Yoga Group: Post-therapy = 108.54 ± 9.22 ng/mL; Waitlisted Controls: Post-therapy = 88.63 ± 6.22 ng/mL; p = 0.088). We have not found any significant difference in the levels of cortisol either after four weeks of yogic intervention in yoga group (Pre-therapy = 98.21 ± 6.04 ng/mL and Post-therapy = 108.54 ± 9.22 ng/mL; p = 0.157) or standard therapy in waitlisted controls (Pre-therapy = 96.79 ± 6.79 ng/mL and Post-therapy = 88.63 ± 6.22 ng/mL; p = 0.208) when post-therapy versus pre-therapy concentrations were compared (Table 5 & Figure 22b). Glutamate concentration in the serum of yoga group patients and waitlisted controls were comparable at baseline (Yoga Group: Pre-therapy = 244.86 (185.50, 434.42) μg/mL; Waitlisted Controls: Pre-therapy = 237.67 (167.96, 279.87) μg/mL; p = 0.189). Concentration of glutamate in fibromyalgia patients of yoga group were significantly lower from waitlisted controls at post-therapy (Yoga Group: Post-therapy = 206.49 (163.09, 309.27) μg/mL; Waitlisted Controls: Post-therapy = 246.07 (200.76, 431.11) μg/mL; p = 0.034). We have not found any significant difference in the levels of glutamate either after four weeks of yogic intervention in yoga group (Pre-therapy = 244.86 (185.50, 434.42) μg/mL and Post-therapy = 206.49 (163.09, 309.27) μg/mL; p = 0.605) or standard therapy in waitlisted controls (Pre-therapy = 237.67 (167.96, 279.87) μg/mL and Post-therapy = 246.07 (200.76, 431.11) μg/mL; p = 0.871) when post-therapy versus pre-therapy concentrations were compared (Table 5 & Figure 22c). Level of serotonin in the serum of yoga group patients and waitlisted controls were also comparable at baseline (Yoga Group: Pre-therapy = 35.79 (23.70, 63.49) ng/mL; Waitlisted Controls: Pre-therapy = 38.55 (23.43, 85.30) ng/mL; p = 0.837). Concentration of serotonin in fibromyalgia patients of yoga group were not different from that of waitlisted controls at post-therapy (Yoga Group: Post-therapy = 38.88 (29.84, 64.55) ng/mL; Waitlisted Controls: Post-therapy = 31.97 (21.66, 57.02) ng/mL; p = 0.148). We have not found any significant difference in the levels of Serotonin either after four weeks of yogic intervention in yoga group (Pre-therapy = 35.79 (23.70, 63.49) ng/mL and Post-therapy = 38.88 (29.84, 64.55); p = 0.388) or standard therapy in waitlisted controls (Pre-therapy = 38.55 (23.43, 85.30) ng/mL and Post-therapy = 31.97 (21.66, 57.02) ng/mL; p = 0.402) when post-therapy versus pre-therapy concentrations were compared (Table 5 & Figure 22d). Concentration of Substance P in the serum of yoga group patients and waitlisted controls were comparable at baseline (Yoga Group: Pre-therapy = 72.94 (20.90, 149.07) pg/mL; Waitlisted Controls: Pre-therapy = 73.19 (14.50, 145.55) pg/mL; p = 0.180). Concentration of Substance P in fibromyalgia patients of yoga group were not significantly different from waitlisted controls at post-therapy (Yoga Group: Post-therapy = 40.23 (12.54, 107.21) pg/mL; Waitlisted Controls: Post-therapy = 50.47 (16.28, 113.38) pg/mL; p = 0.536). We have not found any significant difference in the levels of Substance P either after four weeks of yogic intervention in yoga group (Pre-therapy = 72.94 (20.90, 149.07) pg/mL and Post-therapy = 40.23 (12.54, 107.21) pg/mL; p = 0.097) or standard therapy in waitlisted controls (Pre-therapy = 73.19 (14.50, 145.55) pg/mL and Post-therapy = 50.47 (16.28, 113.38) pg/mL; p = 0.199) when post-therapy versus pre-therapy concentrations were compared (Table 5 & Figure 22e).

**Figure 22.**
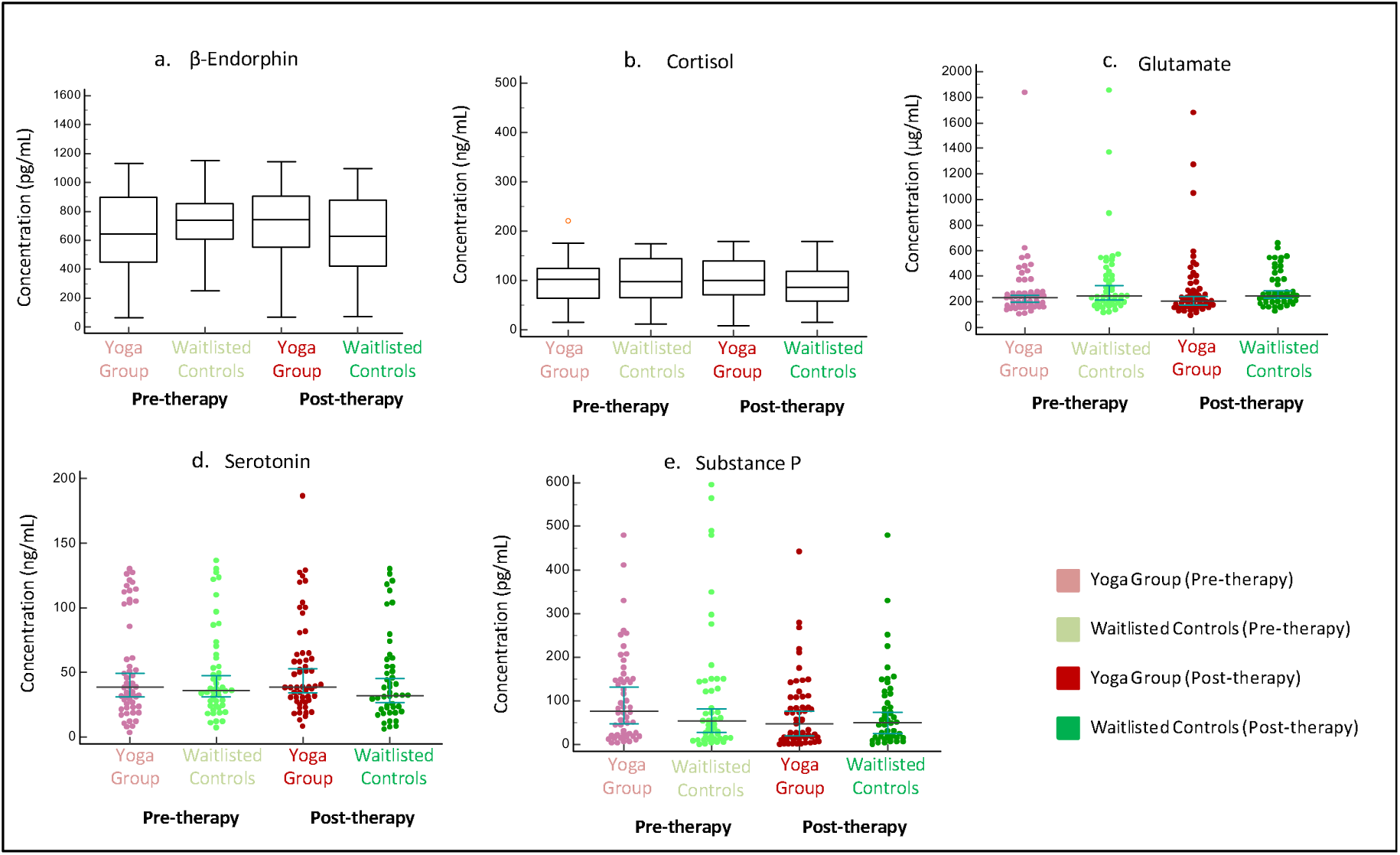
Comparison of levels of pain and related biomarkers in serum: a) β-Endorphin; b) Cortisol; c) Glutamate; d) Serotonin, and e) Substance P in fibromyalgia patients of yoga group and waitlisted controls. Data for β-Endorphin and serotonin were normally distributed (Gaussian distribution) – ‘Shapiro-Wilk test’; expressed in Mean ± SEM. Glutamate, Serotonin and Substance P data were non-parametric – ‘Shapiro-Wilk test’; expressed in Median (Q1, Q3). Paired t-test and Signed rank sum test (Mann-Whitney U test) was performed for comparison between pre-yoga and post-yoga assessment for parametric data and non-parametric data, respectively Data was normally distributed (Gaussian distribution) – ‘Shapiro-Wilk test’; expressed in Mean ± SEM. Paired t-test was performed for intragroup comparison between pre-yoga and post-yoga; two sample (independent) t-test was performed for intergroup comparison between pre- and post-therapy changes. p-value < 0.05 was considered significant. Asterisk (*) depicts intragroup significant differences post therapy.

## Discussion

Fibromyalgia is a psychosomatic disorder, reported predominantly in women, with symptoms such as severe widespread musculoskeletal pain along with morning stiffness, fatigue, anxiety, depression, brain fogging and sleep problems. [2, 35] Most of the patients in our cohort of fibromyalgia patients were middle-aged, non-working women bearing low socioeconomic status with a history of post- partum pain onset (one-fifth patients); females affected in a ratio of 1:5 (Male: Female); shoulder, neck and lower back were the main regions for onset of pain. Evidences from previous literatures favours that disease is present in around one-fifth of the males also, as found in our cohort; although no relevant gender differences in the clinical picture of fibromyalgia were observed on analysing 1023 patients (138 males) and the assumption of well-established gender differences could not be supported. [36] Though, fibromyalgia is considered to be predominantly prevalent in females and are more likely to be diagnosed with psychiatric disorders and Connective Tissue Disease (CTD), while males are more likely to be diagnosed with medical conditions. [37] Till date exact pathophysiology of the disease is not clear; although literatures describe important events influencing the patient such as aberrant pain processing, muscle micro-trauma, impact on neuroendocrine metabolism may lead to development of fibromyalgia syndrome. In our previous studies, we found that fibromyalgia is a chronic pain condition which is characterized by considerable depression, poor coping, higher cortical excitability and catastrophizing pain with sleep problems in over half of the Indian cohort of patients. [21, 23, 25]

Yoga is culturally believed to stand out from the other pain treatment strategies due to its ability to favorably influence sensorimotor functions, attention, attitudes towards incoming painful stimuli and perceptual control over the pain experience. [38] Robust evidence already exists that documents the role of yogic intervention in the management of other chronic pain conditions. [26–28, 39] However, the mechanism of action of yoga induced analgesia is not known. Historical subjective evidences suggest role of yogic training in inducing a quiescence-like mental state towards painful stimuli by altering the attitudes and beliefs towards painful stimuli. [40–42] The hypothesis presents a nascent line of scientific inquiry that has pointed to the effect of expectation of pain relief as an indelible part of yogic intervention, even if partially. Although several corroborating literatures claim the same, this aspect has rarely been formally and objectively investigated. The present study was an attempt to extensively document the beneficial effects of four weeks of regular and supervised yogic intervention on pain status, quality of life, flexibility and range of motion along with cortical excitability in fibromyalgia patients, when compared to same course of standard therapy. Therefore, we aimed to address the neurophysiological basis of pain modulation with both subjective and objective measures and the interdialogue between quality of life, sleep, cortical excitability and flexibility and range of motion reporting in Indian cohort of fibromyalgia patients in the hope to fill up the current lacuna in the field. The objectives were designed first to understand the symptomatology (case control study), then explore (interventional study), and finally validate and compare (randomized controlled trial) various outcomes of fibromyalgia using yoga as a major lifestyle intervention and standard treatment module as a comparator.

### Fibromyalgia pain is devastating with multimodal impact – Evidence from Case Control Study

Fibromyalgia patients of Indian cohort were middle-aged (mean age = 36.68 years), mostly low socioeconomic, non-working women; which corroborates with an Asian study by Prateepavanich et al. 2017, witnessing the same. [43] According to our results on socioeconomic data, we concluded that fibromyalgia patients have a higher illness when their education and socio-economic status are in poor condition, and the complications of pain and illness are further higher in patients having lower socioeconomic status. In the Brazilian population (n=768) authors reported higher prevalence of fibromyalgia in low socioeconomic population. [44] Homemakers and married women were more prone/affected in our cohort of fibromyalgia patients than the others having professional involvements and risk of getting disease further increases post-partum. Around 73% of the patients visiting our clinics were married, bearing mean children of 2.07; one-sixth of them complaint pain onset was postpartum and that after their first/subsequent delivery of the baby. Fibromyalgia syndrome is common among pregnant females; sex hormones and reproductive outcomes may or may not be related with the fibromyalgia [45]. Postpartum onset of the disease is still not clear but a few literature shows that there could be a potential relationship between the conception and fibromyalgia. [45–47] Our study also revealed that the factors affecting fibromyalgia patients are multidimensional arising from potential risk factors to lifestyles involving genetic as well as allogenic factors. Most of the physiological parameters like weight, body mass index (BMI), blood pressure and the heart rate of fibromyalgia patients were not much different from the normal range data of healthy individuals. Studies show that the fibromyalgia patients had more heart rate variability aberrances and indices of increased sympathetic activity, indicating that they are more prone to cardiovascular risks. [48] Morning stiffness is seen in one-fourth of the patients where stiffness lasted from 15 minutes to a couple of hours; Wolfe et al. marked morning stiffness and fatigue as the key symptoms of fibromyalgia syndrome. [35, 49] Factors affecting stress and stress response systems such as loss of sense of effort, economic difficulty, poor quality of life, and many more also render the disease onset. [50] More than one-fifth of the fibromyalgia patients were having genetic history of any rheumatic conditions in their near lineage in either maternal or paternal side. Among various exolred pathophysiological factors; some genes are found to be involved in fibromyalgia such as SLC64A4, TRPV2, MYT1L, and NRXN3; most of them are transport channel genes. [12] Catechol-O-methyltransferase Val158Met polymorphism modulated some psychological variables but not pressure pain sensitivity in fibromyalgia syndrome, because women carrying the Met/Met genotype showed higher disability, depression, and anxiety but similar pressure pain thresholds than those with Val/Met or Val/Val genotypes. [51]

In the present study, pain profiles of the fibromyalgia patients were investigated using VAS scores, tender point counts and duration of pain. Mean VAS score of the fibromyalgia patients was found to be 6.92 ± 0.12 which shows that patients had excruciating pain which can be the most vital reason for poor quality of life, catastrophization and triggering the psychological abnormalities. The VAS score observed in our study was found to be compatible with the previous studies where fibromyalgia patients were reported to experience higher pain than other pain syndromes. [52] In our study, average duration of fibromyalgia pain was found 7.17 ± 1.98 years; fibromyalgia is a chronic pain syndrome with clinical stages of chronicity which may last lifetime with continuous progression in the VAS score and psychological manifestations. [53] Furthermore, it is very important to know the region of onset of fibromyalgia pain; we found that around one-third of the patients in our cohort of fibromyalgia patients were having the first pain felt at either shoulder or neck, or the lower back. Excruciating pain of fibromyalgia is widespread over specified tender points; tenderness at the peculiar tender points is due to allodynia and hyperalgesia associated with peripheral and central sensitization of the patients due to intense nociception and an increased sensitivity in central pain modulation mechanisms (central sensitisation) which results in ‘sensitivity to touch or tenderness’. [54–55] We have found that out of 19 specific tender points for fibromyalgia, on an average 16.33 ± 0.32 tender points were present in the patients. Most of the tender points were seen on the dorsal surface of the patients from the trapezius muscle, knees, lower back and over the gluteus maximus. [56] Most of the tender points reported in our study were bilaterally present on the body surface from neck to toe in the patients with excruciating pain and tenderness. [57–58] On critical agometric analysis of tender points, authors found that these specific points are the hallmark in dignosing fibromyalgia due to associated disruption in the sensory functions [56].

Recently a fMRI study have also shown evidence of functional differences in fibromyalgia within brain regions associated with motivational aspects of pain, as well as differences in neural activity related to nociceptive regulation, arousal, and autonomic homeostatic regulation within the brainstem and spinal cord regions. [59] Fibromyalgia patients generally have a poor sleep and quality of life as compared to healthy controls [60]; it is also evidenced in 43% patients in our study. Sleep problems like lower sleep quality and sleep efficiency; longer wake time after sleep onset, short sleep duration, and light sleep when objectively assessed and more difficulty in initiating sleep were found when the sleep characteristics were subjectively assessed in fibromyalgia patients. [61] Few studies have shown that most of the inhibitory neurotransmitters that regulate circadian rhythm and enhance acute stress responses and pain such as Serotonin, Dopamine and Noradrenaline were low and several peptides of the endogenous opioid system were higher to modulate the pain in fibromyalgia. [62–64] Levels of β- Endorphin and serotonin were also quite higher in healthy controls whereas stress and pain related biomarkers were higher in studied fibromyalgia patients. Endorphins are the hallmark of social, mental and physical well-being; Bidari and colleagues recruited 30 fibromyalgia patients in a study and found lower serum β-endorphin levels than healthy controls. [65] Furthermore, levels of substance P which inhibits basal tone and acute responses of the HPA axis; and excitatory neurotransmitters like glutamate were found to be elevated in the blood of the fibromyalgia patients. [62–66] Similarly, the opioids, which are increased in fibromyalgia, inhibit acute sympathetic and HPA axis stress responses [67]. Reduced pain inhibition (due to hyperlagesia and allodynia) in combination with the increased input of pain signals are considered to cause central sensitization and tenderness at the specific sites in the patients. [53] Pain matrix is a complex circuit in the brain which integrates emotional, social and physical insults of pain to impose nociception on different functional brain areas in fibromyalgia patients. [68] Central sensitization is quite common and is sometimes considered as the key mechanism for fibromyalgia symptomatology. [69] The resultant hyperalgesia and allodynia in the patients is ultimately manifested as tenderness at the affected sites. [54] This could result in lower pressure pain parameters (both threshold and tolerance) at the shoulders and lower back of fibromyalgia patients than that of healthy individuals; whereas it was comparable at reference sites. The pressure pain thresholds and the pressure pain tolerances on both muscle and bone were lower in the fibromyalgia patients than in the healthy controls. [70]

Poor Lumbar flexion of fibromyalgia patients was also evidenced in our fibromyalgia cohort; it is in line with our previous findings. [39] However, there is inadequate evidence till date on the cortical parameters of the fibromyalgia patients; we have a novelty in introducing and correlating pain, musculoskeletal performance and cortical parameters and establishing control over pain. Higher cortical excitability in the fibromyalgia patients have been evidenced in the previous literature also which is possibly due to significant cortical plasticity due to chronic pain. [71]

### Role of yogic intervention in improving sensory function (including pain status) in fibromyalgia - Evidence from Interventional Study and Randomized Controlled Trial

Fibromyalgia syndrome is characterized by the involvement of both higher centre and peripheral sensitization. [54] It is a novel study where an attempt to establish a trilateral dialogue between pain, musculoskeletal function and corticomotor excitability in fibromyalgia patients within multiple study design and experimental set-up. Moreover, primary attention were rendered to objective assessment parameters to unwind the role of yogic intervention at peripheral as well as central level together. Yoga is psychosomatic cost effective and easy-to-adopt practice in day to day life. Yoga enhances parasympathetic nervous system activity and has neuroprotective effects due to serotonin, a pain biomarker. [72] Recently, it has been proven beneficial in relieving pain for various chronic pain conditions such as chronic low back pain, rheumatoid arthritis, chronic neck pain, migraine, chronic fatigue syndrome and many more. [26–27, 73] In our previous study by Arya et al., same yoga regimen was devised for chronic low back pain and reported improvement in pain and related disorders before and after 4 weeks of regular intervention [27]. Since, few tender points in fibromyalgia patients coincide with that of chronic low back pain patients; hence, we proposed same pre-validated yoga program to our cases at the Integral Health and Wellness Clinic. Almost all the patients were having same handedness to avoid errors in the cortical excitability measures using TMS coil. Accountability of the attendance for each patient enrolled in the yoga program was kept stringent in the physical attendance sheet; data for the participants attending less than 90% of the sessions were not included in the study. Travel allowances were not provided to any of the participants for attending yoga classes.

A significant reduction in the systolic blood pressure after four weeks of yogic intervention was observed in fibromyalgia patients while diastolic blood pressure remained unchanged after intervention. Also heart rate variability (HRV) was significantly lower in the fibromyalgia patients than the controls. [74] Blood pressure related pain inhibitory dysfunction in fibromyagia in has been evidenced earlier also. Fibromyalgia pain displayed the expected BP-related inhibitory effects in the clinical settings. [75] There are direct evidences for the betterment of systolic BP after 1-month yoga practice. [76] When twenty sessions of supervised yogic intervention were administered to fibromyalgia patients (n = 60) having a high VAS score of 6.92 ± 0.12; a significant reduction in the VAS score, tender point counts and PCS score of the fibromyalgia patients after the yoga therapy were observed corroborating with other studies. [40–42, 77–78] Pain catastrophization and impact of fibromyalgia correlate with poor quality of life in patients and disturbed sleep too. [40, 42]. Fibromyalgia Impact and quality of life of fibromyalgia patients for all the domains such as physical, psychological, environmental and social, have improved by regular yoga practicing. Also sleep quality improved significantly [79]. Moreover, using questionnaires, we found reduction in the pain scores of MPQ and PCS questionnaire after yoga. On subjective assessment of pain we found significant relief and improvement in pain status in the yoga group as compared to the waitlisted controls. Fibromyalgia impact was also found lesser in fibromyalgia patients who have completed four weeks of supervised yoga than those on standard therapy. Female fibromyalgia patients enrolled in an RCT of weekly yoga program for 8 weeks, a significant improvement in the symptom and functioning, including pain, fatigue, and mood, and in pain catastrophizing, acceptance, and other coping strategies were reported; pointing toward a promising scope for the potential benefits of yoga for women suffering from fibromyalgia. [76, 79] Their follow-up results also showed that patients sustained most of their post therapy benefits, with the fibromyalgia impact score in remaining one-third of the patients improved at the end of 3 months. [41] Fibromyalgia impact score significantly correlates with physical functioning, physical role, and bodily pain in fibromyalgia patients (n = 30). [80] Patients of fibromyalgia made more errors, had significantly increased reaction time for cognitive tasks, marked daytime sleepiness, and impaired quality of sleep [81]; also sleep was found related to depression through pain and physical functioning in the patients with fibromyalgia. [82] Gentle Hatha yoga reduced fibromyalgia-related symptoms in a sample of 10 adult patients who received yoga therapy twice a week for 8 weeks. [83] An uncontrolled pilot study with 36 completers, authors reported psychosocial functioning and a range of symptoms using pre-validated tools before and after a Satyananda yoga, which included weekly in-person sessions for 6 weeks and daily home-based yoga video practice of the same postures; suggests that yoga may reduce pain and catastrophizing, as well as improve sleep; besides physical sessions home yoga practice imparted better impact and considered as supplementary doses. [42] Although there are ample randomized controlled trials available in the literature validating different forms of yoga for efficacy amongst fibromyalgia patients, most of them are subjective studies and guarantee benefits to a shorter extent only. For the robust statement to be derived from the yoga-based trials, authors suggest an organized objective trial in the larger cohort of fibromyalgia patients should be warranted, which can have a wide coverage of symptomatic relief. Therefore, we explored the impact of yogic intervention on fibromyalgia and associated symptoms using objective tools.

Yoga may also exert its effects through physiological mechanisms, as it impacts the hypothalamic-pituitary-adrenal (HPA) axis. Specifically, morning levels of cortisol increased in both depressed patients who participated in a yoga intervention and in yoga practitioners when compared to controls [84] while other studies did not reported any change in cortisol level post-yoga intervention. [85–87] Moreover, using an intention to treat analysis, total cortisol output increased significantly post-intervention. [40] Cortisol impacts many different physiological systems (e.g., immunity, metabolism) and plays a role in augmenting the activity of the autonomic nervous system, such as enhancing the sympathetically mediated cardiovascular response to stress (e.g., increased heart rate). [88] Aerobic and physical exercises also stimulate the hypothalamus causing increased secretion of endorphins in fibromyalgia patients, and this effect helps to attenuate pain sensation and improve mood state and sleep. [89–90, 65] β-endorphins are endogenous opioids, an increased level of which also suggests a significant improvement in mind-body communication indicators following yoga practice. [91] A non randomized control trial in which a 12-week yoga program was conducted in chronic low back pain patients for stress and inflammatory factors evaluation in the serum, cortisol level and other factors were found significantly reduced in the yoga group post therapy. [92] Changes in mindfulness and cortisol levels were also evident in an 8-week long yoga program executed in women with fibromyalgia for 75 minutes twice a week. Cortisol is a stress marker which can negatively affect fibromyalgia patients by pain catastrophization and poor quality of life was explored solely in this paper. In a magnetic resonance spectroscopic study performed in fibromyalgia patients and healthy controls, patients showed higher levels of glutamate/glutamine (*Glx*) compounds; but there is no direct evidence of alteration of glutamate level in fibromyalgia patient after yogic intervention. [93] An increased interstitial concentration of glutamate was found in women with fibromyalgia which was normalized after an exercise therapy in a case-control study. [94–95] A significant difference in the glutamate level was also reported in serum of fibromyalgia patients after yogic intervention and standard therapy. Other blood biomarkers remain unchanged after the interventions.

There is a paucity of literature for the effect of yoga on objective parameters of pain in fibromyalgia. Pressure pain threshold insights scope for the quantification of hyperalgesia and allodynia due to tenderness at affected sites in fibromyalgia. [96] Objective assessment of pain showed significantly higher pressure pain thresholds at shoulder and lower back area after yogic intervention while standard therapy had no effect. Moreover, pressure pain parameters were comparable at reference sites, post intervention. Yoga therapy can improve both the pressure pain parameters at affected sites in the patients like other pain conditions. [28, 39] We reported that four week long yoga program is able to modulate pressure pain threshold and pressure pain tolerance at shoulder and L5 area of fibromyalgia patients, which has also been reported by other studies as well, with other therapeutic approach. [97–98] On the other hand standard therapy is unable to improve the tenderness at affected sites. Quantitative sensory testing using pressure modality serves as a remedy for approximately one-fifth of the patients. [39] We selected the three most painful areas common in fibromyalgia cases to vanish site-specific biases and one painless area which was the dorsum of the hand. [97] Pressure pain parameters in fibromyalgia have been continuously devised to assess pain sensitivity and associated tenderness before and after treatment strategies. In our previous interventional as well as case studies we found an increased pressure pain threshold at the left shoulder and pressure pain tolerance at the right shoulder after 20 sessions and 40 sessions of regular and supervised yoga therapy. [28, 39] Although, we have not found any significant change in the concentration of pain biomarkers such as serotonin and substance P after yoga practice in fibromyalgia patients, glutamate levels between the two arms of the RCT showed a significant difference post-therapy. Yoga intervention has a significant influence on serum serotonin levels in patients with chronic low back pain but its effect on fibromyalgia patients is missing from the literature. [99–100] Physical as well as resistant exercises act as nature’s tranquilizer by helping to boost serotonin in the brain. Substance P is one of the neurotransmitters involved in pain, depression, anxiety and in cognition as well. [101] Although massage therapy and relaxation therapy for 5 weeks have shown to reduce substance P and fibromyalgia pain in 24 fibromyalgia patients [102]; there is a lack of literature showing evaluation of substance P after administering yogic intervention in fibromyalgia patients. Higher RMT and reduced suprathreshold MEP in fibromyalgia patients bilaterally, suggest a global decline in corticospinal excitability not correlated with clinical features. Patients with fibromyalgia also had lower ICF and SICI than controls and were correlated with fatigue, pain catastrophizing and depression. Objective and quantifiable changes in brain function in fibromyalgia is associated with deficits in intracortical modulation involving both GABAergic and glutamatergic pathways, possibly related to certain aspects of the pathophysiology. [71] Glutamate plays a major role in pain neurotransmission, with receptors found in the sensory part of the periphery, the spinal column (Aδ and C fibers), and in the pain processing areas of the brain, such as the anterior cingulate cortex (ACC), insular cortex (IC), primary somatosensory cortex (S1), and many more. [88]

### Role of yogic intervention in improving motor functions (including corticomotor excitability) in fibromyalgia - Evidence from Interventional Study and Randomized Controlled Trial

Yoga postures were designed to stretch muscles throughout the body, which can significantly harness overall flexibility of fibromyalgia patients experiencing muscular rigidity, resistant to movement and stiffness. Authors reported the beneficial effects of short-term intensive medical yoga therapy programs on spinal flexibility in low back pain patients with a higher effect size (1.305). [103] We assessed flexibility and range of motion parameters using Schober’s test, goniometer and sit and reach box, which have not been used earlier in yoga-based or any therapeutic study in fibromyalgia patients. A significant improvement in flexibility and range of motion was reported after four weeks of yogic intervention as compared to standard therapy in fibromyalgia patients. In a case report of adult established case of fibromyalgia, where yoga postures were designed specifically to improve flexibility and movement of joints, daily 1 hour, 6 days/week in the morning and evening for 9 months, patient demonstrated reduction in muscle fatigue and improvement in quality of life and sleep. [104] Yoga not only modulates our minds but also increases the musculoskeletal activity of patients. Objective evidence that medical yoga therapy relieves back pain and improves lumbar flexibility than standard care, was recently explored in our previous study. [27] Although, there is a paucity of evidence on improvement of musculoskeletal performance in fibromyalgia patients using yogic intervention. Yoga impart a beneficial effect in improving overall mobility by gently stretching muscles and promoting body awareness using modulation of flexibility and range of motion in fibromyalgia patients, ultimately resulting in reduced pain and stiffness associated with the disease. [28, 39] We devised the similar yoga regimen administered by Arya et al. for our cohort of patients also, keeping in mind the mimic of few tender points in both the cohorts of chronic pain conditions. The excruciating pain of fibromyalgia is the lead cause of movement-related disability in fibromyalgia patients. [65] Yoga is supposed to modulate our brain areas and is supposed to enhance the musculoskeletal strength and thereby functional activity of the patients with chronic pain. [28, 39] Fibromyalgia pain interferes with lumbar flexibility and range of motion might be due to brain morphometric changes especially motor cortex due to chronic and excruciating pain at tender points; other associated areas are also found affected in some neuroimaging studies. [68, 105] Standard therapy available to fibromyalgia patients on the other hand was less effective in restoring the depressed flexion and related musculoskeletal functions. This is the first study where we have assessed flexibility and range of motion in a RCT setting in fibromyalgia patients. Supplemetary Table 7 lists out all the muscles involved in practicing yoga asasas.

Chronic pain (including fibromyalgia pain) leads to cortical plasticity in the brain due to aberration in the pain pathway and corticomotor axis through patients’ continuous inhibition of the movement of painful areas of the body. [38] Yoga improves corticomotor excitability of fibromyalgia patients by modulating pain inhibition pathways [21, 65]. Cortical activity, specifically RMT, MEPs and MEP recruitment curves, showed a significant improvement in yoga group as compared to waitlisted controls in the present study. Most of these parameters of cortical excitability such as, RMT, MEP and recruitment curve showed significant improvement after other therapeutic interventions as well in fibromyalgia patients [25]. Although there is evidence of assessment of corticomotor excitability in fibromyalgia patients after other therapeutic interventions; none of the study till date has recorded corticomotor excitability before and after the yogic intervention in fibromyalgia patients. This is the first and foremost study to assess cortical changes using yogic intervention in fibromyalgia patients; though, one pilot study was conducted by Govindraj and team in 2018 to elucidate the effects of yoga therapy on brain excitability in healthy individuals by examining the changes in short interval intracortical inhibition (SICI) and cortical silent period (CSP) – two distinct TMS-derived measures of cortical inhibition that are mediated by GABA_A_ and GABA_B_ sub-receptors, respectively after medical yoga therapy training in healthy individuals. [38] Chronic pain in fibromyalgia results in inhibition of the movement of affected areas which results in cortical plasticity in the brain areas and patients show higher resting motor threshold and lower motor evoked potentials due to psychosomatic and functional depression [25, 65]. The study reported significant enhancement of MEP, SICI nd CSP duration also elongated in healthy individuals following 1 month of yoga therapy [38]. TMS was applied on the hand motor areas of both the hemispheres and motor evoked potentials (MEPs) were recorded in female fibromyalgia patients and age-matched healthy controls using single-pulse stimulation for the measurements of the resting motor threshold (RMT) and suprathreshold MEP; paired-pulse stimulation to assess short intracortical inhibition (SICI) and intracortical facilitation (ICF); putative correlations were sought between changes in electrophysiological parameters and some vital clinical features of fibromyalgia, such as pain, fatigue, anxiety, depression and catastrophizing. [65]

Although, cortical excitability parameters have been extensively explored in fibromyalgia patients [21, 25]; none of the study had investigated the role of yoga on it; although, case reports reveal that there is an improvement in cortical plasticity in fibromyalgia with supervised and regular four and eight weeks of yoga practice. In a case series, reduction in resting motor threshold by 2% of the maximum stimulus output was found by our team; while other corticomotor excitability parameters showed some changes or remain unchanged after eight weeks of supervised yogic intervention indicating the beneficial effect of yoga in fibromyalgia patients. [39] At least 4 weeks of regular and supervised yogic intervention can harness pain relief, flexibility, and improve corticomotor excitability in fibromyalgia. [39] We have also found a higher cortical excitability of our patients. RMT and MEP are supposed to be the result of a dialogue between the motor cortex and the muscles and plasticity in the cortical areas lead to lesser recruitment of muscle fibers and thus reduction in the silent period. [25] An increase in the amplitude of maximum voluntary contraction after yoga corroborate with earlier findings in the case report; also, decrease in the slope of the motor evoked potential recruitment curve and an elongation of the cortical silent period from baseline were documented following four weeks of yogic intervention. [25] Although, corticomotor excitability measures in the current study showed alterations in the motor threshold and silent period parameters, the evidence is not sufficient to completely establish and retrieve a robust conclusive hypothesis. Figure 23 describes the objective behind each test and proposed mechanism of symptomatic relief after four weeks of supervised yogic intervention in fibromyalgia patients.

**Figure 23.**
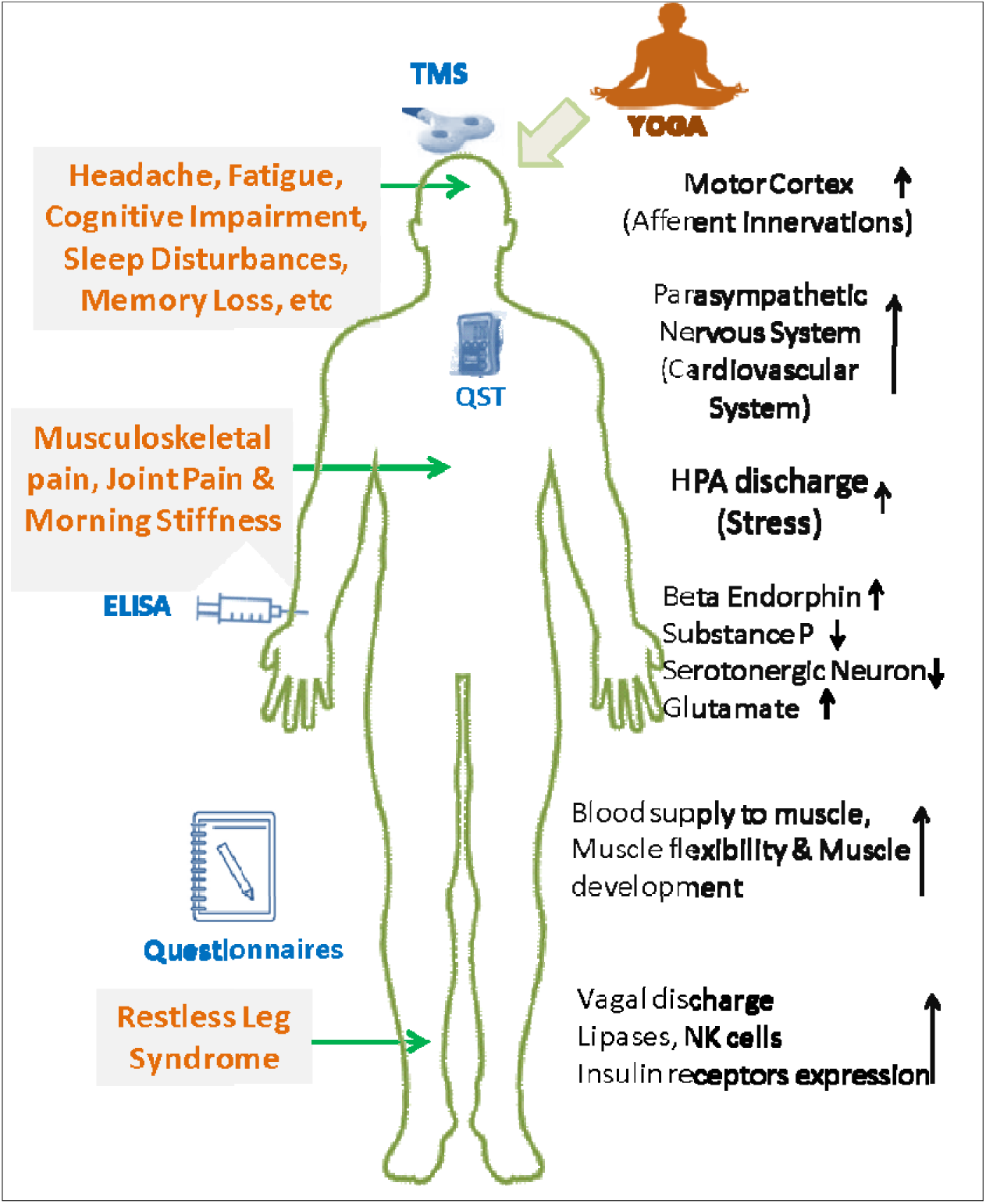
Proposed mechanism of action of yogic intervention in fibromyalgia.

To date, there is no permanent cure of fibromyalgia; patients have to completely rely on standard FDA approved medications which can reduce symptoms up to an extent and for a certain period of time. Repetitive transcranial magnetic stimulation technique has been also validated in our previous study. [5, 21] Physical therapy as well as medical yoga therapy is among the non-invasive lifestyle interventions which can be easily adopted in day-to-life without clinical assistance and do not require any additional equipment; it is also proven effective in various chronic pain conditions such as chronic low back pain, chronic fatigue syndrome including fibromyalgia. [22, 26–28, 106–107] Although yogic intervention has been validated as an effective treatment modality for fibromyalgia patients in the clinical set up in some literatures, but the objective evidence of symptomatic attenuation following it is missing in the literatures. [40–42, 83] Gentle Hatha yoga twice a week has been earlier subjectively proven to improve quality of life and fibromyalgia impact by reducing pain and associated symptoms in 10 patients. [83] We have also found reduction in tender point counts and catastrophization due to pain, which is in line with some recent literature on validation of weekly yoga sessions followed by daily video practice at home. [78, 83, 96] Yoga is a non-invasive and cost effective practice proven to manage various psychological distresses [108]. Rheumatologists and other fibromyalgia experts recommend a multidisciplinary and multimodal approach to symptom management for persons diagnosed with fibromyalgia. [109–110] Complementary mindfulness-based movement practices such as yoga shows a promise as a part of this multimodal approach. [111] Supplemetary Table 7 lists out all the asanas and respective muscle groups targeted during its practice.

To the best of our knowledge, this is the first Randomized Controlled Trial where objective evidence on pain relief and corticomotor excitability using yogic intervention in fibromyalgia patients have been explored. Effect of yoga on fibromyalgia-driven flexibility and range of motion inhibition has also not been explored till date and is a totally novel area. This is the first study where interconnection between pain, cortical excitability, musculoskeletal performance and neurotransmitters level in the serum have been investigated. We have not examined patients beyond 50 years of age to avoid hormonal fluctuations and physiological biases. Our study has certain limitations also which include quantification of blood biomarkers of pain and associated symptoms could have been carried out in a larger population of fibromyalgia patients. More follow-ups can be added in the study to get robust objective evidence on the relief from pain and other symptoms. For robust and firm recommendation of yoga for fibromyalgia patients, objective assessment of pain including biomarker profiling along with real-time neuroimaging of brain, such as Electroencephalography (EEG) and functional Magnetic Resonance Imaging (fMRI) while practicing yoga can be planned. An elaborate electromyographic study of the tender points, and intra-cortical inhibition and facilitation can help in establishing link between peripheral and central control of pain and associated symptoms in fibromyalgia in a holistic way. Also, we have not addressed medication history, which could have revealed potential drug intake and tolerance due to its long-term dependence for transient pain relief; yoga as a lifestyle intervention does not impart any such detrimental physiological changes. Group activities are always better for patient compliance and prevention of premature discontinuation of the treatment trials. In this study 24 patients which is almost one-third of the enrolled patients have premature discontinuation form the intervention or attended less than 90% of the planned sessions. Non-working Indian women (homemakers) are mostly dependent on their husbands and also low socioeconomic may have rendered potential financial strain for communicating regularly for four weeks to the hospital. While validating efficacy of any therapeutic or lifestyle intervention in fibromyalgia patients, assessor should choose more than one objective tool (like nociceptive flexion reflex, thermodes, etc) to quantify pain, apart from pressure parameters which is a semi-objective recording. While most of the researchers prefer to choose pressure pain threshold as one of the primary objective measures, it can render significant variations as two cohorts of fibromyalgia patients behave differently toward pressure pain stimulus. [39] It may be possible to improve the clinical course and pain conditions of the disease by increasing the standards of education and quality of life of fibromyalgia patients. Patients visiting regular OPDs were much more inclined toward the ease of access of the clinics despite any geography based biases as seen in our study. Although, diagnostic criteria for fibromyalgia appear to be used as a vague guide by clinicians and patients, and allow for substantial diagnostic expansion of fibromyalgia. [112] Fibromyalgia patients were mostly referred from rheumatology, Physical Medicine and Rehabilitation and psychiatry clinics; orthopedics and medicine departments are other alternative options for recruitment of fibromyalgia patients in the research based studies. In this study we offered regular and supervised yoga sessions coupled with interactive rounds which helped in creating a comfortable, friendly and trusting environment to increase the recruitment rate and decrease premature discontinuation of the therapy. Also, while planning treatment strategies for fibromyalgia patients, sociodemographic characteristics cannot be ignored to have higher compliance.

### Conclusions and future perspectives

The study extensively concludes that four weeks of regular and supervised yogic intervention may ameliorate pain, improve flexibility and range of motion and changes cortical plasticity in the Indian cohort of fibromyalgia patients, as compared to standard therapy. Yoga-based interventions can improve overall quality of life and sleep impairments in fibromyalgia patients. Patients reported reduced symptoms of fibromyalgia and there was less catastrophization due to pain post-yoga, as compared to the same course of standard therapy. Pain sensation is significantly altered in fibromyalgia specifically at the localized tender points; yoga practice for four weeks can restore the altered pain sensitivity which leads to enhancement of the musculoskeletal performance and improvement in the cortical plasticity of fibromyalgia patients. Considering the impact of age-old yoga-based interventions on human health and fitness, it is suggested from our study results that yoga is beneficial for fibromyalgia syndrome and associated disabilities. However, till date, only a few studies are available which have objectively assessed the effect of yoga-based therapy in fibromyalgia patients. For robust and firm recommendation of yoga for fibromyalgia patients, objective assessment of pain including biomarker profiling along with real-time neuroimaging of brain, such as Electroencephalography (EEG) and functional Magnetic Resonance Imaging (fMRI) while practicing yoga can be planned. An elaborate electromyographic study of the tender points in addition to these can help in establishing link between peripheral and central control of pain and associated symptoms in fibromyalgia. Group activities are always better for patient compliance and prevention of premature discontinuation of the treatment trials. Therefore, we suggest yoga as a effective , low cost pain management strategy with minimal side effects. Yoga can be implemented on a day-to-day basis for the management of fibromyalgia syndrome.

## Supporting information

none

## Data Availability

All data produced in the present work are contained in the manuscript

