## Supplementary material for "Supervised yogic intervention improves pain, cortical excitability and flexibility in fibromyalgia patients: An objective evidence based of journey from case control study to randomized controlled trial": none

**Supplementary Information**

**Validation of yogic intervention in symptom alleviation in fibromyalgia patients - Methodology**

The study was an open-label, though assessor-blinded randomized controlled trial primarily. More details on the study arms is given in Supplementary Table 1.

**Assessment and study parameters at various time-points**

Total duration of the study was four weeks for Objective 2 and Objective 3, apart from two additional days of visits for the assessments at baseline and post therapy. Waitlisted controls were not asked to stop the ongoing medical treatment except for yogic interventions if advised by their specialists for a total duration of 4 weeks, whereas the yoga group patients were given the option either to stop the ongoing therapy/medications or continue. (Supplementary Table 1)

A total of 47/60 participants in the yoga group preferred to stop the ongoing treatment while performing the planned yoga sessions. Patients referred from the clinics after screening and proper diagnosis, were re-screened according to study recruitment criteria in the Pain Research and TMS Laboratory (Day 0). Patients were given an appointment for medical history, baseline assessment and blood sampling (Day 1). On Day 1, patients had to report to the Pain Research and TMS Laboratory at 09:30 AM empty stomach. An overnight starving blood sample (5 mL) was collected immediately after their arrival in a serum vacutainer tube (yellow). Battery of tests involved in the patients was performed on the same day except for ELISA, for which serum samples were stored in – 20^O^ C immediately after isolation. Supplementary Table 2 lists all the assessment parameters of the pain and associated outcomes performed in the participants.

**Supplementary Table 1. Study groups and their relevant attributes for the recruitment in the study**

| **Characteristics** | **Yoga Group**  **(n = 60)** | **Waitlisted Controls (n = 60)** | **Healthy Controls**  **(n = 60)** |
| --- | --- | --- | --- |
| Sample Size (Ethical Approval) | 60 | 60 | 60 |
| CTRI Requirement | Registered | Registered | Not Applicable |
| Assessment Time-points | Baseline and Post-therapy | Baseline and Post-therapy | Baseline only |
| Study Interventions | Yoga and Standard Therapy | Standard Therapy only | None |
| Number of visits | 22 (1 Baseline, 20 Yoga Sessions & 1 Post-therapy) | 2 (Baseline & Post-therapy) | 1 (Baseline only) |
| Duration of Intervention | 4 weeks | 4 weeks | Not Applicable |
| Participation Consent | Yes | Yes | Yes |

**Supplementary Table 2. Assessments and Study parameters**

| **S. No.** | **Assessment** | **Technique** | **Study Parameters** |
| --- | --- | --- | --- |
| 1. | Pain Status | 1. Visual Analogue Scale (VAS)  2. Pain Catastrophizing Scale (PCS)  3. Short-form McGill Pain Questionnaire (SF-MPQ)  1. Quantitative Sensory Testing (QST) | Descriptor’s score  Pressure Pain Thresholds  Pressure Pain Tolerance |
| 2. | Quality of Life | 1. WHO Quality of Life (WHOQOL) Questionnaire  2. Fibromyalgia Impact Questionnaire (FIQ) | Descriptor’s Scores |
| 3. | Fibromyalgia Associated Symptoms | 1. Pittsburgh Sleep Quality Index (PSQI) | Descriptor’s Score |
| 4. | Musculoskeletal Performance | 1. Sit and Reach test  2. Schober’s Test  3. Goniometry | - Lumbar Flexion  - Lumbar Flexion  - Range of motion |
| 5. | Cortical Excitability | Transcranial Magnetic Stimulation (TMS) | - Resting Motor Threshold  - Motor Evoked Potential  - Cortical Silent Period  - MEP Recruitment Curve  - CSP Recruitment Curve |
| 6. | Quantification of Blood Biomarkers | Enzyme Linked Immono-sorbent Assay (ELISA) | β-Endorphin, Cortisol, Glutamate, Serotonin and Substance P |

Pain assessment (both subjective and objective), recording of corticomotor excitability, measurement of flexibility and range of motion were performed at Pain Research and TMS Laboratory, Department of Physiology, All India Institute of Medical Sciences (AIIMS), New Delhi, at baseline (pre-therapy or at the time of recruitment) and at end of 4-weeks (post-therapy). [1] Subjective parameters for pain such as Visual Analogue Scale, McGill Pain Questionnaire and Pain Catastrophizing Scale; and fibromyalgia specific questionnaires - Fibromyalgia Impact Questionnaire (FIQ) were not performed in the healthy controls. Regular (5 days a week) supervised yogic intervention was carried out at the Integral Health and Wellness Clinic (IHWC), Department of Physiology, All India Institute of Medical Sciences (AIIMS), New Delhi. Quantification of blood biomarkers’ using ELISA for the stored serum samples were performed at the end of the study or whenever respective ELISA kits were available according to the carrying capacity of the kits for the samples. A total of 40 samples were run for ELISA with separate standard curves for each experiment. Biochemical assessments were performed in the Central Biochemical Laboratory, Department of Physiology, AIIMS, New Delhi.

**Descriptors and Questionnaires for Subjective assessment of pain and associated symptoms of fibromyalgia**

Visual analogue scale is a numerical pain rating scale developed as a linear and continuous scale to quantify pain intensity. It is represented as a 11-cm long line with 0= no pain and 10= maximum pain experienced by a subject in the last 24 hours. Fibromyalgia patients were asked to rate their present pain score based on their pain experience in the past week on the scale. The distance between ‘0’ and participant’s score was measured in cm, resulting in scores ranging from 0 to 10. [2] It is validated as an effective tool to measure pain subjectively for fibromyalgia under both clinical and laboratory settings. [3]

McGill Pain Questionnaire (MPQ) is a general questionnaire which is used to describe the attributes of pain in chronic pain patients. It is a score based intensity scale of pain along with the type of pain sensations perceived by the patients. This questionnaire was used to evaluate the quality of patient’s pain perception. It consists of 20 descriptors with four options indicating severity of the symptoms, “0” means absence of the symptom, “1” means mild, “2” means moderate, and “3” means severe. The patient was asked to rate according to the pain experienced in the past one week. The ratings were reported as total scores (sum of rating for all descriptors). The questionnaires were also categorized into four sub-classes: sensory (question 1 to 10), affective (question 11 to 15), and evaluative (question 16), and miscellaneous components of pain (question 17 to 20). [4]

Pain Catastrophizing Scale is a 13-item self-report subjective measure designed to assess the catastrophizing thought of the pain by the patients suffering from chronic pain. It was developed by Sullivan et al. and has been widely used in the research and surveys in chronic pain patients. Pain catastrophization is consistent with the exaggerated nociception in the illness and simultaneous impact from the routine day-to-day lifestyle. This scale has been developed to quantify multidimensional components of catastrophe at descriptive level - rumination, magnification and helplessness as predictor of the intensity of physical and emotional distress. [5] PCS is useful in quantification of pain experience of an individual, with respect to how and what one feels and thinks when apprehended by pain. PCS score is slightly correlated with fatigue, depression and anxiety but it is strongly associated with emotional stress. PCS scores are directly related to the threat associated with the pain condition. Compared to other ways of measuring pain-related thoughts, this questionnaire is unique in that the individual does not need to be in pain while completing it. Engagement of patients with the pain is subjectively self-examined with respect to the disability and threat driven due to illness in fibromyalgia patients. Recently, PCS has been evaluated and validated in fibromyalgia and other chronic pain conditions in different cohorts of patients. [6]

Fibromyalgia Impact Questionnaire (FIQ) is a pain related disability and associate quality of life assessment and evaluation descriptive tool, specifically developed for fibromyalgia patients’ status, motivation, progress and outcomes. It has been designed to measure the components of health status that are believed to be most affected by fibromyalgia. The FIQ is composed of 10 items. The first item contains 11 questions related to physical functioning. Items 2 and 3 ask the patient to mark the number of days they felt well and the number of days they were unable to work (including housework) because of fibromyalgia symptoms. Items 4 to 10 are horizontal linear scales marked in 10 increments on which the patient rates work difficulty, pain, fatigue, morning tiredness, stiffness, anxiety and depression. [7]

Fibromyalgia patients, like other psychiatric disorder patients, often complaint with sleep disturbances, the Pittsburgh Sleep Quality Index (PSQI) was designed to evaluate overall sleep quality in clinical populations. A PSQI score of ≥7 has high diagnostic sensitivity and specificity in distinguishing patients with sleep problems from normal subjects. [8] Each item among the 19-items of the descriptor belongs to one of the seven subcategories: subjective sleep quality, sleep latency, sleep duration, habitual sleep efficiency, sleep disturbances, use of sleeping medication, and daytime dysfunction. Five additional questions rated by the respondent’s roommate or bed partner are included for clinical purposes which are not scored. This questionnaire consists of a combination of Likert-type and open-ended questions. Respondents are asked to indicate how frequently they have experienced certain sleep difficulties over the past month and to rate their overall sleep quality. Scores for each question range from 0 to 3, with higher scores indicating more acute sleep disturbances. The test has recently been successfully translated and validated for Indian population.

World Health Organization defines quality of life as - “An individuals’ reception of his position in life in context to culture and value systems in which they live and in relation to their goals, expectations, standards, and concerns”. This questionnaire has 26 items to assess the overall quality of life affected by chronic pain in the past two weeks. [9] It is a valid assessment tool consisting of four domains: physical, psychological, social, and environmental domains and is sound and cross-culturally accepted. Each item has five options, higher score indicating increasing quality of life, except Q3, 4, and Q26 which are negatively scaled. The vernacular version questionnaire was validated and is applicable for Hindi-speaking patients. [10]

**Quantitative Sensory Testing (QST): Objective pain assessment**

Algometer is a device which is designed to quantify and document levels of tenderness and pain sensitivity by measuring pressure pain threshold and pressure pain tolerance. Pressure algometry is a reliable measure of pain in muscles, fascia, joints, tendons, ligaments and the periosteum. Medoc AlgoMed; (Model: Wagner FPIX Instrumentations, Medoc, Ramat-Yishai, Israel) is a computerized (digital) pressure algometer device for the quantitative assessment of small nerve fibre dysfunctions by measuring the sensory thresholds for tenderness in term of pain sensitivity in chronic pain patients. It has three components apart from a system which has the pre-installed software tool to establish communication and run the device. i) A digital Algometer (Medoc AlgoMed) is the primary unit which is attached with the system through a USB port. Power consumption of the AlgoMed Device is DC 5 Volts and 0.5 A; ii) A sensory output feedback device called Patient Response Unit or just the response unit which is connected along with the algometer in an another USB port of the system to communicate through Medoc Main Station software tool, and iii) A CoVAS (**Co**mputerized **V**isual **A**nalogue **S**cale) device which is also attached through USB to scale the ratings of pain experienced by the patient. Pressure pain usually deviates from the normal range in peripheral nerve disease, neuropathies and chronic pain conditions. Supplementary Figure 1 shows components and details of Medoc AlgoMed device.

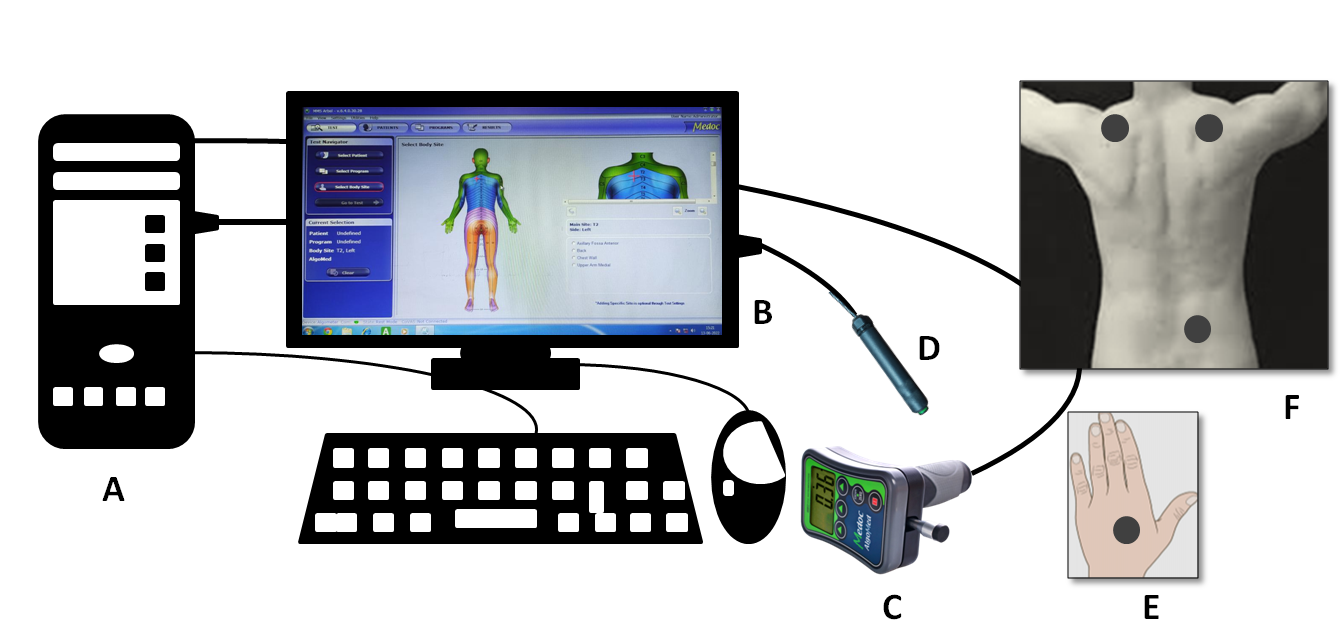

**Supplementary Figure 1:** **Schematic representation for the Set-up of Quantitative Sensory Testing. A. Central Processing and Data Storage System; B. Monitor for Recording Pain Sensitivity; C. Medoc AlgoMed Device; D. Patient Response Unit; E. Reference Site for QST (Dorsum of hand and F. Test sites for QST. Each circular solid dot represents the site for Algometer placement at reference and test sites.**

***Principle of operation***

Quantitative sensory testing equipment for the pressure pain parameters are widely used to assess tenderness at the test site which was basically tender points in the fibromyalgia patients. Pressure being a mechanical force can be used to measure the hyperalgesia and allodynia related to the pain perception. Responses of the individual were recorded in term of kiloPascals (kPa). The minimum intensity of pressure which generates the very first pain (VAS Score of 1) is called the pressure pain threshold whereas the maximum intensity of pain that a patient can bear (VAS score of 5) is called the pressure pain tolerance. These two parameters are very important in assessing the pain status of the patients before and after an intervention. For quantifying pressure pain parameters, a small circular rubber knob with flat surface is used which is capable of delivering a uniform and blunt pressure on the localized target dermatome on the patient's skin. Algometer works on the principle of load cells which generates a quick balanced force manually by the operator by wrapping the ergonomic hand grip. Pressure exerted on the test site was in close proximity with the algometer tip having a circular interface of 1 cm^2^ surface area. Pressure was implied manually by holding the AlgoMed device (main unit) at a right angle to the test surface. Algometer has an ergonomic handle that allows the operator to grip it and apply embedded pressure. Pressure algometry using manual mode helps applying a continuous ascending pressure at a constant rate on the patient's tender points, with its real time monitoring on the screen of the computer. Patients can stop the test at any time by clicking the Patient Response Unit. Alternatively, the operator can stop the test by simply removing the AlgoMed device from the patient's selected target site. The AlgoMed software (Medoc Main Station) offers body site selection, and any additional body site can be personalized in the software. The software allows easy management of patients, programs and test results. Results were displayed in a customizable colour report which is stored in the system and can be exported for further analysis, after the test or later. Pressure pain parameters can be assessed using mechanosensory force exerted per unit area on the dermatomes of the test site by holding the ergonomic handle of the AlgoMed device and exerting pressure at a constant rate. Mechanical stimuli per se pressure modality can be regulated by examiner and the subject both; former can stop the test by removing the algometer from the test site whereas the subject can respond using the ‘response unit’ held in the hand. The pressure is increased slowly and uniformly at the test site till the pain threshold or the pain tolerance threshold at the test site is elicited in fibromyalgia patients or healthy subjects. [11] Supplementary Figure 1 demonstrates the actual set-up for the recording.

***Test Instructions:*** Quantitative Sensory Testing (QST) was done at 09:30 AM at time the patient reported early morning. Water was allowed. Participants were told to sit on the comfortable chair with their eyes closed in a temperature regulated room and a trial run was carried out to remove any anticipatory false responses. They were explained about the test and numerical pain rating scale (VAS scoring) - a continuous 11-point visual analogue scale for rating the pain intensity. They were then explained the use of ‘patient response unit’ and press vertically to stop the ongoing pressure exertion.

***Test Protocol -***

- Participants were instructed to sit on the comfortable chair and the test paradigm was explained along with introducing the pain rating scale. Patient was asked to be attentive and focus on the test, and not to look at the screen but can close their eyes without sleeping.
- After switching on the computer, the AlgoMed device and Patient Response Unit was plugged into the USB port of the system.
- Test sites were wiped out with spirit using a sterile cotton swab.
- ‘Medoc Main Station’ software was opened and the participant’s profile was created which includes full name, details of birth, unique test identity (enrolment number), gender and assessor’s name.
- The AlgoMed device was now switched on and calibrated by pressing the ‘Escape Zero’ button and holding it in the vertical position, right-angled to the dermatomal surface. Autozero will reset the device by nullifying force per unit area which can render false positive results due to machine error.
- Entered the patient profile and selected dermatomes for first the reference site (dorsum of right hand) and then test sites (left shoulder, right shoulder and lower back) for performing sensory testing using pressure modality (Supplementary Figure 1). ‘Go To Test’ menu was selected and the test was started by the operator by holding the AlgoMed device in the right hand using an ergonomic handle. Participants were instructed to hold the response unit in their right hand and press it quickly to respond, when VAS =1 (Pressure Pain Threshold) or VAS = 5 (Pressure Pain Tolerance) was felt over the test site by the exerted pressure using the device.
- Pressure was increased very slowly and uniformly at the constant rate (rate not more than 50 kPa/second) between dotted line (window) to avoid large standard deviation in the test.
- Four trials were performed at all sites which were averaged at the end of the test which can be noted down from the test report.
- Data was imported to an external file and analyzed offline. The results of all the participants of the same group were averaged for each test site separately at the end of the study.

**Flexibility and range of motion assessment**

Lumbar flexibility was measured using Schober’s test and Sit and Reach Box while Range of motion was measured using a Goniometer. Schober’s test was performed using a simple measuring tape.

***Goniometer***

Goniometry is the science of measuring joints’ range of motion in the plane of the joint and a goniometer is an instrument that measures the maximum possible [range of motion](https://www.physio-pedia.com/Range_of_Motion) along a joint in clinical settings to assist in surgical recovery, treatment and related rehabilitation. It was earlier used as the standard tool for measuring the degree of dorsiflexion at Ms Johnson’s ankle and an absent reflex is determined by placing it in a neutral position before striking the stretched Achilles tendon with the percussion hammer (Nursing Clinics of North America, 2014). There are two "arms"— one “stationary” and one “mobile” and both are hinged together at the centre of the circular pie scale (Supplementary Figure 2). They can be positioned at specific points on the body with the centre of the fixed goniometer arm aligned at the joint of interest. Marks over the hinge allow precise measurement of range of motion in degrees. We have used a standard goniometer (SAEHAN^®^, South Korea) for measuring lumbar flexion before and after yoga therapy; whereas healthy controls were assessed once at baseline only. [12]

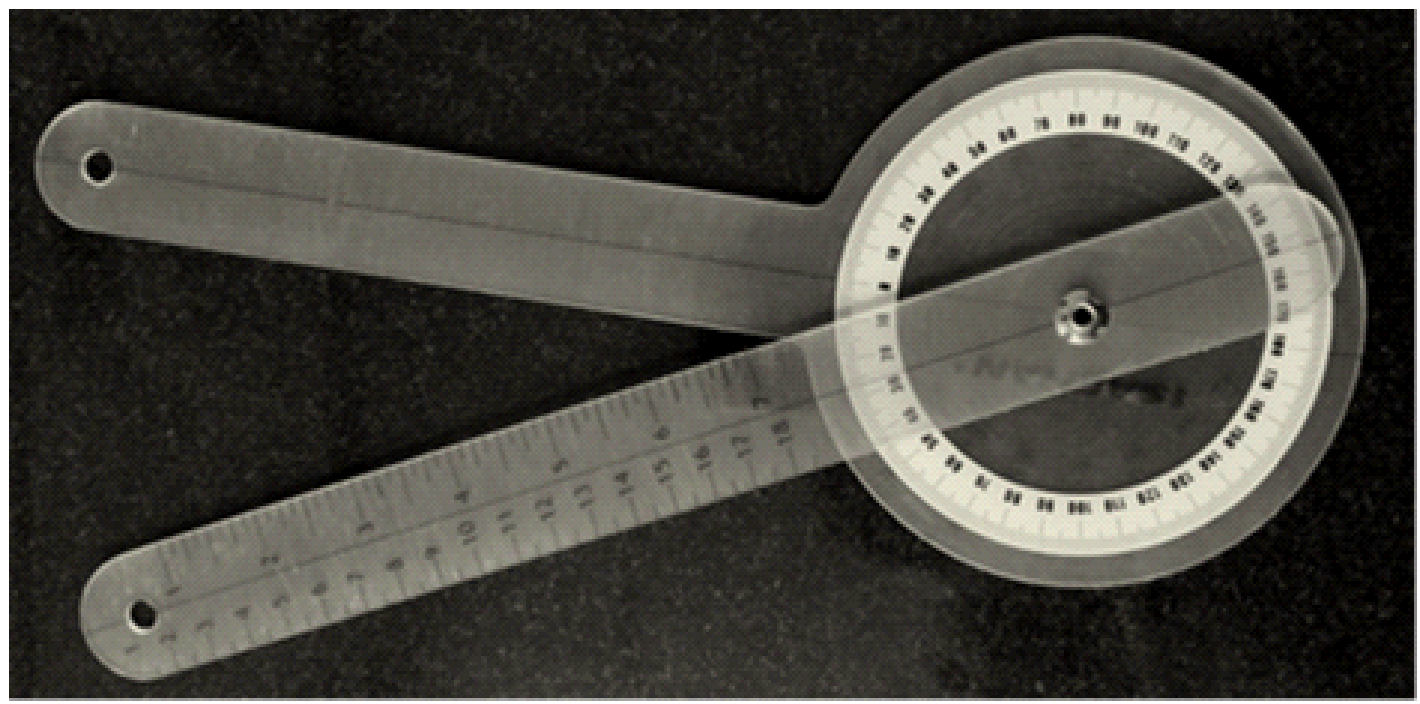

**Supplementary Figure 2: Goniometer used for measurement of range of motion and lumbar flexion of the study participants.**

Test Protocol -

- Ask the participant to stand aside with its back facing straight with the hands touching side of the respective leg.
- Now, mark the L3-L4 region with a dot.
- Place the axis of the goniometer (hinged pie scale) fixed at the dot at L3-L4 vertebrae.
- Align the inactivated goniometer along the participants’ vertebral/spinal axis.
- Now, ask the participant to bend first toward left and then toward right slowly.
- Rotate and align the free/mobile arm of the goniometer along with the extent of the participants’ flexion in lateral direction slowly.
- The angle measurement was done thrice and averaged for each side (left and right).

***Modified Schober's test***

The Modified Schober’s test is the simplest and renowned method for measuring lumbar flexibility and associated range of motion using a measuring tape.

Test Protocol -

- Modified Schober’s Test uses two marks - one over the spine connecting two posterior superior iliac spines (PSIS) and another over 15 cm superior to the first mark. It eliminates the errors in identification of lumbosacral junction and makes sure that the entire lumbar spine was included.
- Measurements of lumbar flexion and extension were carried out with measuring tape using standardized method of Modified- Schober’s Test based on guidelines provided in the American Medical Association (AMA) Guides (1993).
- The participants were instructed to remove their shoes and expose their back from gluteal fold to mid-thoracic spine with left and right PSIS fully exposed.
- Then participants were asked to stand erect, arms at their sides, and feet placed apart parallel to shoulders.
- For better clarity to the participants, the examiner demonstrated the proper procedure of forward bending with the arm hanging in front and keeping knees straight.
- Examiner kneeled behind the standing participants and identified both the PSIS with her thumb. Inferior margins of the volunteer’s PSIS were marked, and a ruler was used to locate and mark a midline point on sacrum (inferior mark).
- Then the final mark (superior mark) was marked on the lumbar spine 15 cm above the midline sacral mark (inferior mark).
- The examiner then aligned the tape measure between two skin marks with zero at inferior mark and 15 cm at superior skin mark.
- The measuring tape was kept firmly against the participants’ skin while they were asked to bend forward with the instruction “Bend forward as much as you can while keeping your knees straight”. The measuring tape was maintained against the participant’s back during the movement but was allowed to unwind to accommodate motion.
- For each of the spinal motion measured, the end of the range of motion was defined by instructing the participants to report that they cannot move any further.
- At the end of flexion, the distance between the two marks was noted. The range of motion was the difference between 15 cm and the mark made anterior to it (length measured at the end of motion).
- After each measurement, the participant was told to come back to a comfortable/normal standing position.

***Sit and Reach box (American College of Sports Medicine)***

The Sit and reach test is one of the linear [flexibility](https://www.physio-pedia.com/Flexibility) tests used for measurement of extensibility of the hamstrings and lower back. It is a compact useful tool with dimensions of 21" x 12" x 13". The Standard testing box has a chart over the top of the box that helps to evaluate testing results, based on which the flexibility can be classified as either poor, satisfactory, fair, good and extremely good. It has a smooth gliding stretch indicator called slider, which holds at the maximum reading until reset, ensuring accurate results. Slider moves forward and backward between two scaled linear spaces. [13] The box is easy to store and transport as is lightweight and compact (Supplementary Figure 3).

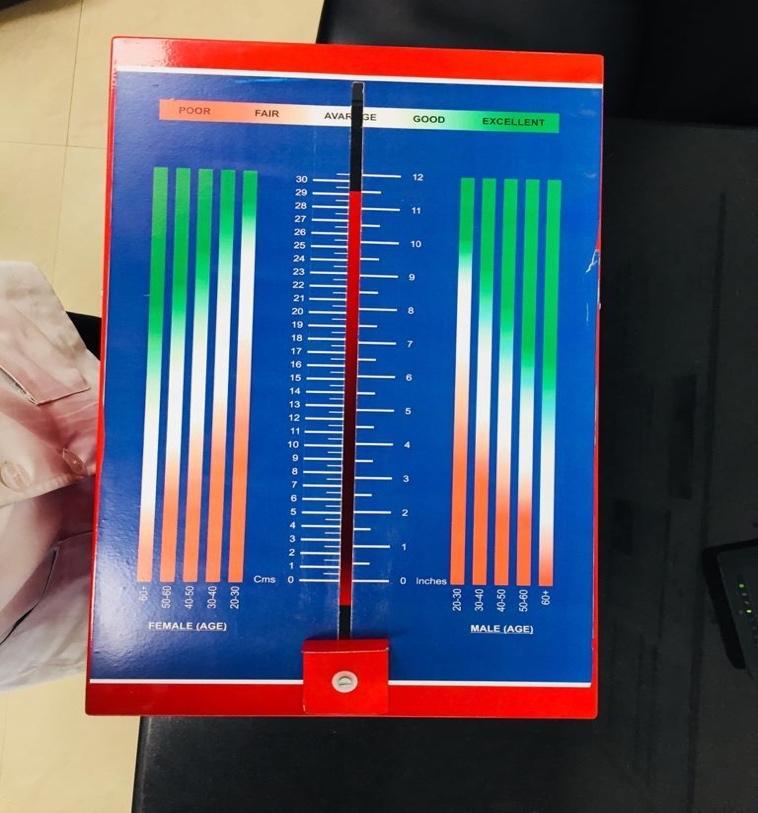

**Supplementary Figure 3: Sit and Reach test box, used for assessment of flexibility of lower back.**

Test Protocol -

- Participants were asked to sit comfortably on the yoga mat with his head facing toward the sit and reach box and asked to lift both of their hands parallel in the air and take down slowly to touch the slider of the Sit and Reach box.
- With the palms facing downwards the subject was asked to reach forward along the measuring line as far as possible and the distance was recorded. They were also instructed to perform the test in one smooth movement rather than a bouncing or jerking movement.
- The participant was instructed by the examiner to slowly reach forwards as far as possible with the palms of their hands parallel and hold that stretch for three seconds. It was also ensured that the participants always kept their hands on the box with their knees extended and normally breathing throughout the test and did not hold their breath.
- The test was repeated thrice, and values were noted in centimetres and averaged.
- Note that The American College of Sports Medicine (ACSM) presented normative values for adults completing the Sit and Reach Test and the values obtained of the participants were compared to them, indicating grading of flexibility as poor, fair, and good.

**Transcranial Magnetic Stimulation (TMS): Assessment of cortical excitability**

Transcranial Magnetic Stimulation (TMS) is a non-invasive brain stimulation technique to assess cortical excitability and also to provide therapeutic interventions to patients suffering from depressive disorders and chronic pain conditions. For depression it has been FDA (Food and Drug Administration), USA approved in 2008. Neurosoft TMS (Model- Neuro MS/D) device with self-cooled figure-of-eight coil FEC-02-100-C was used for recording corticomotor excitability in fibromyalgia patients and age and gender matched healthy controls. The coil has two loops of wiring which are wound in a figure of 8 shapes so that coils lies side by side in centre. It generates a more focused and localized magnetic field at the intersection point of two loops inside and gives stimulation at specific areas with greater precision than the circular and butterfly coils. Two coils are slightly bent below the horizontal plane to adjust to the contour of the skull and proper placement over the scalp. It can penetrate up to 5 cm inside the scalp. Maximum output capacity of the magnetic stimulator unit of the TMS device is 4 Tesla. Standard biphasic pulse has duration of around 250-330μs. TMS Compatible Electrode: Motor Evoked Potential (MEP) recording electrodes are A-1000 Ag-AgCl ECG patch electrodes (Fiab, Italy) were placed over belly of Abductor Pollicis Brevis (APB) muscle of left hand and similar reference electrode placed over the interphalangeal joint of right thumb. Electrodes (Anode, Cathode and Neutral) were separately connected to the MEP acquisition box by alligator clips. Earthening (Grounding) was provided using a button electrode placed inside a velcro strap wrapped around the wrist over the styloid process of ulna. Figure of eight coil (F8 coil) was placed over the scalp which was then navigated to be set at a point where the TMS pulse produced the highest amplitude of MEP. Maximum Stimulator Output (MSO) is the maximum capacity of the machine to deliver a stimulus at the target site; thresholds for the participants were determined according to the percentage of the maximum output of the stimulator. [14]

Transcranial Magnetic Stimulation represents a painless and non-invasive technique to investigate the function and integrity of the primary motor cortex (M1) and corticospinal pathway. It is based on Faraday’s law of induction and Lenz’s law. Faraday’s law of induction makes use of a magnetic flux through a region of space inside the brain. Parameters recorded were Resting Motor Threshold (RMT), Motor Evoked Potential (MEP), Cortical Silent Period (CSP), MEP AND CSP Recruitment Curves.

**ΦB = ∫B·dA**

Σ where, ΦB = magnetic flux through a region of space; dA = element of surface Σ enclosed by a loop of wire, B = magnetic field enclosed by a wire loop.

A time-varying magnetic field created by an electrically-generated magnetic object (like the Transcranial magnetic stimulation coil) can induced the generation of **Eddy’s currents** or Foucalt’s currents in the nearby conductive, stationary object vicinity (like our brain tissues). The magnitude of the current generated would be proportional to the strength of the magnetic flux, area of the loop, rate of change of the flux, and resistivity of the tissue. Barker et al. in 1985 reported induction of a magnetic ﬁeld over M1 by a coil (where a transient and large electrical current transits from a capacitor system) could depolarize the corticospinal cells. At a sufficient level of intensity, the stimulus produces a muscle response; referred to as motor evoked potential (MEP) recorded by electromyography (EMG) electrodes. MEP latency and amplitude are considered the primary outcomes studied to probe the corticospinal function. [15]

Neurosoft MS/D machine comprises of three units to deliver non-invasive stimulus – a) Main unit (or magnetic stimulator) which controls all other units and can be used as a stand-alone machine (if equipped with the coils). The front panel contains digital indicators displaying all parameters of stimulator and control output to adjust parameters. To attach the stimulator to PC, main unit has USB port. Stimulus delivery can be regulated from both the display board over here and software tool as well. b) Cooling unit which is a significant part of therapeutic magnetic stimulator. There is a tank with coolant filled inside it, high-tech compressor-free cooler and the pump running continuously the coolant via the coil to ensure the baseline temperature so that machine can perform under the safety limits of the temperature range. The liquid cooling system is much more efficient and less noisy in comparison with the air ones. And, c) Extra power supply unit is designed to operate with up to 30 Hz frequency. However, the main unit alone can deliver maximum intensity only at 5-7Hz frequency. The extra power supply unit makes it possible to increase the maximum frequency up to 100 Hz and obtain the maximum intensity at 20-25 Hz frequency depending on coil type (Supplementary Figure 4).

***Test Protocol (for recording Cortical Excitability parameters)***

Methods for assessing corticomotor excitability using TMS are described in accordance with the latest guidelines on its methodology for reporting. [16] Participants were asked to sit on a comfortable chair with elbow at rest and hand flexed facing straight without avoiding strain over the neck. Surface electromyographic activity was recorded from left abductor pollicis brevis muscle using disposable Ag/AgCl electrodes (7mm; Fiab, Vicchio Firenze, Italy). Pulses were delivered with a figure-of-eight transcranial magnetic stimulation coil (70 mm; Neuro-MS/D, Ivanova, Russia) at an angle of 45º to the sagittal plane of the scalp to generate antero-posterior waves inside the brain. Inter-stimulus intervals were randomized between 5 to 10 seconds depending on the previous stimulus. Figure –of-eight coil can penetrate a maximum of 5 cm inside the scalp. The acquired signals were amplified (1000x), bandpass filtered between 10 Hz to 2 kHz (Neuro-MEP-Micro, Ivanovo, Russia) and analysed offline (Neurosoft.EP software, Ivanovo, Russia).

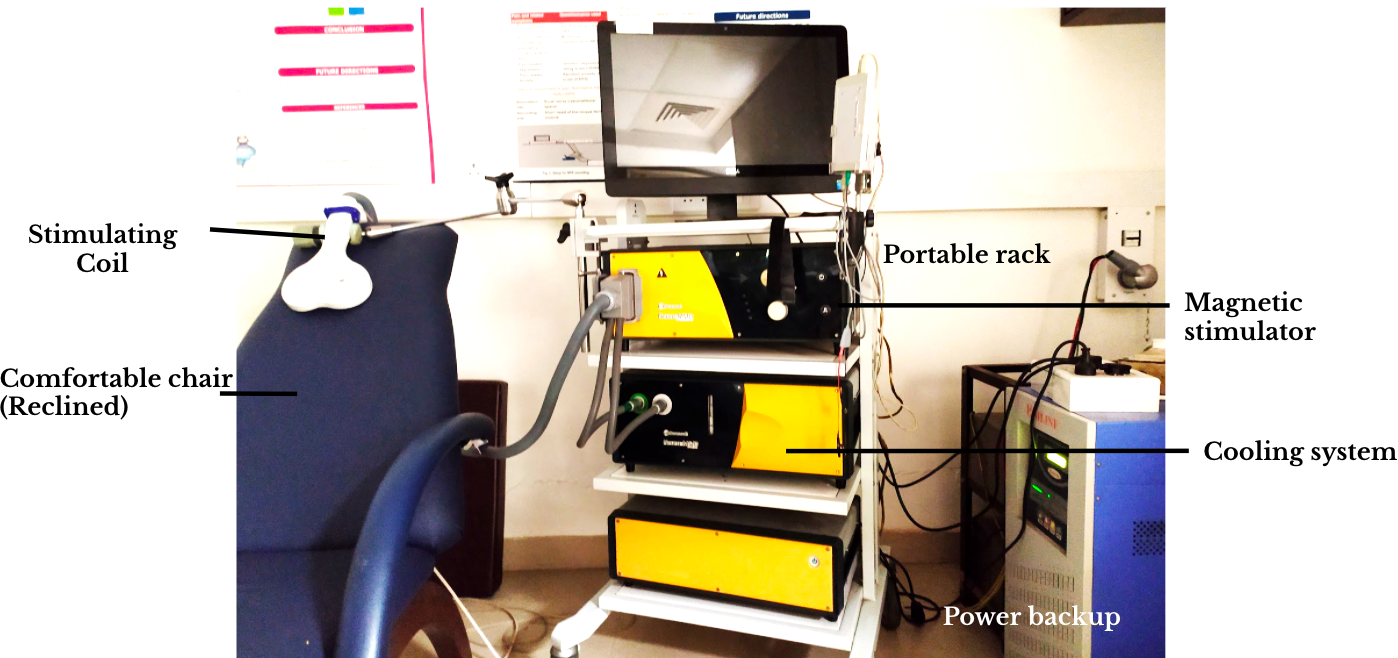

**Supplementary Figure 4: Set-up of Transcranial Magnetic Stimulation (TMS) machine at Pain Research and TMS Laboratory (Figure of 8 Stimulating Coil, Portable rack, Magnetic stimulator, Comfortable chair (Reclined), Cooling system), at pain research and TMS laboratory, Department of Physiology, AIIMS, New Delhi.**

**Determination of tentative motor hotspot**

Scalp landmarks - vertex, nasion and inion was marked; on the basis of inter-tragus and nasion-inion arbitrary lines using EEG 10:20 system, a ‘tentative hotspot’ for the hand region of the primary motor cortex was localized. Localization of tentative hotspot is marked as a light dot as it can be or cannot be the actual target site for stimulation. (Supplementary Figure 5) Therefore, confirmation of the motor hotspot was done by placing TMS coil on the scalp on the sagittal plane and delivering few pulses at the tentative hotspot in steps of increasing the percentage of maximum stimulatory output (MSO %) till a visible twitch was observed in the thumb. Failing to achieve the same, we have started navigating in both the sagittal and coronal planes to find out the actual hotspot for M1. Following cortical excitability parameters were recorded using TMS both before and after the yogic intervention (Supplementary Figure 4).

Electrodes (Anode, Cathode and Neutral) were separately connected to the MEP acquisition box by alligator clips. Earthening (Grounding) was provided using a button electrode placed inside a velcro strap wrapped around the wrist over the styloid process of ulna. Figure-of-eight coil (F8 coil) was placed over the scalp which was then navigated to be set at a point where the TMS pulse produced the highest amplitude of MEP. Maximum Stimulator Output (MSO) is the maximum capacity of the machine to deliver a stimulus at the target site, i.e. 100%; thresholds for the participants were determined according to the percentage of the maximum output of the stimulator. Supplementary Table 3 summarises stimulation paradigm of MEP-Recruitment Curves of all the study groups.

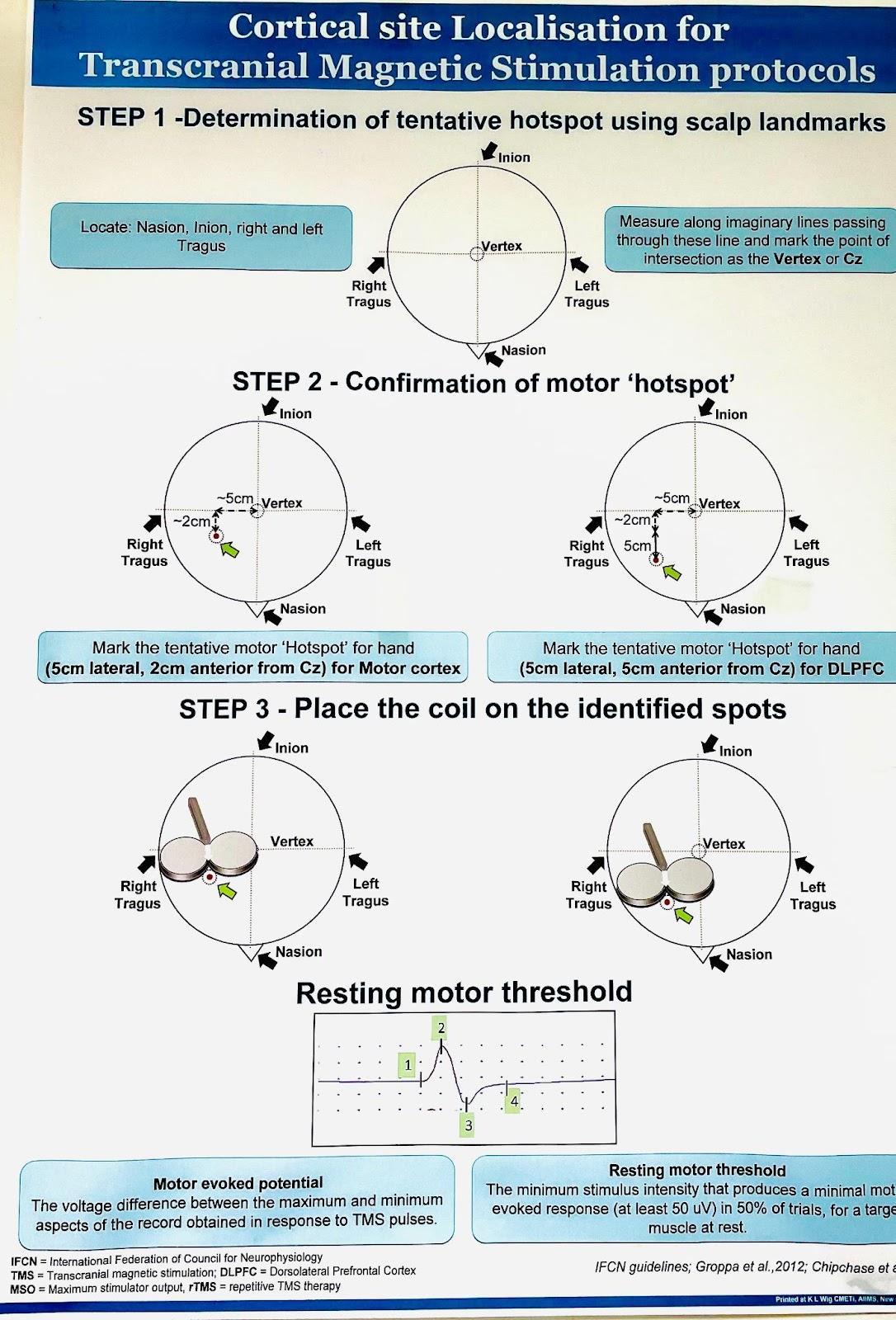

**Supplementary Figure 5: Figure represents manual method for determination of tentative motor hot spot.**

**Supplementary Table 3. Stimulation paradigm of MEP-Recruitment Curves of all the groups**

| **Healthy Controls** | | | |
| --- | --- | --- | --- |
| **Percentage Maximum Stimulator Output (MSO)** | **Latency (ms)** | **Duration (ms)** | **Area (Amplitude x Duration)** |
| **90 %** | 20.18±2.76 | 8.48 ±3.75 | 0.09 ±0.07 |
| **100 %** | 20.73 ± 7.13 | 9.62 ±2.92 | 0.17 ±0.95 |
| **110 %** | 19.70 ±6.93 | 12.29 ± 4.51 | 0.64 ± 0.50 |
| **120 %** | 21.50 ± 4.67 | 12.83 ± 3.34 | 1.24 ± 0.73 |
| **130 %** | 20.82 ± 2.69 | 14.32 ± 3.26 | 2.58 ± 1.77 |
| **140 %** | 21.55 ± 5.83 | 14.85 ± 3.64 | 3.73 ± 1.89 |
| **150 %** | 21.24 ± 0.87 | 15.46 ± 2.74 | 4.51 ± 1.99 |
| **Fibromyalgia Patients of Yoga Group** | | | |
| **Percentage Maximum Stimulator Output (MSO)** | **Latency (ms)** | **Duration (ms)** | **Area (Amplitude x Duration)** |
| **90 %** | 22.16 ±7.95 | 8.39 ±4.13 | 0.17 ±0.08 |
| **100 %** | 25.56 ± 3.21 | 9.84 ±2.93 | 0.26 ±0.25 |
| **110 %** | 21.80 ±3.24 | 13.15 ± 2.81 | 1.01 ± 2.06 |
| **120 %** | 21.27 ± 1.89 | 13.70 ± 2.98 | 1.27 ± 1.14 |
| **130 %** | 20.89 ± 4.29 | 14.30 ± 3.64 | 2.91 ± 3.69 |
| **140 %** | 20.95 ± 1.50 | 14.85 ± 3.28 | 3.72 ± 4.15 |
| **150 %** | 21.06 ± 1.49 | 18.83 ± 2.79 | 5.93 ± 5.12 |
| **Fibromyalgia Patients of Waitlisted Controls** | | | |
| **Percentage Maximum Stimulator Output (MSO)** | **Latency (ms)** | **Duration (ms)** | **Area (Amplitude x Duration)** |
| **90 %** | 23.46 ±7.73 | 12.12 ±11.70 | 1.17 ±3.25 |
| **100 %** | 22.11 ± 6.38 | 10.73 ±3.77 | 0.39 ±0.81 |
| **110 %** | 27.91 ±3.04 | 12.84 ± 2.90 | 0.85 ± 0.82 |
| **120 %** | 21.18 ± 2.75 | 13.62 ± 3.14 | 1.74 ± 1.38 |
| **130 %** | 20.92 ± 3.37 | 14.53 ± 2.83 | 2.74 ± 2.27 |
| **140 %** | 20.51 ± 2.06 | 13.65 ± 2.83 | 3.78 ± 2.88 |
| **150 %** | 20.63 ± 1.48 | 13.78 ± 2.44 | 6.62 ± 5.71 |

Recruitment curves for MEP and CSP for the respective data sets are depicted in Supplementary Table 4 and 5 respectively for the fibromyalgia patients at baseline and four weeks of regular yogic intervention and standard care therapy.

**Supplementary Table 4: Motor Evoked Potential Recruitment Curves (MEP-RC) of Fibromyalgia Patients of Yoga Group and Waitlisted Controls at Baseline and Post-therapy.**

| **% RMT/mV** | | **Yoga Group (n=60)** | **Waitlisted Controls (n=60)** | **p-value (between groups)** |
| --- | --- | --- | --- | --- |
| 90 % | Pre-therapy:  Post-therapy:  p-value: (within group) | 0.037 ± 0.003  0.036 ± 0.001  0.827 | 0.041 ± 0.004  0.031 ± 0.002  0.089 | 0.462  **0.023** |
| 100 % | Pre-therapy:  Post-therapy:  p-value: (within group) | 0.102 ± 0.013  0.055 ± 0.004  0.082 | 0.106 ± 0.014  0.087 ± 0.004  0.667 | 0.476  **0.027** |
| 110 % | Pre-therapy:  Post-therapy:  p-value: (within group) | 0.350 ± 0.038  0.369 ± 0.036  **0.021** | 0.324 ± 0.041  0.324 ± 0.033  0.875 | 0.625  **0.024** |
| 120 % | Pre-therapy:  Post-therapy:  p-value: (within group) | 0.731 ± 0.068  0.653 ± 0.060  **0.020** | 0.581 ± 0.065  0.605 ± 0.063  0.451 | 0.117  **0.029** |
| 130 % | Pre-therapy:  Post-therapy:  p-value: (within group) | 1.12 ± 0.11  0.972 ± 0.15  0.280 | 1.01 ± 0.11  0.977 ± 0.11  0.776 | 0.509  0.976 |
| 140 % | Pre-therapy:  Post-therapy:  p-value: (within group) | 1.52 ± 0.15  1.46 ± 0.19  0.580 | 1.42 ± 0.16  1.32 ± 0.14  0.327 | 0.625  0.554 |
| 150 % | Pre-therapy:  Post-therapy:  p-value: (within group) | 2.51 ± 0.32  2.10 ± 0.31  0.332 | 2.10 ± 0.23  2.06 ± 0.25  0.998 | 0.308  0.723 |

*Motor Evoked Potential (MEP) Recruitment Curves of fibromyalgia patients of yoga group and waitlisted controls. Data was normally distributed (Gaussian distribution) – ‘Shapiro-Wilk test’; expressed in Mean ± SEM. Paired t-test was performed for intragroup comparison between pre-yoga and post-yoga; two sample (independent) t-test was performed for intergroup comparison between pre- and post-therapy changes. p-value < 0.05 was considered significant. MEP – Motor Evoked Potential (measured in mV); RMT – Resting Motor Threshold.*

**Supplementary Table 5: Cortical Silent Period Recruitment Curves (CSP-RC) of Fibromyalgia Patients of Yoga Group and Waitlisted Controls at Baseline and Post-therapy.**

| **% RMT/ms** | | **Yoga Group (n=60)** | **Waitlisted Controls (n=60)** | **p-value (between groups)** |
| --- | --- | --- | --- | --- |
| 90 % | Pre-therapy:  Post-therapy:  p-value: (within group) | 88.81 ± 4.44  92.43 ± 4.95  0.701 | 102.05 ± 4.97  94.34 ± 4.82  0.244 | 0.051  0.785 |
| 100 % | Pre-therapy:  Post-therapy:  p-value: (within group) | 116.70 ± 6.32  109.60 ± 5.49  0.246 | 112.25 ± 5.69  119.59 ± 6.01  0.293 | 0.602  0.229 |
| 110 % | Pre-therapy:  Post-therapy:  p-value: (within group) | 133.99 ± 7.41  135.18 ± 7.01  0.828 | 146.06 ± 5.81  139.82 ± 8.03  0.496 | 0.198  0.670 |
| 120 % | Pre-therapy:  Post-therapy:  p-value: (within group) | 156.17 ± 6.82  159.84 ± 6.31  0.513 | 161.74 ± 5.85  169.51 ± 6.81  0.394 | 0.534  0.305 |
| 130 % | Pre-therapy:  Post-therapy:  p-value: (within group) | 174.09 ± 7.89  160.12 ± 6.87  0.474 | 178.92 ± 6.57  177.41 ± 7.80  0.878 | 0.637  0.103 |
| 140 % | Pre-therapy:  Post-therapy:  p-value: (within group) | 184.21 ± 7.14  191.19 ± 11.24  0.383 | 194.89 ± 6.66  181.22 ± 7.48  0.211 | 0.276  0.452 |
| 150 % | Pre-therapy:  Post-therapy:  p-value: (within group) | 192.04 ± 9.31  197.25 ± 9.47  0.911 | 192.50 ± 10.54  193.86 9.69  0.766 | 0.974  0.803 |

*Data was normally distributed (Gaussian distribution) – ‘Shapiro-Wilk test’; expressed in Mean ± SEM. Paired t-test was performed for intragroup comparison between pre-yoga and post-yoga; two sample (independent) t-test was performed for intergroup comparison between pre- and post-therapy changes. p-value < 0.05 was considered significant. RMT – Resting Motor Threshold; CSP – Cortical Silent Period (measured in ms).*

**Enzyme Linked Immunosorbent Assay (ELISA): Objective measurement of pain and associated symptoms of fibromyalgia**

Ready-made Enzyme linked Immuno-Sorbent Assay (ELISA) kits which were pre-standardised in our laboratory in several other experiments, were used to quantify the blood biomarkers. Fresh blood sample (5mL) was collected in a sterile vacutainer (Equi Vac. Gel & Clot Activator, Krupa Labequi, India) by venipuncture with single use 24 guage needle. Used needles were disposed immediately using a needle incinerator (Sapna Health Solutions, India). Serum was isolated directly by centrifuging 13 X 75 mm blood collection tube at 3000 rpm in a centrifugation machine with capacity of 6 tubes (R-303, Remi Elekrotechnik Ltd. Vasai, India) in the Pain Research and TMS Laboratory. Serum was stored at -20° C for its further use during the ELISA experiment using ready de kits (Manufacturer – FineTest Pvt. Ltd. Wuhan, China). In vitro quantitative determination of biomarkers’ concentrations in 50 μL serum samples of participants were performed in the Central Biochemical Facility, Department of Physiology, AIIMS, New Delhi at room temperature. Mixing of reagents and incubation of ELISA plates were carried out using Hermle M130 Vortex Mixer and Picofuge Mixer and GeNei^TM^ 37°C incubator (Without CO_2_ Feeding). The biomarkers’ assay was performed by the Competitive-ELISA detection method. The microtiter plate provided in the kit has been pre-coated with neurotransmitter. During the reaction, neurotransmitter in the samples or standard solutions competes with a fixed amount of Β-Endorphin on the solid phase supporter for sites on the Biotinylated Detection Antibody specific to it. Excess conjugate and unbound sample or standard are washed from the plate, and HRP-Streptavidin (SABC) is added to each microplate well and incubated. Then, 3,3′,5,5′ Tetramethylbenzidine (TMB) substrate solution was added to each well. The enzyme-substrate reaction is terminated by the addition of an acid solution and the colour change was measured spectrophotometrically at a wavelength of 450nm (Spectrophotometer Model: Epoch 2.0, BioTek, Agilent Technologies, Santa Clara, USA). Concentrations of neurotransmitter in the samples were then determined by comparison of the ODs of the samples to the standard curve. The concentration of the target substance was inversely proportional to the OD450 value recorded in the 96-well ELISA plates.

***Blood Sampling***

For blood sampling, collected samples (whole blood sample) were kept at room temperature for 2 hours and centrifuged for 15 minutes at 1000 xg or 3000 rpm. Supernatant was collected in a separate vial and stored immediately at -80^o^ C. We have made 5 separate aliquots for each blood sample for the later assays.

***Precautions for the ELISA experiment***

- Blood collection tubes were disposable and free from any endotoxin.
- Hemolyzed and lipemic samples were avoided.
- The best sample storage condition: less than 5 days at 2-8℃; within 3 months at -20℃; within 2 years at -80℃. Samples were stored in liquid nitrogen for a longer storage.
- When using different Elisa kits, proper labelling was done to avoid mixing of components and failed assay.
- Sterile and disposable tips were utilised during the assay.
- After use, the reagents bottle cap were tightened to avoid the microbial contamination and evaporation.
- While washing, tips or pipettes for adding wash buffer were kept away from the well; because insufficient washing or contamination can easily causes false positive and high background.
- During the assay, required reagents for next step were prepared freshly in advance. After washing, reagents were added into the well in time to avoid dryness; as dry plate can result in the failure of the assay.
- Before confirmation, reagents from other batches or sources were not used in this kit.
- Tips and tubes to were not reused to avoid cross contamination.
- After loading, plates were sealed to avoid the evaporation of the sample during incubation.
- We have completed the incubation process at recommended temperature.
- Lab coat, mask and gloves were always put on while doing experiment. Especially, while loading blood samples of the patients or other body fluid samples; followed the regulations on safety protection of biological laboratory.
- Zero tube was stored with dissolved standards at 2-8℃ and use it within 12 hours. Other diluted working solutions containing standards were used within 2 hours.
- Primary and secondary antibodies were used within 30 minutes of its making and can't be stored for a long time.
- For each test a separate standard curve was plotted.
- Reagent reservoirs were strictly not used by HRP couplings or antibody in the subsequent next step directly without proper rinsing.

***Standard Preparation***

Standard tube provided in the kits was centrifuged for 1 minute at 10000g. It was labelled as ‘Zero Tube’. We added 1 mL of sample dilution buffer into the standard tube and tightened the tube cap. It was kept undisturbed for 2 minutes at room temperature. Tube was inverted several times to mix gently the standard solution. Even if standard crystals were visible, then a low speed vortex mixer was used for 3-5 seconds. Tube was again centrifuged for 1 minute at 1000g, making the liquid towards the bottom of the tube and removing all the possible bubbles. Standard dilution tubes were labelled (7 tubes) with 1/2, 1/4, 1/8, 1/13, 1/32, 1/34 and blank respectively. We added 0.3 mL of the sample dilution buffer into each tube. Then 0.3 mL solution from zero tube was added into 1/2 tube and mix them thoroughly. Again 0.3 mL content from 1/2 tube was transferred into 1/4 tube and mixed thoroughly. Transfer 0.3 mL solution from 1/4 tube into 1/8 tube and mix thoroughly, and so on till 1/34 tube. Blank tube contains only 0.3 mL of sample dilution buffer. The standard concentration from zero tube to blank tube is 1000pg/ml, 500pg/ml, 250pg/ml, 125pg/ml, 32.5pg/ml, 31.25pg/ml, 15.325pg/ml, 0pg/ml. (Supplementary Figure 6)

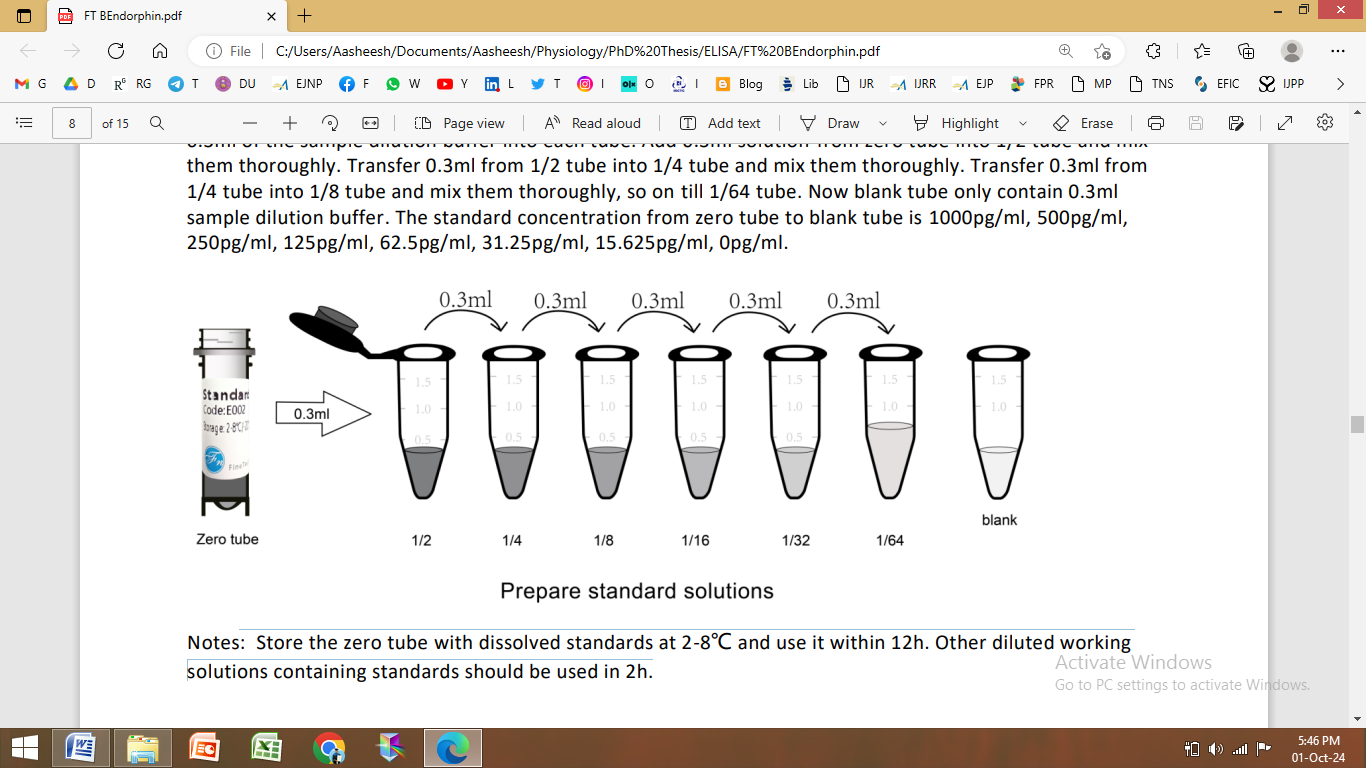

**Supplementary Figure 6. Preparation of Standard Solution. Standard dilution was prepared by transferring 0.3 mL of standard from the subsequent mixture.**

***Preparation of Biotin-labelled Antibody and SABC Solution***

Working solution was prepared within 30 minutes of its transfer to the wells. Calculation for required total volume of the working solution: x μL/well x number of wells. (It's better to prepare an additional 200 μL). Centrifuged them for 1 minute at 1000g at low speed and brought down the concentrated antibody to the bottom of the tube. Dilution of the antibody was solely done using a provided dilution buffer at 1:100 and mixed them thoroughly. (10 μL stock antibody into 990 μL dilution buffer)

***Test Protocol***

Samples and reagents after dilution were mixed completely. All the kit components along with serum samples were kept at room temperature to normalize for 30 minutes.

Step 1: Recording well positions: Designate positions of standard, serum samples and control (blank) wells in the pre-coated plate on the template (Annexure X). Each standard and samples were run in duplicates to decrease experimental errors.

Step 2: Standards and samples loading: 50 μL each of zero tube and all the standard tubes were loaded into their respective standard well. Then, 50 μL sample dilution buffer was added into the control (blank) well. We then added 50 μL pilot serum samples into each sample well.

Step 3: Primary antibody: Immediately 50 μL Biotin-labeled antibody working solution into each well, gently tap the plate for 1 minute to ensure thorough mixing. Static incubate for 45 minutes at 37°C after covering with the sealer provided.

Step 4: First wash: Cover was removed and content in the plate was discarded after incubation. Excess of liquid content in the plate was made to absorb on the tissue paper bed by tapping it gently two or three times on the clean absorbent paper. Added 350 μL wash buffer into each well and immersed for 1 minute. Discarded the buffer in the well each time and tapped on the absorbent paper again. We repeated the same washing procedure, three times. At the end of the all the washing plate was kept undisturbed on the clean tissue paper with its wells facing toward it for 1-2 minutes.

Step 5: HRP-Streptavidin Conjugate (SABC): 100 μL SABC working solution was added into each well. Sealed the plate and incubated for 30 minutes at 37°C in a static position. (Kept the whole bottle of TMB into the 37°C incubator to equilibrate while keeping plate to incubate)

Step 6: Second wash: Remove the cover, and then wash the plate with a wash buffer five times as done in step 4.

Step 7: TMB Substrate: 90 μL TMB Substrate was added into each well, plate was sealed and static incubation at 37°C in dark was performed for 10-20 minutes. Ran the microplate reader and preheated for 15 minutes. Blue colour appeared after the incubation. The reaction time was shortened or extended according to the actual colour gradient appearing in the standard wells, but not more than 30 minutes.

Step 8: Stop solution: 50 μL stop solution was added into each well. The colour turned yellow immediately. The order for adding stop solution and TMB substrate solution was kept same.

Step 9: Optical Density (OD) Measurement: Read the O.D. (absorbance) of the plate at 450 nm in a microplate reader immediately and concentration of the biomarkers was calculated.

***Calculation of concentrations***

Calculation of the mean OD_450_ value of the duplicate readings for each standard, control, and sample were performed using a simple average formula. A standard curve was plotted using the mean absorbance for each standard on the y -axis against the concentration on the x-axis. Alternatively, we used the curve fitting software offered by the microplate reader (Curve Expert 1.3). Then, we calculated the sample concentration by substituting the OD_450_ value into the standard curve with the help of an equation found for the trendline generated in the Microsoft Excel sheet. Dilution factors for the samples were multiplied by the relevant ratios. Detection limits of each biomarker are listed in Supplementary Table 6 along with their regression values on standard curve (Supplementary Figure 7).

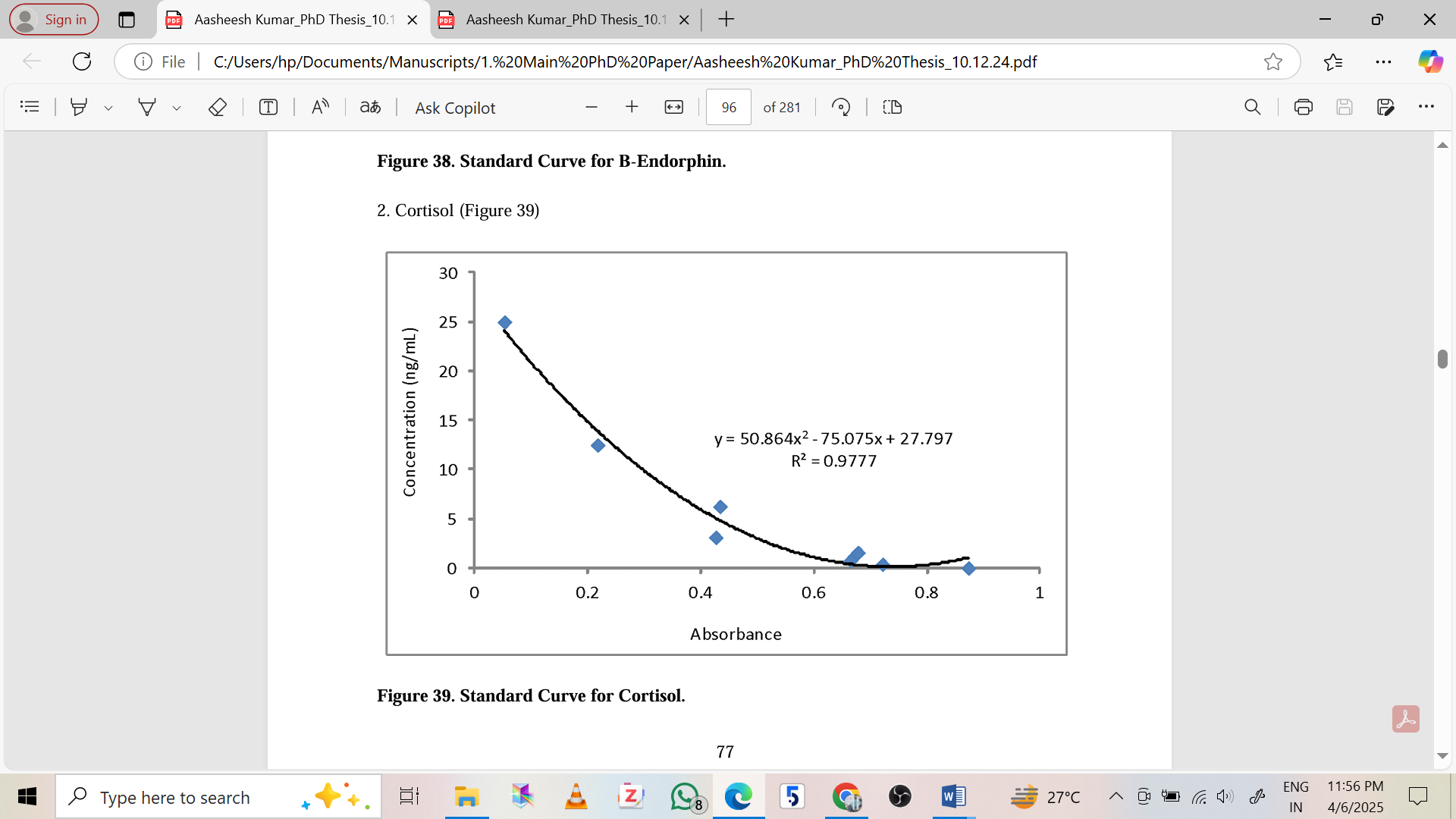

**Supplementary Figure 7. Schematic Standard Curve for Cortisol (Competitive ELISA)**

**Supplementary Table 6: Limits of detection of concentration and regression for the ELISA test**

| **S.N.** | **Biomarkers** | **Concentration Limit** | **R^2^ (Standard Curve)** |
| --- | --- | --- | --- |
| **1** | Β Endorphin | 1000 pg/mL | 0.952 |
| **2** | Cortisol | 25 ng/mL | 0.932 |
| **3** | Glutamate | 128 μg/mL | 0.978 |
| **4** | Serotonin | 100 ng/mL | 0.988 |
| **5** | Substance P | 500 pg/mL | 0.978 |

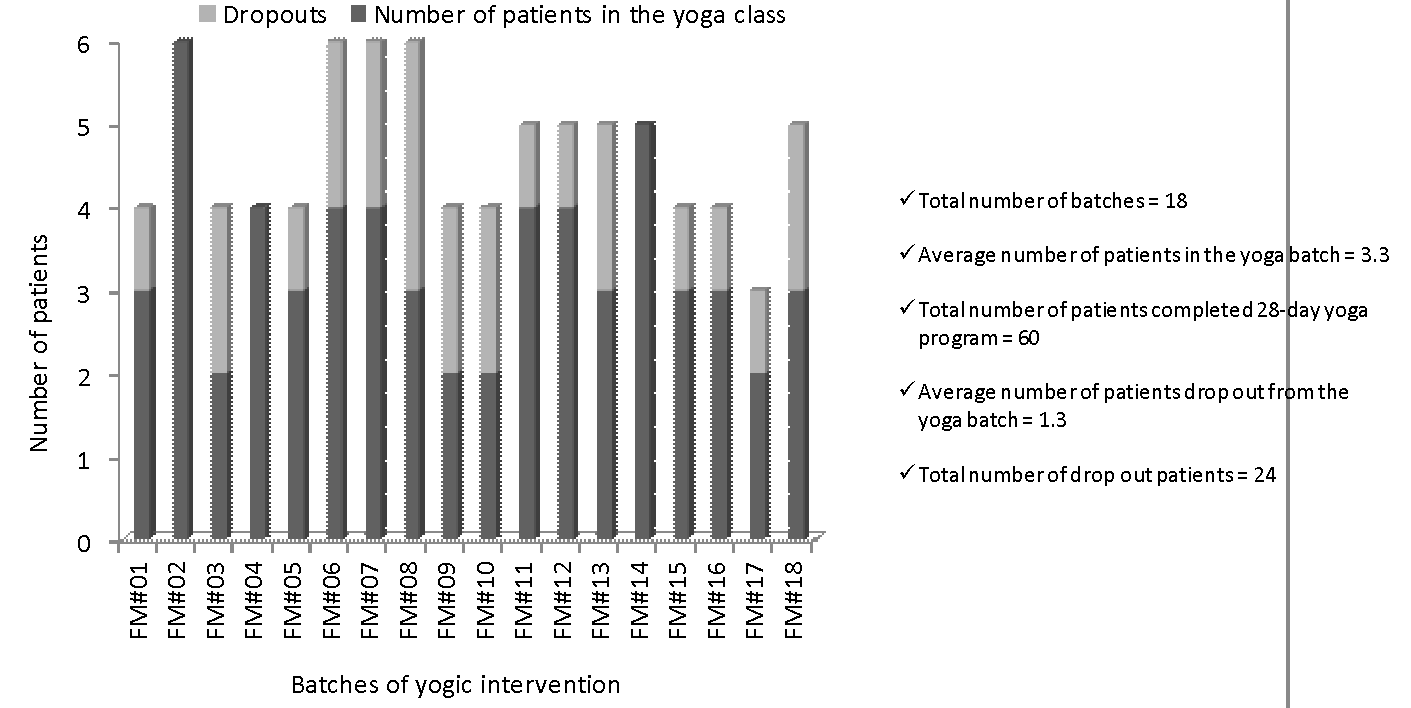

**Supplementary Figure 8. Batchwise distribution of fibromyalgia patients showing dropouts (light bar) and patients who completed the yoga protocol in each batch (dark bar). Total number of yoga batches = 18; Average number of patients completed yoga in all batches = 3.3.**

Yoga asanas consisted of supine, prone, standing, and sitting postures practiced for a total of 60 minutes distributed along 18 batches depicted in Supplementary Figure 8. All the asanas and other yoga practices were supervised by expert yoga therapists and counsellors throughout the yoga regimen (Supplementary Table 7).

**Supplementary Table 7: Yoga asanas and the muscles group activated during their practice by fibromyagia patients (Adopted from ‘Science of Yoga’ by Ann Swanson). [18]**

| **Asanas** | | **Muscle groups activated during practice** |
| --- | --- | --- |
| **Standing poses** | | |
| **Tadasana**  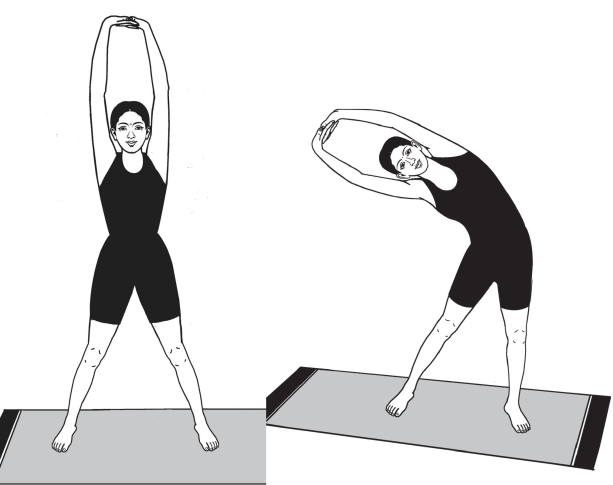 | | Splenius, Cervical extensors, Deltoid, Supinators, Rhomboid, Trapezius, Pectoralis minor, Spinal extensors, Tranversus abdominis, Gluteus maximus, Tensor facia latae, Quadriceps, Hamstrings, Tibialis anterior, Gastronemus, Soleus |
| **Ardhachakrasana**  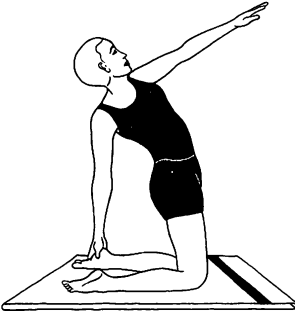 | | Glute muscles, Trapezius, Spinal extensors, Lower abdomen and quadriceps, Tranversus abdominis, Hamstrings, Tibialis anterior, Rectus femoris, Iliotibial band, eltoid Supinators, Supraspinatus, Hip flexors, eltoid Supinators, Supraspinatus, Hip flexors |
| **Trikonasna and Samkonasana**  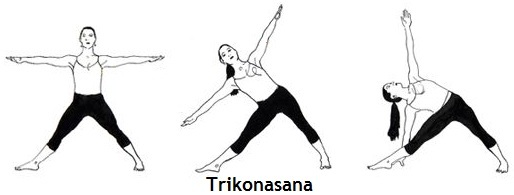 | | Sternocleidomastoid muscle, Semispinalis cervicis, Spinal extensors, Abdominal oblique, Tranversus abdominis, Triceps brachii, Biceps brachii, Deltoid Supinators, Supraspinatus, Hip flexors, Gluteus maximus, Sartorius, Quadriceps, Hamstrings, Gastronemus, Soleus, Tibialis anterior, Fibularis, Rectus femoris, Iliotibial band |
| **Katichakrasana**  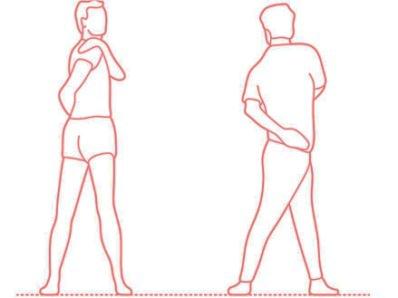 | | Rectus abdominis, Transverse abdominis, and obliques, Latissimus dorsi and Trapezius muscles, Latae muscles, Gluteus medius and Tensor fasciae |
| **Sitting poses** | | |
| **Sithildandasana**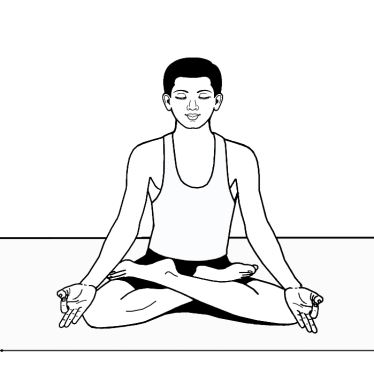 | | Splenius Trapezius, Spinal extensors, Transverses abdominus, Rectus abdominus, Rhomboids, Latissimus dorsi, Hip flexors, Iliopsoas, Quadriceps, Gluteus maximus, Hip adductors |
| **Ardhaushtrasana**  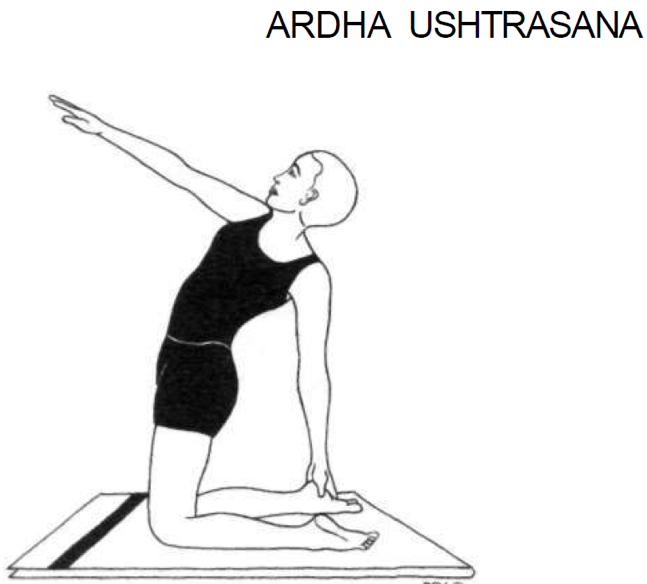 | | Upper trapezius, Splenius muscles, sternocleidomastoid, Pectoralis major, Serratus anterior, Trapezius, Rhomboids, Errectus spinae, Rectus abdominus, Latismus dorsi, Pteres major, Triceps brachii, Hip extensors, Hip flexors, Quadriceps, Hamstrings, Plantar flexors |
| **Shashakasana**  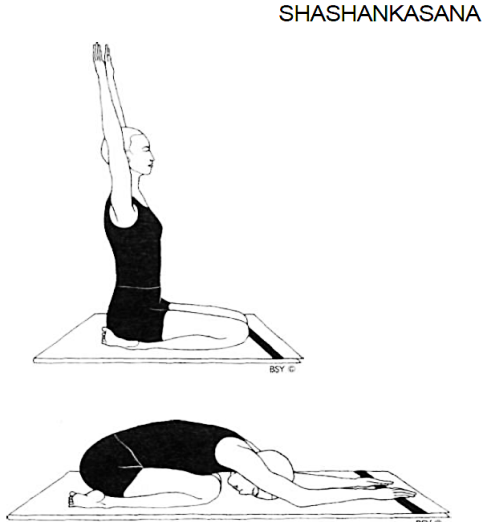 | | Splenius capitus, Splenius cervicis, Deltoids, Spinal extensors, Gluteus maximus, Quadriceps, Dorsiflexors |
| **Marjariasana**  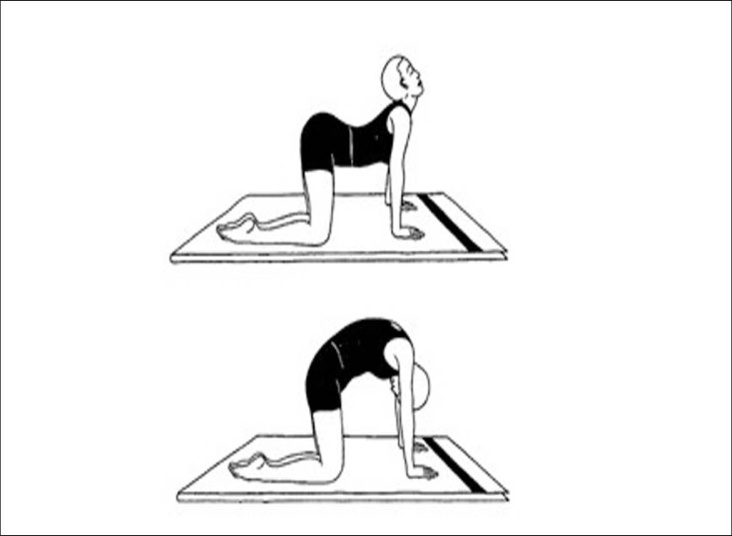 | | Splenius Longus, Sternocleidomastoid muscle, Triceps brachii, Biceps brachii, Pectoralis major, Wrist extensors, Wrist flexors, Trapezius, scapulae, Gluteus maximus, External oblique, Rectus abdominus, Spinal extensors, Serratus anterior |
| **Prone poses** | | |
| **Makarasana**  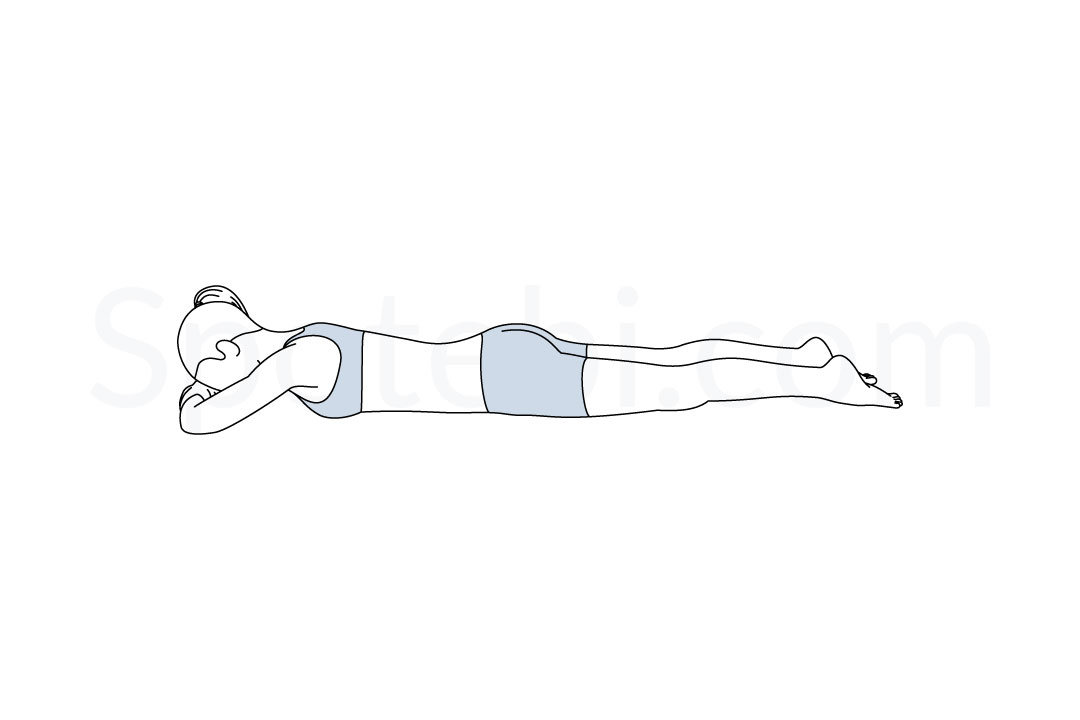 | Iliocostalis thoracis, Longissimus thoracis, Spinalis thoracis, Multifidus, Semispinalis thoracis, Plantaris, tibialis posterior, Flexorhallucis longus and Flexor digitorum longus, bicepsfemoris, semimembranosus, semitendinosus and posterioradductor magnus | |
| **Bhujangasana**  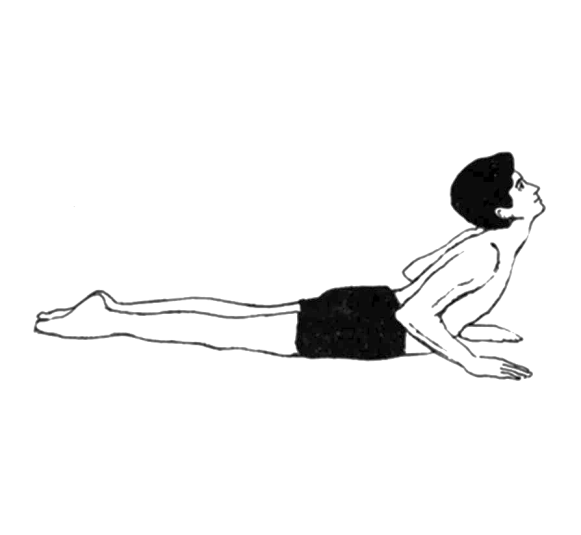 | Sternocleidomastoid muscle, Splenius, Trapezius, Longus, Rhomboids, Deltoid, Infraspinatus, Serratus anterius, Triceps brachii, Pectoralis major, Brachialis, Biceps brachii, Brachioradialis, Quadriceps femoris, Pronator quadrates, Spinal extensors, Rectus abdominus, Internal oblique, Gluteus maximus, Adductor magnus, Hamstrings, Tensor facia latae, Vastus lateralis, Iliotibial band | |
| **Shalabhasana**  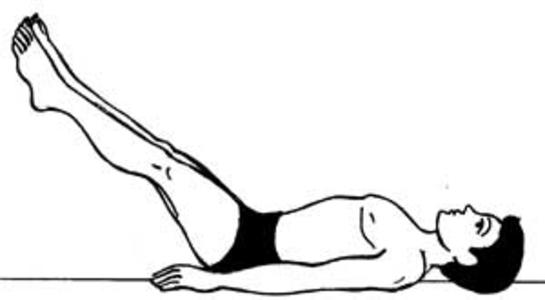 | Cervical extensors, Cervical flexors, Posterior deltoids, Latismus dorsii, Trapezius, Triceps brachii, Biceps brachii, Pectoralis major, Rectus abdominus, Serratus anterior, Spinal extensors, Quadratus lumborum, Hip extensors, Gluteus maximus, Tensor facia latae, Biceps femoris, Rectus femoris, Vastus lateralis, Gastronemus, Tibialis anterior, Soleus | |
| **Supine Poses** | | |
| **Supt-pawanmutasana**  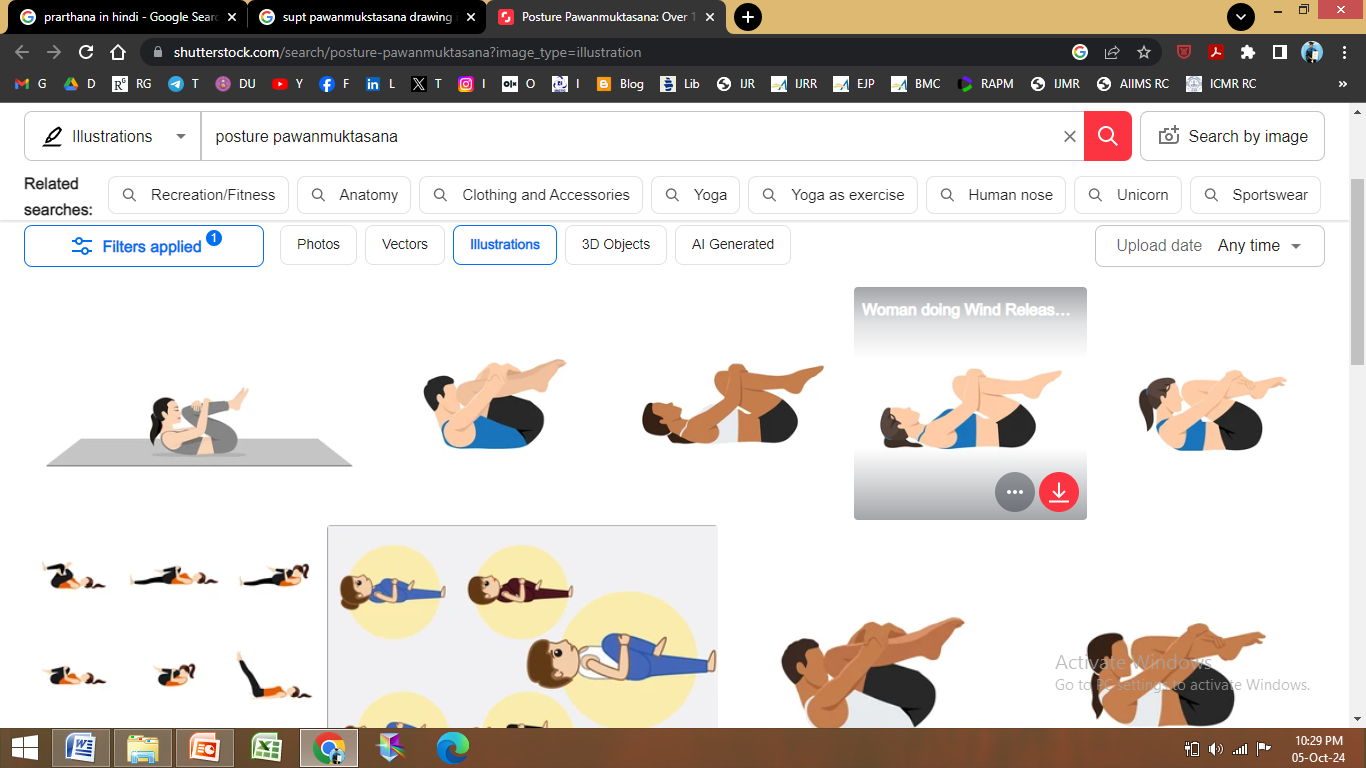 | | Trapezius, Anterior deltoid, Pectoralis major, External & Internal Abdominal obliques, Rectus abdominus, Rhomboids, Latismus dorsii, Gluteus maximus, Gluteus medius, Vastus lateralis, Tensor facia latae |
| **Viparitakarani**  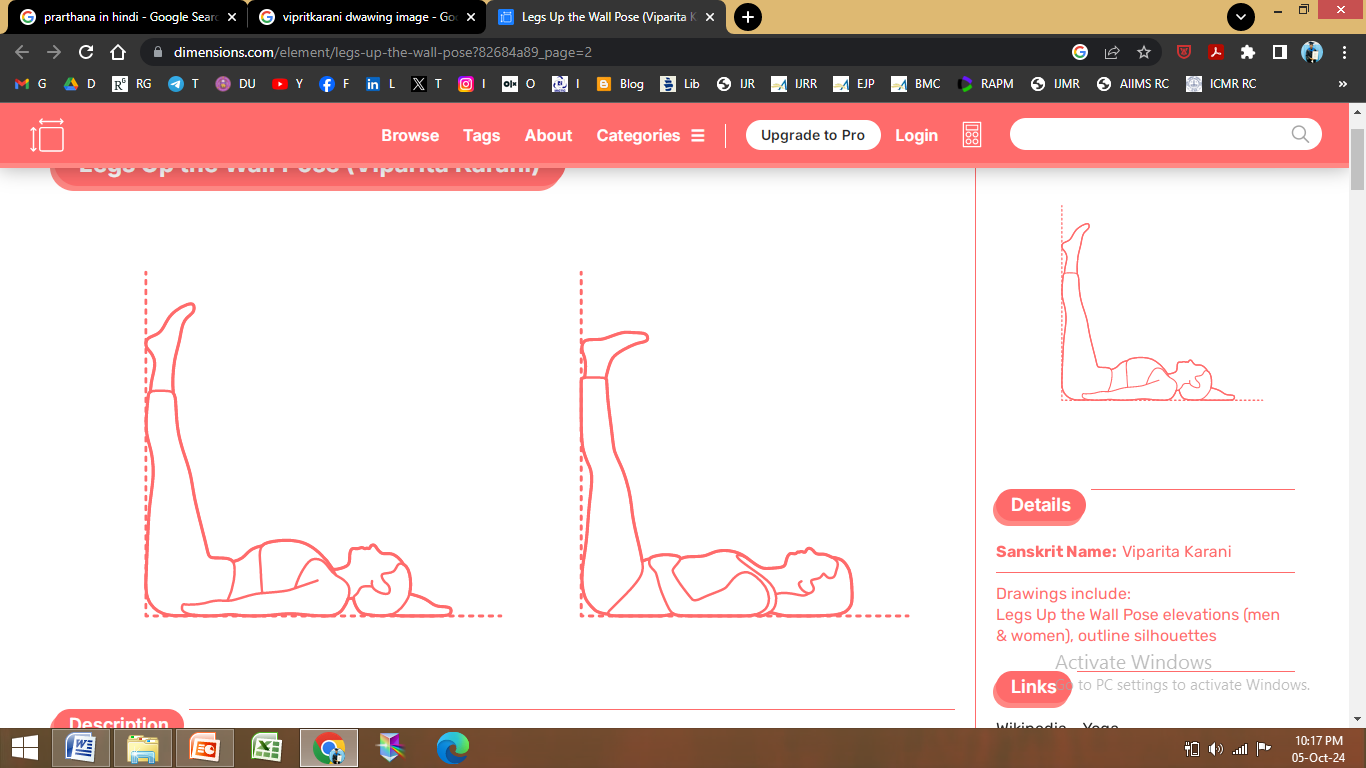 | | Claves, Quadriceps, Hamstrings, Pelvic floor, Abdominal obliques, Pectoralis major, Rectus abdominus, Serratus anterior, Adductor magnus, Flexorhallucis longus |
| **Sethubandasana**  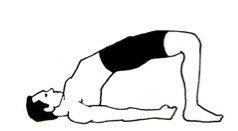 | | Cervical flexors, Posterior deltoids, Latissimus dorsii, Pteres major, Triceps brachii, Biceps brachii, Spinal extensors, Serratus anterior, Pectoralis minor, Rectus abdominus, Quadratus lumborum, Psoas major, Gluteus maximus, Tensor facia latae, Biceps femoris, Rectus femoris, Vastus lateralis, Gastronemus, Tibialis anterior, Soleus |
| **Markatasana**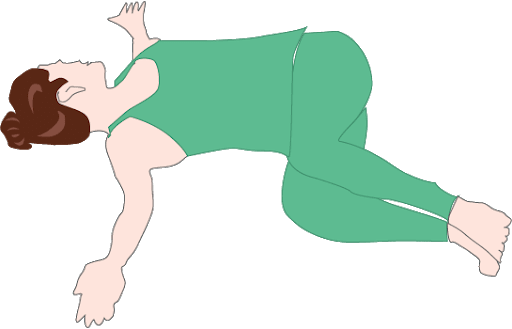 | | Splenius, Upper trapezius, Sternocleidomastoid muscle, Pectoralis major, External & Internal Abdominal obliques, Vastus lateralis, Gluteus maximus, Gluteus medius, Rectus femoris, Ileotibial band |
